# Is any job better than no job? : A systematic review

**DOI:** 10.1101/2021.11.23.21266736

**Authors:** Sian Price, Hannah Shaw, Fiona Morgan, Rocio Rodriguez Lopez, Kirsty Little, Ciarán Humphreys

## Abstract

**Objectives:** This systematic review addresses the question “Is any job better than no job?” Specifically, it compares health and well-being outcomes in those who are unemployed with those who are in jobs that could be considered poor or low quality and the impact of any movement between them.

**Method:** We conducted a systematic review following a PROSPERO-registered protocol (CRD42020182794). Medline, Embase, PsycINFO, HMIC, ASSIA, TRIP, Google Scholar and 10 websites were searched in April 2020 and again in May 2021 without date limits. Two reviewers working independently screened search results against the inclusion/exclusion criteria. A checklist for quantitative studies reporting correlations was used to critically appraise articles included at full text. We undertook synthesis without meta-analysis (narrative synthesis) and explored a range of variables (for example, study design and quality, type of outcome measure) that we considered might have an impact on the association between exposure and outcome.

**Results:** We included 25 studies reported in 30 journal articles. All 25 studies involved secondary analysis of data from national cohorts, including six from the UK. The most frequent outcomes reported were measures of mental well-being. There was considerable heterogeneity across included studies in terms of design, population, definition of poor/bad or low quality job and outcome types and measures. Overall the quality of the included studies was moderate. The evidence base is inconsistent. There are studies that suggested either labour market position might be preferable, but a number of studies found no statistically significant difference. Cohort and case-control studies looking at mental well-being outcomes showed some support for a poor job being better than unemployment. However, we did not find sufficient numbers of well-designed studies showing a strong association to support a causal relationship. Most included study designs were unable to distinguish whether changes in employment status occurred before a change in outcome. Three studies looking at employment transitions found that moving to a poor job from unemployment was not associated with improved mental health, but moving from a poor job to unemployment was associated with a deterioration.

**Conclusion:** Evidence that better health and well-being outcomes are more likely to be associated with a poor/bad or low quality job than with unemployment is inconsistent. Studies conducted in the UK suggest that a poor job is not significantly associated with better health and well-being outcomes than unemployment. The studies we identified do not allow us to distinguish whether this lack of association is the result of a state welfare regime preventing some of the worst ills associated with unemployment, or a reflection of job quality. The evidence base has significant limitations in study design and conduct. In summary, the evidence we found suggests it is not safe to assume that, in the UK, any job will lead to better health and well-being outcomes than unemployment.

## Introduction

In 1999 the International Labor Organization (ILO) made “securing decent work for all” its “primary goal” for the new millennium. This was defined as “productive work for women and men in conditions of freedom, equity, security and human dignity” (ILO, 1999).

There is no universally recognised definition of what constitutes a decent job, but discussions on the quality of work usually encompass employment stability, task control, workplace safety and appropriate remuneration. In *Fair Society, Healthy Lives* (Marmot et al, 2010) cited 10 core components of good work. These included jobs:

- Free of the core features of precariousness (lack of stability with high risk of job loss, lack of employment protection and of physical safety measures)
- Allowing an element of control with the worker having some influence on aspects of their work such as timing and tasks to be undertaken
- Placing appropriately high demands on the worker in terms of quantity and quality of work without harming their physical or mental well-being
- Fair, earnings reflecting productivity with employers committed towards guaranteed job security
- Offering opportunities for skills development and promotion
- Preventing social isolation, discrimination and violence
- Participation in organisation decision making and collective bargaining with guaranteed procedural justice in case of conflicts
- Aiming to support work life balance
- Supporting integration/reintegration into full employment of those with ill health and long-term conditions
- Supporting workers to meet their needs for self-efficacy, self-esteem, sense of belonging and meaningfulness

The last twenty years has seen a significant increase in atypical forms of working, much of it insecure: zero-hours contracts, casualisation and platform work where workers are considered freelance or independent contractors in a ‘gig economy’ (Taylor et al, 2017). Issues arising from this led to the UK government commissioning a review of modern working practices in 2016. This review, chaired by Matthew Taylor, was to look at how employment law needed to adapt to keep pace with modern employment practices. It concluded that “all work in the UK economy should be fair and decent with realistic scope for development and fulfilment.” (Taylor et al, 2017). Devolved administrations in Scotland and Wales have also been looking into this with a Fair Work Convention in the former (2015) and a Fair Work Commission (2019) in the latter.

Whilst much of this work was in process, the UK Trades Union Congress (TUC) published *Living on the Edge* (2016). This report indicated that 3.2 million people in the UK were part of an “insecure workforce” who experienced fewer rights and protections, were in low paid self-employment, various forms of temporary work or on zero-hours contracts (The TUC specifically excluded workers on fixed term contracts as they had the same rights as those afforded to those employed on permanent contracts).

It is widely accepted that employment is beneficial for health and well-being (Carré et al, 2012, Waddell and Burton, 2006). However, increasingly the importance of job quality and the differential impacts of good or poor jobs is being recognised. The Taylor Report clearly stated that “while having employment is itself vital to people’s health and well-being, the quality of people’s work is also a major factor in helping people to stay healthy and happy…” (Taylor et al, 2017, P6)

A conclusion of *Fair Society, Healthy Lives* (Marmot et al, 2010) was that good employment is usually protective of health, whilst unemployment, particularly in the long-term, is a significant contributor to poor health. Poor quality work, or a bad job, was recognised as a significant contributor to poor health, particularly mental health.

Since 2010, the labour market has continued to change, as indicated in the various government reports noted above. In a follow up report, *Health Equity in England*, Marmot et al (2020) highlighted that while rates of unemployment had decreased, changes in employment practices meant that work is more often low-paid, unskilled, self-employed, short-term or zero hours contract jobs. The report also noted that rates of pay have not increased and more of those in poverty are now in rather than out of work. All these factors have clear implications for health and wellbeing.

A number of systematic reviews have explored the relationship between labour market status and health and well-being. One from 2014 (van der Noordt et al) reported a strong protective effect for employment on both depression and mental well-being. However, the review found insufficient evidence for impact on general physical health, general health and other health outcomes.

More recent systematic reviews have explored the impact of quality of employment. Amiri and Behnezhad (2020) looked at the relationship between job strain and mortality. Job strain was characterised as work placing high demands on an individual who had low job control. Results suggest that, in European studies, there was an increase in risk of mortality for those exposed to job strain. The increased risk in women alone was not significant, nor was there an increase in risk found in studies conducted in the USA.

A review published in 2015 (Kim and von dem Knesebeck) explored the health-related risk of job insecurity and unemployment. Job insecurity was based on self-reports rather than any objective measure. Review authors reported that the evidence was inconsistent, with both exposures being associated with impaired health. They found a slightly stronger association between job insecurity and somatic symptoms, whilst general health and increased mortality was worse with unemployment. They reported a strong association between mental health and both exposures, but this finding was inconsistent with no clear direction of effect.

Rönnblad et al (2019) looking at precarious employment and mental health, concluded that job insecurity is likely to have an adverse effect on mental health. However, the authors noted the lack of a clear multi-dimensional definition of precarious employment and lack of high-quality prospective studies with policy-relevant results. A subsequent review looking at the same topic (Utzet et al, 2020) attempted a more multi-dimensional perspective on precariousness and one that took gender and household situation into account. This review recognised that the relationship between mental health and employment status is complex. Of the included studies, 20 considered job insecurity, 12 temporariness, 10 used a multidimensional approach (subdivided by the reviewers into job quality approach and other multi-dimensional approaches), nine working time arrangements, five downsizing and major organisational restructuring and four remuneration.

Most of the studies looking at job insecurity, temporariness and those using multi-dimensional approaches reported a significant association with mental health problems. However, results for working time arrangements and downsizing were inconclusive and were inconsistent for sex dependent differences in the impact of precarious employment.

A 2018 review (Koranyi et al) looked at precarious employment and occupational accidents and injuries. It explored aspects of precarious employment. Specifically temporary employment, multiple jobs, working for a subcontractor at the same worksite/temp agency, part-time, self-employment, hourly pay, union membership, insurance benefits, flexible versus fixed work schedule, wages, job insecurity, work-time control and precarious career trajectories. The review authors concluded there was an association between some aspects of precarious employment and injuries, notably multiple jobholders and employees of temporary employment agencies or subcontractors.

Finally, Kim and colleagues (2012) undertook a review looking at whether welfare states act as a mediator between flexible employment (in this case job insecurity and precarious employment) and health outcomes (chronic disease, mental health, health behaviours). They did not look at the impact of unemployment. Included studies were allocated to one of six welfare state regimes. They found that precarious workers in Scandinavian welfare states reported better or equal health status to those in permanent employment. However, precarious work in the other welfare state regimes (including the UK) was associated with worse outcomes.

Currently jobs and employment prospects appear to be improving, although there are still uncertainties. As at September 2021, figures from the Office for National Statistics indicated that the number of pay-rolled employees had returned to pre-pandemic levels. (Leaker, 2021). There were also reports of double digit pay rises in a few sectors - transport and food processing - because of a shortfall of labour. (Nabarro, 2021). However, despite projections that the UK will achieve pre-pandemic levels of economic activity by early 2022, the Office for Budget Responsibility (2021) sees future UK economic prospects being subject to several pressures. Brexit is expected to reduce the size of the British economy by about 4% in the longer term, with the pandemic cutting UK GDP by a further 2%. Added to this, inflation is expected to peak at around 5% in 2022.

Nabarro (2021) highlights the likely impact of profound economic adjustments arising from Covid-19 and Brexit. The pandemic has led to significant longer term changes in household spending, whilst Brexit is inevitably changing UK trade patterns. Labour market changes have yet to catch up with these new realities. Crossley et al (2021) note that longer term, the pandemic appears to have had a greater impact on older workers, with younger people significantly more likely to have started working again either in their previous employment or in a new role. This is particularly salient given that research suggests young jobseekers see opportunities in under-resourced sectors such as retail, transport and storage, social care, food processing, hospitality and construction, as insecure jobs that are physically demanding with long hours, low pay and little likelihood of promotion. (Davies, 2021).

Given the effects of the pandemic and Brexit on the UK economy in the longer term, the Westminster government and the devolved administrations will need to make efforts to maintain and create employment. However, in order to maximise the benefits of employment for health and well-being the quality of employment will also need to be carefully considered. An individual’s employment and mental and/or physical health status will be closely linked and subject to a range of other determinants. This has been clearly noted in the recently published review of health equity in Greater Manchester (Marmot et al, 2021).

Systematic reviews have looked at aspects of what might be considered poor or bad employment and its relationship with health and well-being outcomes. These suggest that there are differential impacts on health and well-being depending on the nature or quality of work. However, none compared the relationship with unemployment against that of low quality or poor jobs. The possibility that some jobs may have a negative, rather than a positive, impact on health and well-being needs consideration. Whilst a move from unemployment to paid work may lead to an improvement in health outcomes, other events and alternative relationships and relationship directions between labour market and health status should also be considered.

To investigate these relationships, this systematic review looks to answer the question “Is any job better than no job?” Specifically it compares health and well-being outcomes in those who are unemployed with those who are in jobs that may be considered poor or low quality and the impact of movement between these.

## Method

A protocol describing *a priori* the methods used to conduct this systematic review was registered in the PROSPERO database (CRD42020182794).

### Searching

We searched Medline, Medline In-Process & Other Non-Indexed Citations and Daily & Medline Epub Ahead of Print, Embase, PsycINFO, Health Management Information Consortium (HMIC), Applied Social Sciences Index and Abstracts (ASSIA), TRIP database and Google Scholar and 10 websites in April 2020. To ensure currency, an update search was conducted in May 2021. Full details are in the supplementary material. No limits were placed on year of publication, but only sources with an English language abstract were included.

### Screening

Screening, against the criteria in Table 1, was conducted independently at title, abstract and full text by two reviewers. Disagreements were resolved through discussion, involving a third reviewer where necessary. We checked relevant systematic reviews for missed studies. Details of the studies we excluded at full text with reasons for exclusion are in the supplementary material.

**Table 1:** Inclusion/exclusion criteria.

| **Inclusion** | **Exclusion** |
| --- | --- |
| **Study type**<br>Primary studies of observational, case control, cross sectional or cohort that include a relevant exposure and comparator | **Study type**<br>Systematic reviews<br>Studies that do not include a relevant exposure and comparator |
| **Source type**<br>Primary studies reported in journal articles, grey literature, dissertations and conference proceedings | **Source type**<br>Other types of source |
| **Population**<br>Adults of working age in OECD countries<br>Studies including data from samples drawn from a national population | **Population**<br>Studies only including population sub groups, for example those with disabilities, chronic disease, migrant workers, older workers, specific occupational groups<br>Studies where the sample is not drawn from a national population |
| **Exposure**<br>Poor or low quality work however defined. Including but not limited to low paid work, work perceived as low status, work that is insecure or temporary, work presenting physical risks, work where workers rights are limited, work that adversely affects family life | **Exposure**<br>Work, however defined, not considered by the study authors to be poor or low quality or potentially detrimental to health and well-being, for example work described as secure or permanent employment |
| **Comparator**<br>Unemployed (and actively seeking work) | **Comparator**<br>Unemployed and not actively seeking employment<br>Not in the workforce for other reasons (e.g. long-term conditions, stay at home carers) |
| **Outcome**<br>Any self-reported or objective measures of physical and mental health or mental well-being, or health behaviours<br>Any family or community impacts | **Outcome**<br>Studies only reporting measures of job satisfaction |

### Data extraction

Data was extracted into a standard template and included source, study design, sample, data collection method, definition of employment quality, outcome measures and covariates, method of analysis, key results and information about the social security arrangements in the country where the study took place. One reviewer extracted data, this was checked by a second.

### Quality assessment

Two reviewers independently assessed the quality of all included studies. Disagreements were resolved thorough discussion, involving a third reviewer where necessary. We used a checklist for quantitative studies reporting correlations (National Institute for Health and Care Excellence, 2012) as this was appropriate for use with all our included study designs. Most included studies used data from national cohorts so additional sources (journal articles and study websites) were needed in order to undertake critical appraisal. We made some modifications to the checklist to accommodate this (details available on request). We tested the checklist to ensure that both reviewers had a common understanding of the questions. There are a number of likely confounders (for example education and deprivation) but we considered that the critical confounders (covariates associated with both the exposure and outcome) would be age and underlying chronic health conditions.

Using the criteria and rating system in the checklist, each study was given an overall grading for internal validity (IV), (the extent to which the study measures what it is intended to measure) and a separate one for external validity (EV), (generalisability, the extent to which the findings of the study are likely to apply to other settings):

- ++ (rated good) all or most of the checklist criteria have been fulfilled, where they have not been fulfilled the conclusions are very unlikely to alter
- + (rated moderate) some of the checklist criteria have been fulfilled, where they have not been fulfilled, or not adequately described, the conclusions are unlikely to alter
- - (rated poor) few or no checklist criteria have been fulfilled and the conclusions are likely or very likely to alter.

### Synthesis

Because there was considerable heterogeneity between studies we undertook synthesis without meta-analysis. We explored a range of variables we considered might have an impact on the association between exposure and outcome.

Where included studies did not directly compare poor job and unemployment but appropriate data were available, we extracted this and calculated an unadjusted relative risk (RR) (for longitudinal studies) or odds ratio (OR) (for cross sectional studies). Where there was no appropriate comparison or data, we judged whether the difference between the outcomes for poor work and unemployment was likely to be significant based on overlap (or not) of confidence intervals of the effect sizes.

## Results

### Study selection

The flow of literature through the selection process is in Figure 1.

**Figure 1:**
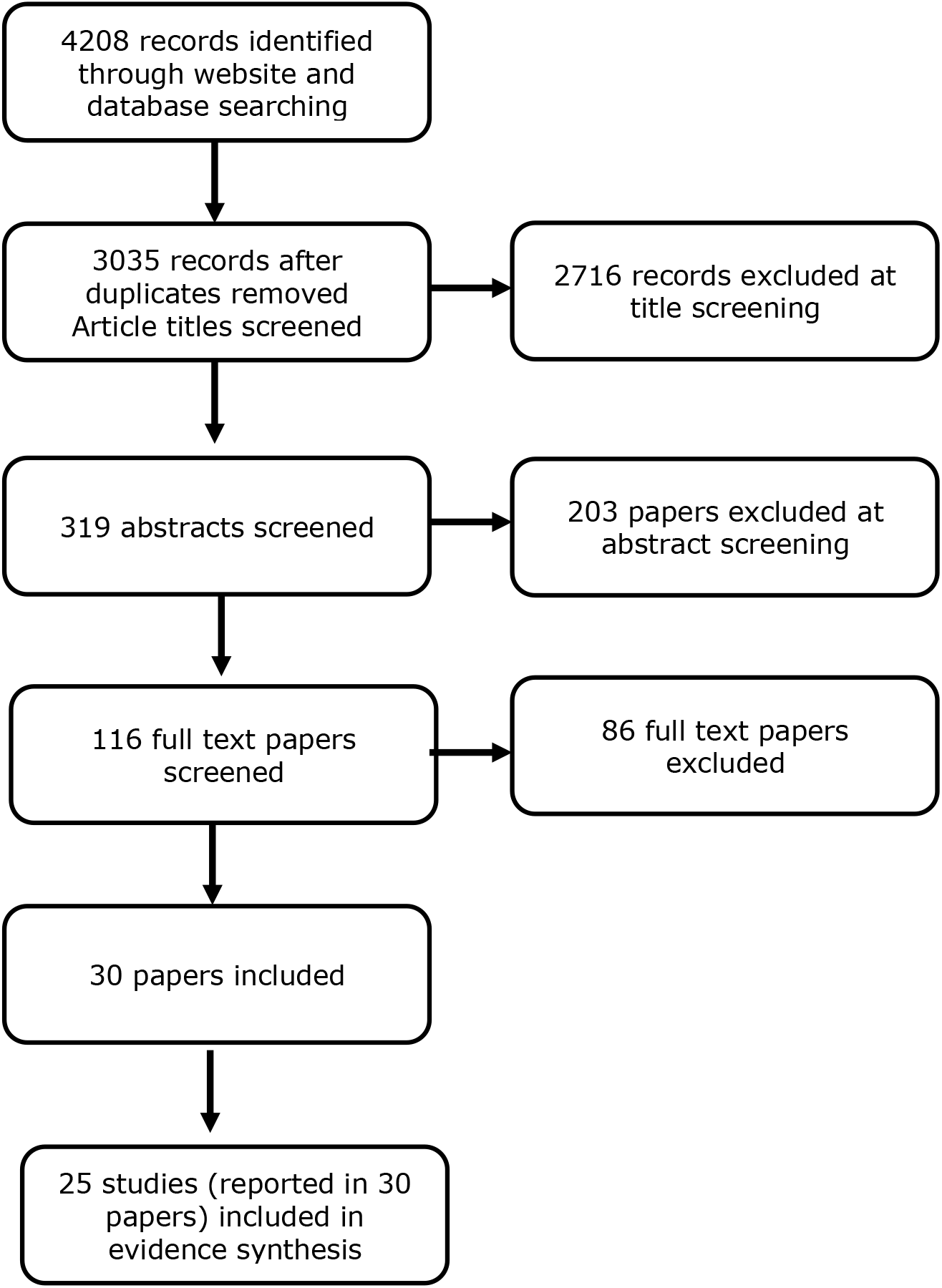
Flow of information through the review.

### Characteristics of included studies

Table 2 provides the main characteristics of the included studies.

**Table 2:** Characteristics of included studies.

| **Study** | **Int/ext validity** | **Population age range** | **Data source** | **Poor job** | **Outcome** |
| --- | --- | --- | --- | --- | --- |
| Bently et al. 2020 (Australia)<br>Cross sectional | +/+ | 25 to 64 | Household Income and Labour Dynamics in Australia 2018 | Insecure employment; included self-employment, causal or labour hire but not fixed term contracts (Simple) | Alcohol and tobacco use |
| Butterworth et al. 2013 (England)<br>Cross sectional | +/+ | 21 to 54 | English Adult Psychiatric Morbidity Survey, 2007 | Psychosocial job quality; demands, security, esteem from colleagues, clients, customers and managers, job control (Complex) | Common mental disorders (CIS-R) |
| Butterworth et al. 2011 (Australia)<br>Cohort | ++/+ | 20 to 55 at wave 1 | Household, Income and Labour Dynamics in Australia 2001 to 2007 | Psychosocial job quality; demand, complexity, control, security, hours and shift work (Complex) | Mental health (SF-36) |
| Chandola and Zhang 2018 (UK)<br>Cohort | +/+ | 35 to 75 | Understanding Society, UK Household Longitudinal Survey 2009 to 2012 | Job quality; earnings, security, quality of working environment (Complex) | Allostatic load – biomarkers related to chronic stress. Physical and mental well-being (GHQ-12, SF-36) |
| Cortes-Franch et al. 2019 (Europe)<br>Cross sectional | +/+ | 16 to 64 | European Social Survey 2010 | Low quality job; earnings, prospects, intrinsic job quality, working time quality, participation and representation (Complex) | Mental well-being (WHO-5 well-being index) |
| Cortes-Franch et al. 2018 (Spain)<br>Cross sectional | +/+ | 25 to 64 | Spanish National Health Survey 2006 to 2007 | Insecure employment, temporary or no contract (Simple) | Mental health (GHQ-12) |
| Fiori et al. 2016 (Italy)<br>Cross sectional | +/+ | 18 to 39 | Health Conditions and Access to Health Services Survey, Italy, 2005 and 2013 | Insecure employment (Simple) | Mental health (SF-12) |
| Flint et al. 2013 (UK)<br>Cohort | +/+ | 16 to 65 | British Household Panel Survey 1991 to 2007 | Insecure employment (Simple) | Psychological well-being (GHQ-12) |
| Fornell et al. 2018 (Spain) Cross sectional | +/+ | 16 to 65 | Survey of Living Conditions, Spain, 2007 to 2011 | Insecure employment (Simple) | General health (self-report) |
| Gebel and Vosemer 2014 (Germany) Case control | ++/+ | 16 to 54 | German Socio-Economic Panel 1995 to 2010 | Insecure employment (Simple) | Psychological and physical health (SF-12 and self-report) |
| Griep et al. 2016 (Finland) Cross sectional | +/- | 18 to 64 | Living Conditions Survey, Finland, 1994 | Perceived job insecurity (Simple) | Psychological complaints, self-rated health, life satisfaction (all self-report) |
| Grzywacz and Dooley 2003 (USA) Cross sectional | -/- | 25 to 74 | National Survey of Midlife Development, USA, 1995 | Inadequate employment; low pay and psychological aspects (Complex) | Physical health (self-report), depression (CIDI-SF) |
| Guseva Canu et al. 2019 (Switzerland) Cohort | +/+ | 18 to 65 | Swiss census data 1990 to 2004 | Occupational status lowest skilled and unskilled employees (Simple) | Suicide |
| Inanc 2018 (UK) Cross sectional | +/+ | 20 to 65 | British Household Panel Survey, 8 waves 1991 onwards | Insecure employment (Simple) | Psychological well-being (GHQ-12) and life satisfaction (single survey question) |
| Jang et al. 2015 (South Korea) Cohort | +/- | 19 to 65 | Korean Welfare Panel Study 2007 to 2013 | Insecure employment (Simple) | Severe depressive symptoms (CES-D) |
| Kim et al. 2019 (South Korea) Cohort | -/- | 25 to 59 | Korean Welfare Panel Study 2008 to 2011 | Insecure employment (Simple) | Suicidal ideation in the past year (single question) |
| Kim et al. 2020 (South Korea) Cross sectional | +/- | 19 to 65 | Korean National Health and Nutrition Examination Survey, 2015 | Precarious employment (Simple) | C-reactive protein (inflammatory marker) |
| Maeda et al. 2019 (Japan)<br>Cross sectional | -/- | 20 to 59 | Comprehensive Survey of Living Conditions, Japan, 2010 | Not regularly employed – not further defined (Simple) | Insomnia related symptoms (self-report) |
| Matilla-Santander et al. 2020 (Europe)<br>Cross sectional | -/- | 15 and over | European Working Conditions Survey, 6<sup>th</sup> wave, 2015 | Precarious employment; temporariness, not being able to exercise rights, vulnerability, disempowerment and wages (Complex) | Bad health, health problems (self-report) |
| Minelli et al. 2014 (Italy)<br>Cross sectional | -/- | 15 to 64 | Survey on Household Income, Italy, 2006 to 2010 | Insecure employment (included apprenticeships, on-project jobs and seasonal jobs)(Simple) | General health (self-report) |
| Park et al. 2020 (South Korea)<br>Cross sectional | +/- | 15 to 59 | Korean National Health and Nutrition Survey, 2013 to 2017 | Insecure employment (Simple) | Mental health (PHQ-9), general health (EuroQol - 5D), health behaviours (self-report) |
| Scheuring et al. 2021 (Germany)<br>Cohort | +/+ | 18 to 65 | German Socio-Economic Panel 1995 to 2017 | Fixed term employment (Simple) | Life satisfaction (self-report) |
| Sumner et al. 2020 (UK)<br>Cross sectional | -/- | 16 to 64 | Understanding Society Survey, UK, 2010 to 2011 | Insecure employment (Simple) | Biomarkers C-reactive protein and fibrinogen |
| Van Aerden, Gadeyne and Vanrolen 2017 (Belgium)<br>Cross sectional | -/- | 18 to 64 | Belgian Generations and Gender Study, 2008 to 2010 | Precarious employment ; unstable, low income and involuntary part-time employment (Complex) | Mental health, general health (self-report) |
| Yoo et al. 2016 (South Korea)<br>Cohort | +/- | 18 years and above | Korean Welfare Panel Study 2008 to 2011 | Insecure employment (Simple) | Depression (CES-D) |

We included 25 studies reported in 30 papers in the review. Seven of the included papers used data from the Korean Welfare Panel Study (KOWEPS) (Jang et al, 2015; Kim et al, 2012; Kim et al, 2013; Kim et al, 2019; Kim and Park 2021; Yoo et al, 2016 and Yoon et al, 2020). These were very similar but had slight differences in either baseline population, outcome, dates or analysis. We extracted data from them all. However, based on the recency and duration of the reported data, we have only included Jang et al (2015), Kim et al (2019) and Yoo et al (2016) in the synthesis.

All the studies we included used data from national cohorts but many were cross sectional in design. Where the study analysis compared outcomes for bad and no job at a single time point/survey round we considered studies to be cross sectional regardless of how the study authors identified them. Our synthesis therefore included 25 studies, eight cohorts, one case control and 16 cross sectional studies.

Six studies were conducted in the UK; one study included 35 European countries, two each in Italy, Germany, Spain and Australia one each in Switzerland, Belgium, Japan and the USA. The remaining six studies were conducted in South Korea The most common outcome was mental well-being; this included general measures of mental or psychological well-being, measures of depression, suicide ideation and suicide. Measures were predominantly self-report, however, some of these used validated tools. Other outcomes included general health, physical health, health behaviours and injuries. Three studies reported bio-marker outcomes; two studies reported impact on partner, but we found no studies reporting other family outcomes.

We only included studies that were representative of national populations who were active in the labour market (working or seeking employment). Age ranges across the individual studies varied quite considerably, as did sample sizes. The labour market status of the participants included in the analysis relevant to our question also varied; some included all those active in the labour market, but others only included specific types of employment or all those who were not in paid work. Others, generally cohort studies, looking at employment transitions, varied in the participants included at baseline.

We did not asses publication bias however; findings in either direction are likely to be of equal interest, so we felt it was unlikely to be a significant issue.

### Definition of poor job

The definitions used varied across included studies. We grouped these as complex if they were multidimensional and simple where they were not (Table 2). None of the definitions used in the included studies encompassed all the components of good work identified in *Fair Society, Healthy Lives* (Marmot et al, 2010). However, the definitions we considered complex did consider elements of precariousness, worker control and the appropriateness of the demands placed on the worker.

### Quality of included studies

Overall the quality of the included studies was moderate (Table 2). We found no studies that we considered to have good external validity or generalisability to the United Kingdom (UK). For some this was because of likely differences in the labour market between the UK and the countries where the studies were conducted. For UK studies, and some conducted in other countries, because of poor response rates and attrition from longitudinal studies, we had concerns about how representative the sample was likely to be of the working population in the UK.

We considered only two studies had good internal validity, these fulfilled all or most of the checklist items (rated IV good ++/EV moderate +); one a cohort study (Butterworth et al, 2011), the other case control (Gebel and Vosemer, 2014). Eleven studies were of moderate quality and fulfilled some of the checklist criteria (rated IV good +/ EV good +); seven were cross sectional and four cohort. A further five studies met some of the checklist criteria for internal validity but few or none for external validity (rated IV moderate +/ EV poor -); two were cohort studies and three cross sectional. We rated seven studies as poor. These were six cross sectional studies and one cohort that fulfilled few or none of the checklist criteria (rated IV poor -/ EV poor -).

The design of the included studies meant that in most cases it was not possible to know the direction of the relationship between exposure and outcome. This is inherent in cross sectional studies where data on both exposure and outcome are collected at the same time point. Cohort studies can assess the impact of transitions in employment status. However, data on outcome and exposure in the cohort studies we identified were collected annually. Therefore, it was not possible to know whether the change in exposure – transition in employment status – led to the outcome, or if a change in the outcome (for example improvement or deterioration in participants’ mental health status) resulted in a change in their labour market status. This is a major weakness in the evidence base.

### Findings

There was considerable heterogeneity across studies in terms of design, population, definition of poor job and outcome types and measures so quantitative meta-analysis could not be undertaken. Where we calculated an OR or RR we found our interpretation of the results comparable to those in the paper.

Table 3 provides a summary of results from the studies included in our synthesis. Table S1 in the supplementary material includes the relevant results from all the studies we included. Many of the studies we included reported findings on more than one outcome or relationship.

**Table 3:**
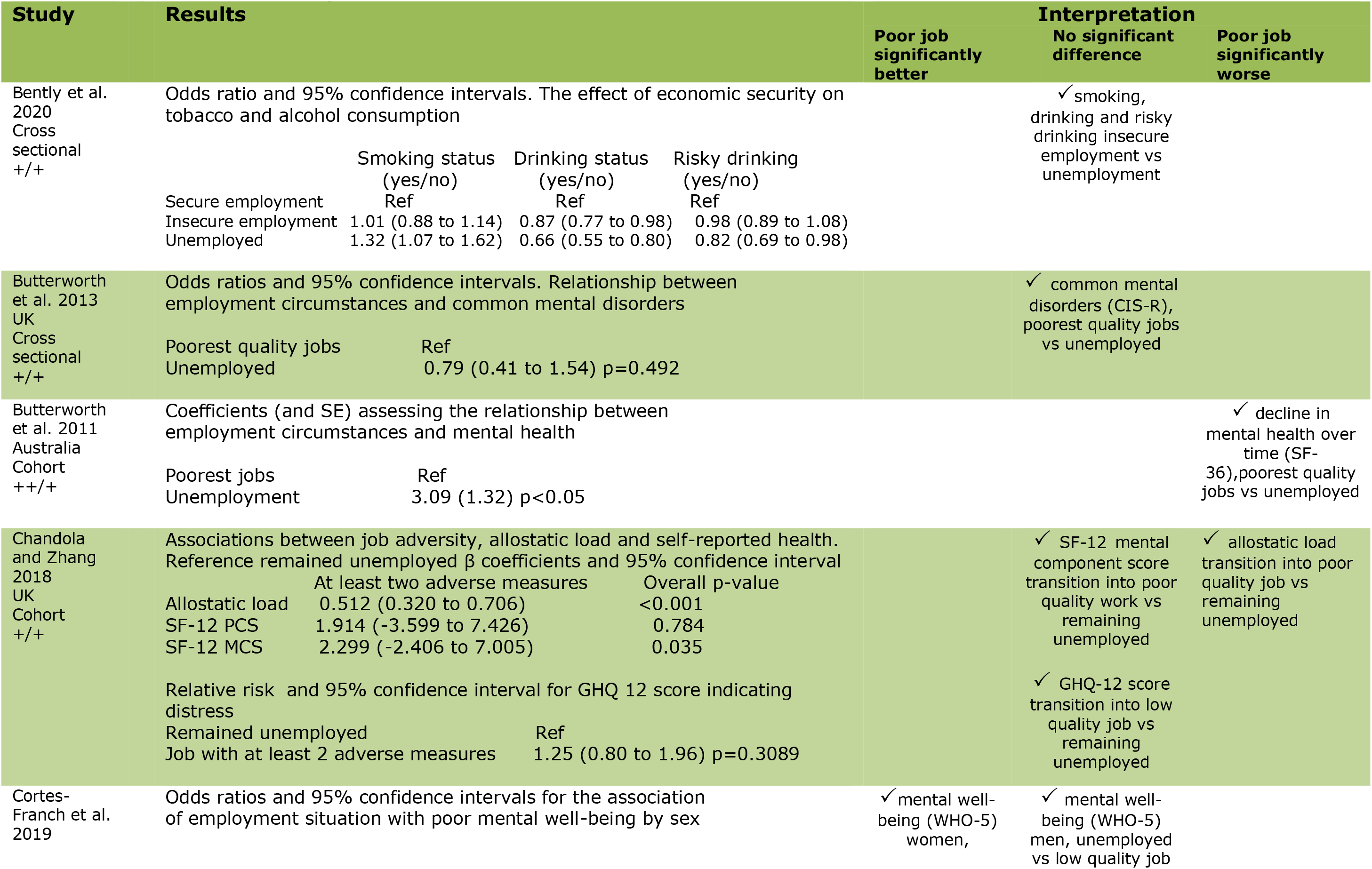
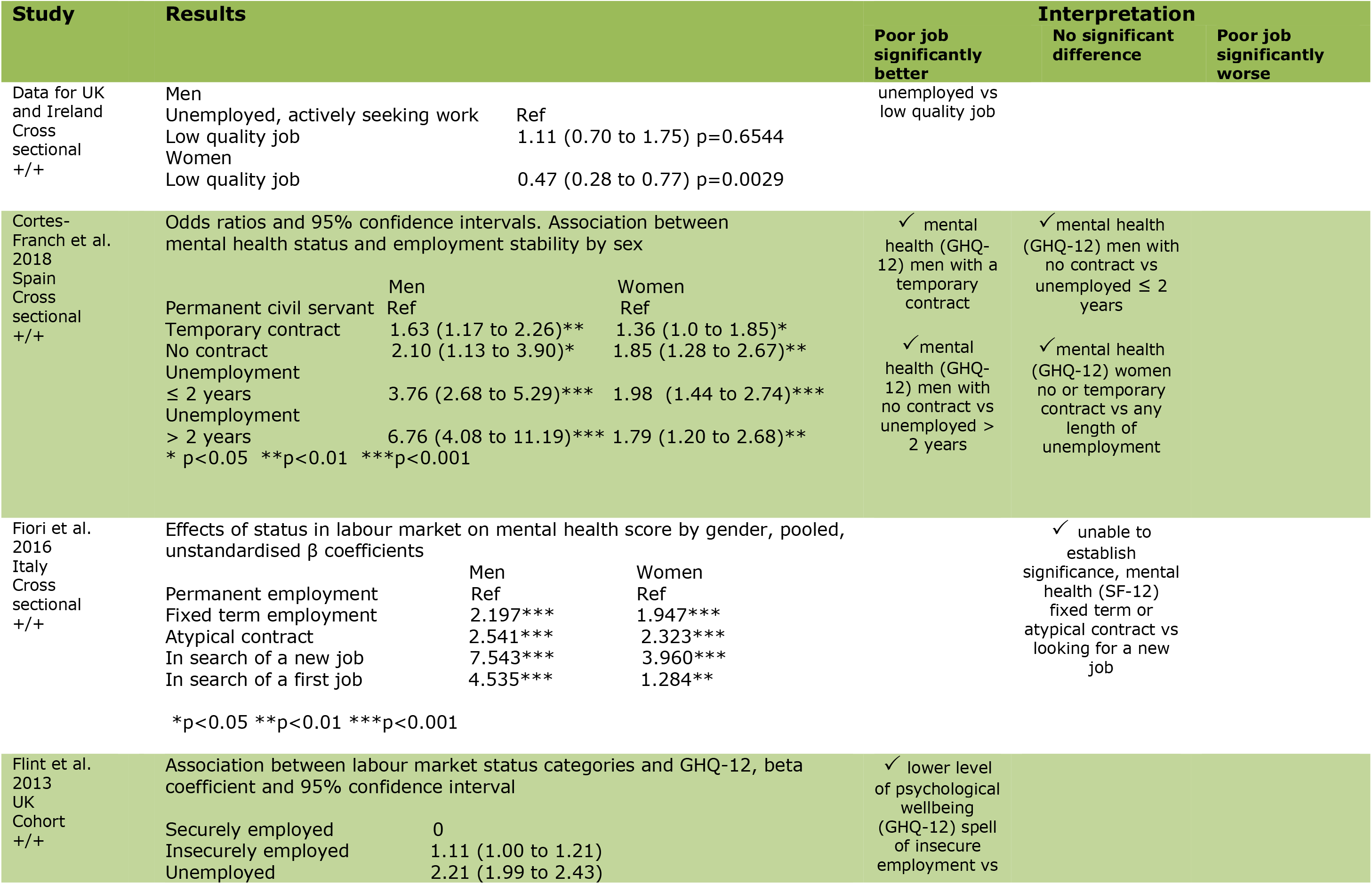
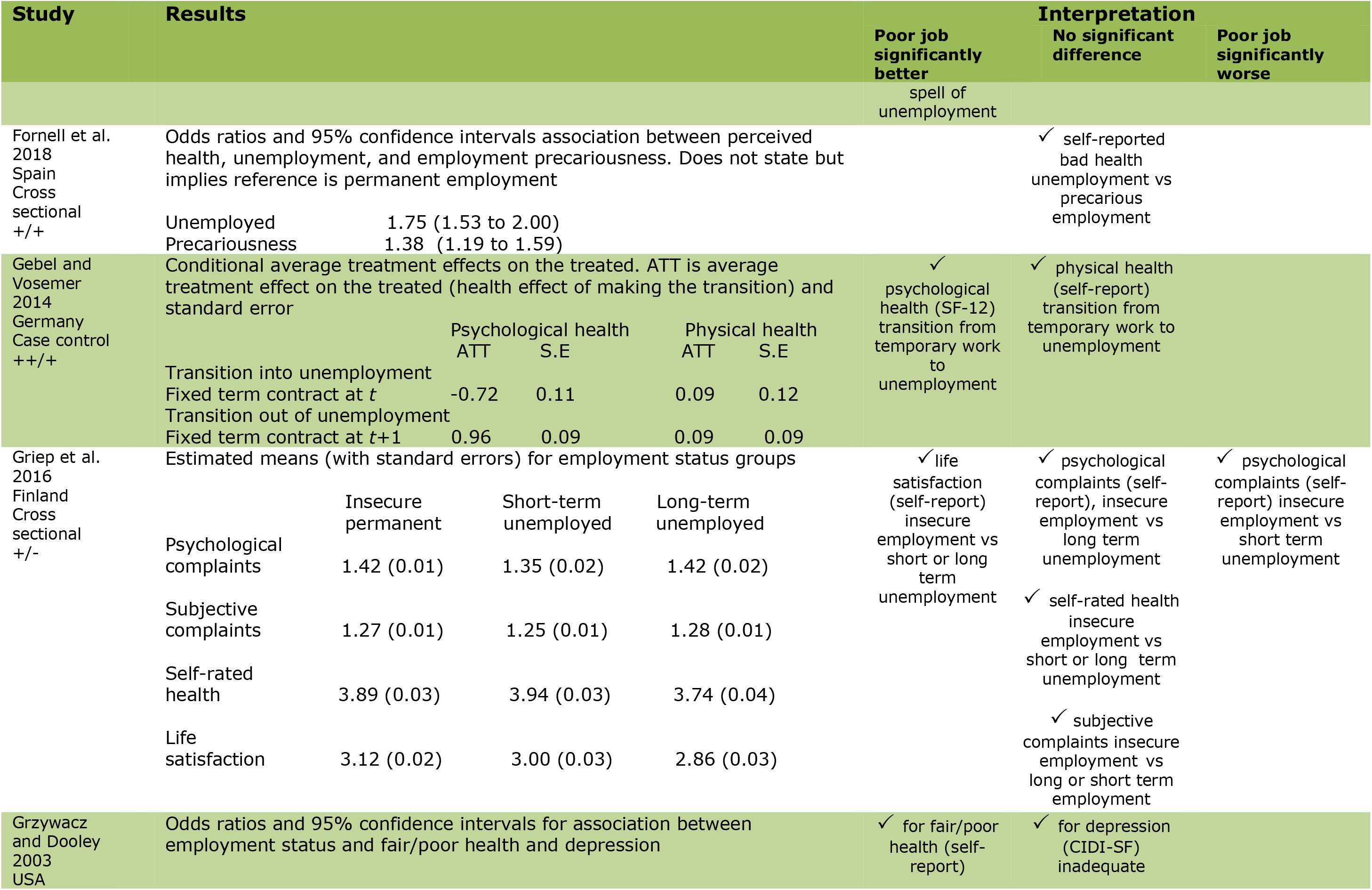
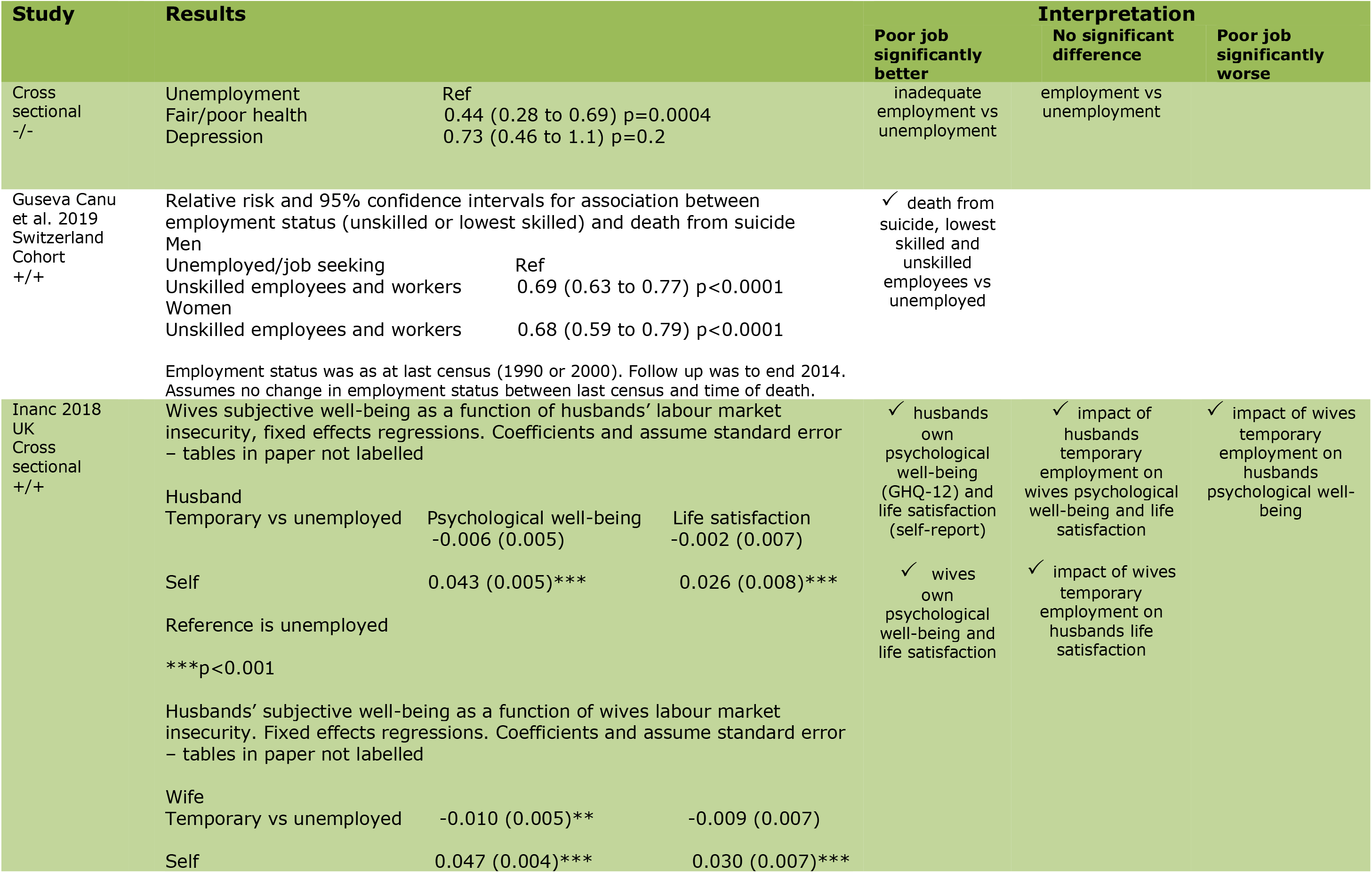
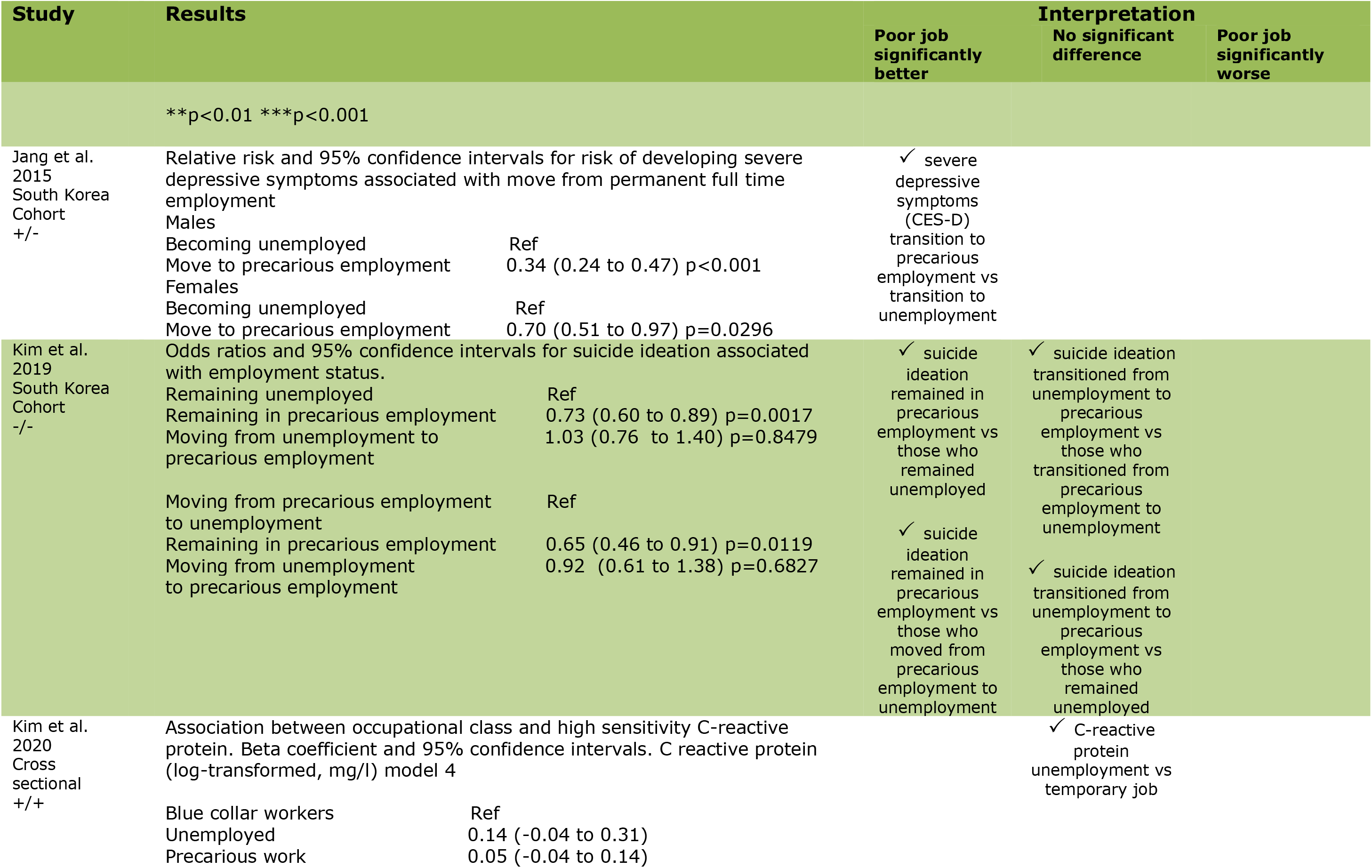
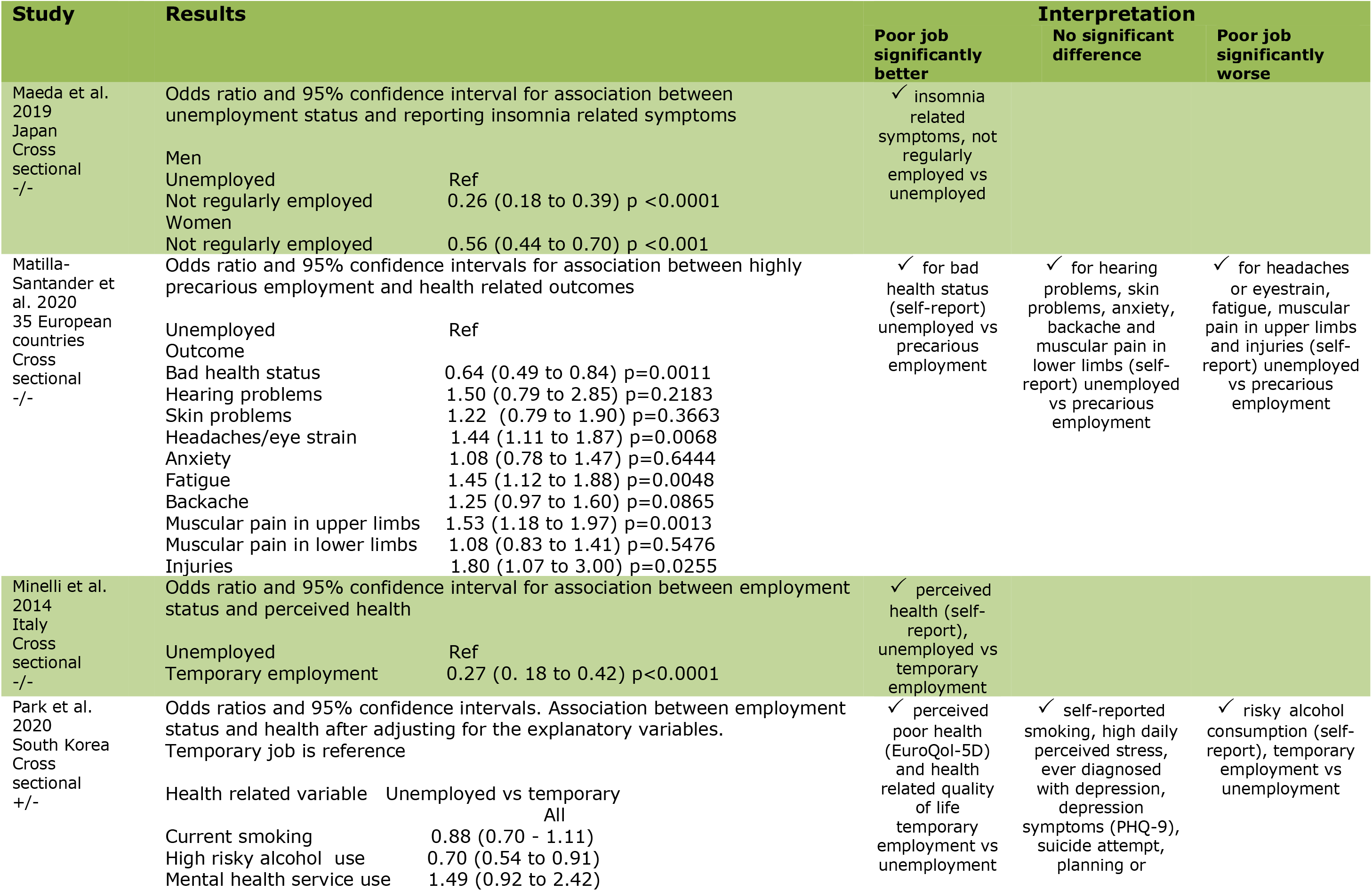
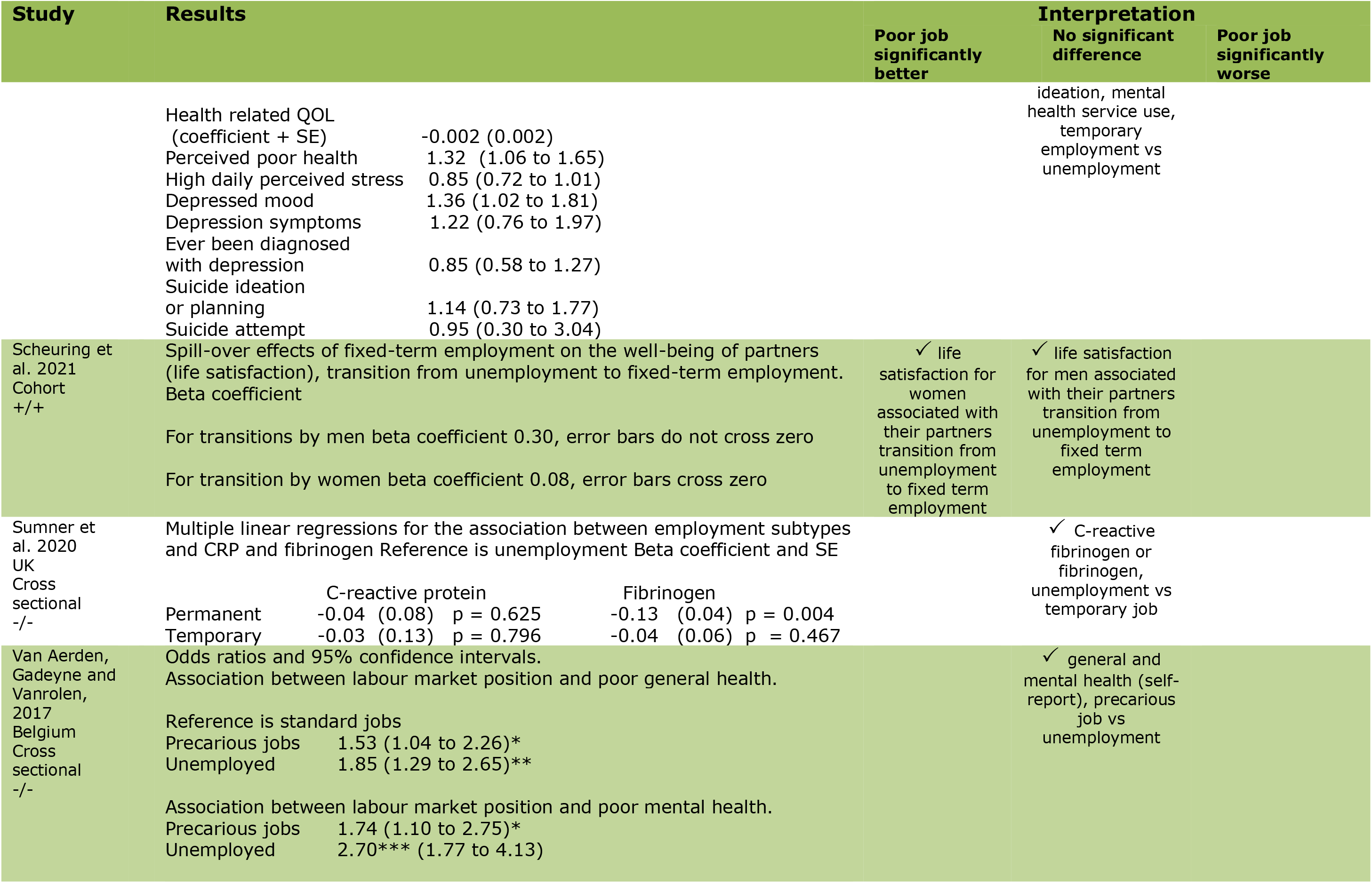
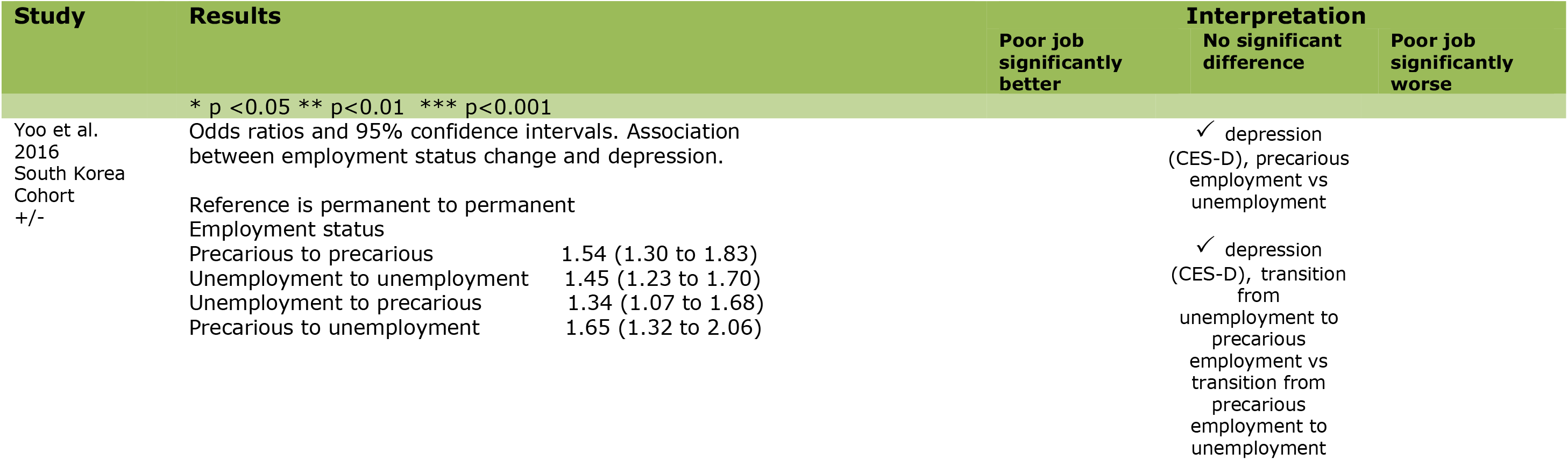
Results used in the synthesis.

We found no consistent direction of evidence (Table 3). However, five studies reported a significant association suggesting that a poor job was significantly better than unemployment (Flint et al, 2013; Guseva Canu et al, 2019; Jang et al, 2015; Maeda et al, 2019 and Minelli et al, 2014). One further study also suggested this, but it was not possible to establish if this result was significant (Fiori et al, 2016).

One study suggested that a poor job was significantly worse than unemployment (Butterworth et al, 2011). Another found that allostatic load (health and chronic stress related bio-markers) was significantly higher in those with a poor job than in those who were unemployed but no difference between these groups for mental component score (Chandola and Zhang, 2018). A further seven studies showed no significant difference (Bently et al, 2020; Butterworth et al, 2012; Fornell et al, 2018; Kim et al, 2020; Sumner et al, 2020; Van Aerden, Gadeyne and Vanrolen, 2017 and Yoo et al, 2016).

One study reported a significant association favouring poor job for an individuals’ own psychological well-being and life satisfaction; however, findings were inconsistent for impact on their partners (Inanc, 2018). A similar study (Scheuring et al, 2021) reported a significant improvement in a women’s life satisfaction if her male partner moved from unemployment to a poor job but no significant difference for men if their female partner made the same transition. One study found a non-significant difference between poor job and unemployment in men but a significant result favouring poor job in women (Cortes-Franch et al, 2019). In another study with the same lead author (Cortes-Franch et al, 2018) the result was significant, favouring a poor job for men but was non-significant for women. The remaining six studies had mixed results that varied by outcome measure or type of employment transition

We explored a range of study variables to see if a clear direction of effect emerged.

### Study design

Looking at the results by study design (Table 3) suggests that the more robust designs (cohort and case control studies) are more likely to find that a poor job is associated with better outcomes than unemployment. Given that cohort or case control studies might support conclusions about causation where there are sufficient numbers of well-designed studies showing a strong relationship, we explored these designs further (Table 4).

Three of the nine studies reported results showing that a poor job was significantly better than no job. Flint et al (2013) identified elevated GHQ- 12 score among the insecurely employed (B 1.11, 95% CI 1.00 to 1.21) and unemployed (B 2.21, 95% CI 1.99 to 2.43). Guseva Canu et al (2019) found death from suicide was reduced among those in a poor job (men RR 0.69, 95% CI 0.63 to 0.77; women RR 0.68, 95% CI 0.59 to 0.79). Finally, Jang et al (2015) found reduced depressive symptoms among men (RR 0.34, 95% CI 0.24 to 0.47) and women (RR 0.70, 95% CI 0.51 to 0.97).

One study (Butterworth et al, 2011) identified a significant result in the opposite direction. This found a greater decline in mental health among those in the poorest quality jobs compared with those who were unemployed (coefficient 3.09, SE 1.32, p<0.05). A further study (Chandola and Zhang, 2018) identified significantly higher allostatic load in those who transitioned into poor quality jobs vs those remaining unemployed (β coefficient 0.512, 95% CI 0.32 to 0.71). However, results for other outcomes reported in this study were not significant (GHQ score indicating distress RR 1.25, 95% CI 0.80 to 1.96). One further study looking at employment status changes and depression (Yoo et al, 2016, reference remaining in permanent employment) found no significant differences between those remaining in precarious employment (OR 1.54, 95% CI 1.30 to 1.83) those remaining unemployed (OR 1.45, 95% CI 1.23 to 1.70), those moving from unemployment to precarious employment (OR 1.45, 95% CI 1.23 to 1.70), or those moving from unemployment to precarious employment (OR 1.34, 95% CI 1.07 to 1.68).

The three remaining studies had mixed results. Gebel and Vosemer (2014) reported significant psychological benefits of being in fixed term employment compared with unemployment for psychological health (average treatment for transition from fixed term contract into unemployment (-0.72, SE 0.11); transition from unemployment to fixed term contract (0.96, SE 0.09). But no significant difference for physical health (average treatment effect transition from fixed term contract to unemployment (0.09, SE 0.12); transition from unemployment to fixed term contract (0.09, SE 0.09). Kim et al. (2019) looked at suicide ideation (using a reference of remaining in permanent employment) for remaining in precarious employment (OR 1.86, 95% CI 1.21 to 2.85), for remaining in unemployment (OR 1.57, 95% CI 0.86 to 2.87), for transition from precarious employment to unemployment (OR 1.43, 95% CI 1.05 to 1.95) and for transition from unemployment to precarious employment (OR 1.11, 95% CI 0.49 to 2.50). Scheuring et al (2021) looked at the impact on life satisfaction of a partner’s transition from unemployment to fixed-term employment in heterosexual couples. For men the impact of their partners transition was not significant (β coefficient 0.08, no confidence intervals but error bars crossed zero). For woman their partner’s transition was associated with a significant improvement in life satisfaction (β coefficient 0.30, no confidence intervals but error bars did not cross zero)

### Study quality

Investigating the results by study quality (Table 3) does not suggest any consistent pattern in outcomes between a poor job and no job. The studies we rated as having the highest internal validity show one significant result favouring unemployment (Butterworth et al, 2011) and one with mixed findings (Gebel and Vosemer, 2014). These also differed by outcome. One study we rated as moderate quality (Chandola and Zhang, 2018) reported a positive finding favouring unemployment for allostatic load but non-significant results for other outcomes.

We rated a further ten studies as moderate quality including three finding no significant difference in the outcomes. Bently et al (2020) found no significant differences in smoking status between insecure employment (OR 1.01 95% CI 0.88 to 1.14) and unemployment (OR 1.32 (95% CI 1.07 to 1.62 1.01); drinking status (insecure employment OR 0.87 95% CI 0.77 to 0.98; unemployment OR 0.66, 95% CI 0.55 to 0.80) or risky drinking (insecure employment OR 0.98, 95% CI 0.89 to 1.08; unemployment; reference is secure employment).

Butterworth et al (2013) looked at common mental disorders among unemployed (OR 0.79, 95% CI 0.41 to 1.54; reference is poorest quality jobs). Fornell et al (2018) reported on bad health among unemployed (OR 1.75, 95% CI 1.53 to 2.00) and precarious employment (OR 1.38, 95% CI 1.19 to 1.59; assume reference is permanent employment as not stated in paper). Four studies rated as moderate reported mixed results; Cortes- Franch et al (2019) for poor mental well-being (unemployed males OR 1.1, 95% CI 0.70 to 1.75; unemployed females OR 0.47, 95% CI 0.28 to 0.77, reference is low quality job); Cortes-Franch et al (2018) for mental health status (males; temporary contract OR 1.63, 95% CI 1.17 to 2.26; no contract OR 2.10, 95% CI 1.13 to 3.90; unemployment 2 years or less OR 3.76, 95% CI 2.68 to 5.29; unemployment more than two years OR 6.76, 95% CI 4.08 to 11.9; females; temporary contract OR 1.36, 95% CI 1.0 to 1.85; no contract OR 1.85, 95% CI 1.28 to 2.67; unemployment two years or less OR 1.98, 95% CI 1.44 to 2.74; unemployment more than two years OR 1.79, 95% CI 1.20 to 2.68; reference is permanent civil servant) and Inanc, 2018 (coefficient for negative impact psychological well-being; for men, wife temporary vs unemployed -0.010 SE 0.005, self 0.043 SE 0.005; for women, husband temporary vs unemployed -0.006 SE 0.005, self 0.047 SE 0.004). Scheuring et al (2021) reported a significant impact on women’s life satisfaction associated with their partner’s transition from unemployment to fixed-term employment but no significant difference for men.

Three studies also rated as moderate quality favoured a poor job; Fiori et al, 2016 (unstandardised β coefficients mental health score; men; fixed term employment 2.197, atypical contract 2.541; in search of new job 7.543; women; fixed term employment 1.947, atypical contract 2.323, in search of new job 3.960; reference is permanent employment) Flint et al. (2013) identified elevated GHQ-12 score among insecurely employed (B 1.11, 95% CI 1.00 to 1.21) and unemployed (B 2.21, 95% CI 1.99 to 2.43).

Guseva Canu et al. (2019) found death from suicide was reduced among those in a poor job (men RR 0.69, 95% CI 0.63 to 0.77; women RR 0.68, 95% CI 0.59 to 0.79)

There were seven studies that we considered weak. Of these, one (Van Aerden, Gadeyne and Vanrolen, 2017), found no significant difference in outcomes. The study reported on poor general health among those in precarious jobs (OR 1.53, 95% CI 1.04 to 2.26), unemployed (OR 1.85, 95% CI 1.29 to 2.65); and poor mental health among those in precarious jobs (OR 1.74, 95% CI 1.10 to 2.75), unemployed (OR 2.70, 95% CI 1.77 to 4.13; reference is standard jobs).

Five of the six remaining studies we considered weak found at least one significant result. Of these, three had mixed findings depending on outcome (Grzywacz and Dooley, 2003; Matilla-Santander et al, 2020) or type of employment transition (Kim et al, 2019). Two studies favoured poor jobs (Maeda et al, 2019; Minelli et al, 2014).The final study found no significant difference (Sumner et al, 2020).

### Outcome and outcome measure (type and objectivity)

Comparison by type of outcome was difficult because of the range of tools used but shows no consistent pattern of results (Table 3). We also looked at the objectivity of the outcome measures used (Table S2, supplementary material). However, this also suggests no consistent pattern. Of the studies reporting objective outcome measures, three studies used biomarkers. Two (Kim et al, 2020; Sumner et al, 2020) looked at markers of peripheral inflammation and reported non-significant findings (β coefficient and 95% CI for unemployed 0.14, -0.04 to 0.31, precarious work 0.05, -0.04 to 0.14, reference is blue collar workers; β coefficient and SE for temporary employment C-reactive protein -0.03 SE 0.13, p=0.796; fibrinogen -0.04 SE 0.06, p=0.467, reference is unemployment). The third study, Chandola and Zhang (2018), used allostatic load, markers related to chronic stress reported a significant outcome favouring unemployment. Data from one further study (Guseva Canu et al. 2019) using an objective outcome, death through suicide, showed a significant result favouring poor job (men RR 0.69, 95% CI 0.63 to 0.77; women RR 0.68, 95% CI 0.59 to 0.79).

### Mental health outcomes

Mental health outcomes were the most frequently reported in the included studies. Eighteen looked at these (Table 5). Overall the results suggest no consistent pattern. Seven studies reported results finding no significant difference (Butterworth et al, 2013; Chandola and Zhang 2018; Fiori et al, 2016; Grzywacz and Dooley, 2003; Matilla-Santander et al, 2020; Van Aerden, Gadeyne and Vanrolen, 2017; Yoo et al, 2016).

Four studies found that a poor job was significantly better than unemployment (Flint et al, 2013; Gebel and Vosemer 2014; Guseva Canu et al, 2019; Jang et al; 2015). One study reported that a poor job was significantly worse (Butterworth et al, 2011).

The remaining seven studies reported mixed results. Cortes-Franch et al (2019) found that mental well-being was significantly better in women with a poor job but no significant difference for men. Cortes-Franch et al (2018) had results that differed by sex, contract type and length of unemployment. The results reported by Griep et al (2016) also differed by length of unemployment. They found no significant difference for long term unemployment, but a poor job was significantly worse than short term unemployment. Inanc (2018) looked at the impacts on spouses’ employment as well as that on self. Findings indicated that a poor job was significantly better than unemployment for a husband’s and wife’s own psychological well-being, but there was no difference on a wife’s psychological well-being if her husband was in temporary employment. By contrast, a husband’s psychological well-being was significantly worse if his wife was in temporary employment rather than unemployed.

Scheuring et al (2021) found a significant improvement in life satisfaction for women associated with their male partner’s transition from unemployment to fixed term employment but no significant difference for men associated with the same transition by their female partners. Kim et al (2019) used suicide ideation as an outcome and looked at employment transitions. This longitudinal study found that a poor job was significantly better, comparing those who remained in precarious employment with those who remained unemployed or who moved from precarious employment to unemployment. There was no significant difference between those who moved from unemployment to being precariously employed compared with those whose transition was the other way, or those who moved from unemployment to precarious employment when compared with those who remained unemployed. Finally Park et al (2020) found no significant difference for a range of mental health related outcomes, although a poor job was significantly worse for risky alcohol consumption.

Whilst overall the results for mental well-being are inconsistent, the cohort and case-control studies do show more support for any job being preferable to unemployment. However, the evidence is not strong. Of eight studies, five find that a poor job is significantly better than no job for at least one outcome (Flint et al, 2013; Gebel and Vosemer, 2014; Guseva Canu et al, 2019; Jang et al, 2015; Kim et al, 2019). Of these one reports a strong relationship between exposure and outcome (Flint et al, 2013). Jang et al (2015) reported a moderate to strong relationship for men only. Both studies used a simple definition of a poor job and were rated as being of moderate internal validity. The study by Butterworth et al (2011), which we rated as having good internal validity and which used a complex definition of a poor job, found those in poor jobs reported a significantly worse decline in their mental health than those who were unemployed. One other study (Chandola and Zhang, 2018) also used a complex definition of a poor job and this found no significant difference.

### Definition of poor job

As described earlier, we grouped the definitions of poor job as complex if they were multidimensional and simple where they were not (Table 2). Again results are mixed but suggest a trend toward no difference between poor jobs and unemployment.

Seven studies used a more complex definition of poor job. Of these Butterworth et al (2011) reported a significant result favouring unemployment. Chandola and Zhang (2018) also found a significant result favouring unemployment job for allostatic load but the findings for the other outcomes this study reported were not significant. Butterworth et al (2013) and Van Aerden, Gadeyne and Vanrolen (2017) also reported non-significant findings. The remaining studies had mixed results. For women, Cortes-Franch et al (2019) found a significant result favouring poor job, but for men this was not significant. The findings reported by Grzywacz and Dooley (2003) differed by outcome with data showing a significant result favouring poor job for self-reported health (OR 0.44, 95% CI 0.28 to 0.69) but not significant for depression (OR 0.73, 95% CI 0.46 to 1.19).

The final study (Matilla-Santander et al, 2020) also had results differing by outcome. Data from this showed a significant result favouring poor job for the likelihood of reporting bad health (OR 0.64, 95% CI 0.49 to 0.84). Non-significant results were found for hearing problems (OR 1.50, 95% CI 0.79 to 2.85), skin problems (OR 1.22, 95% CI 0.79 to 1.90), anxiety (OR 1.08, 95% CI 0.78 to 1.47), backache (OR 1.25, 95% CI 0.97 to 1.60) and muscular pain in lower limbs (OR 1.08, 95% CI 0.83 to 1.41). Significant results, favouring unemployment, were found for headaches/eyestrain (OR 1.44, 95% CI 1.11 to 1.87), fatigue (OR 1.45, 95% CI 1.12 to 1.88), muscular pain in upper limbs (OR 1.53, 95% CI 1.18 to 1.97) and injuries (OR 1.80, 95% CI 1.07 to 3.00).

### Employment transitions

Six studies (Table 6) used longitudinal data to examine the impact on health of employment transitions. Comparison is difficult because most are looking at slightly differing transitions. Overall results are mixed, however, two studies looked at the transition from a poor job to unemployment, both finding that a poor job was significantly better than unemployment for mental health outcomes (Gebel and Vosemer, 2014; Kim et al, 2019). Kim et al. (2019) also explored the impact of transition in the other direction as did Chandola and Zhang (2018) but both found no significant difference when compared with the impact of remaining unemployed. Scheuring et al (2021) report impact on partners’ life satisfaction associated with a transition from unemployment to fixed-term work but did not report findings for the partner making the transition. These findings suggest that moving to a poor job from unemployment was not associated with improved mental health but moving from a poor job to unemployment was associated with a deterioration.

### Welfare state regime

We also considered the impact of welfare state regime for European Countries (Table S3 supplementary material). We categorised these based on the typology used by Cortes-Franch et al (2019). The number of studies in each group outside the UK are small and do not suggest a consistent pattern of results.

Of the six studies conducted in or including the UK, four are cross sectional. One cohort reported significant results favouring a poor job (Flint et al. 2013). The other found that for allostatic load unemployment was significantly better than a poor job but findings for mental well-being related outcomes were not significant (Chandola and Zhang, 2018). Two further studies reported non-significant results (Butterworth et al, 2013; Sumner et al, 2020). The remaining two findings were mixed. Cortes-Franch et al (2019) reported a significant result favouring a poor job for women but not men. Finally Inanc (2018) reported significant results favouring a poor job for men and women, a non-significant finding for the impact of man’s employment on his wife, as well as the impact of a woman’s temporary employment on her husband’s life satisfaction, but a significant effect favouring unemployment for the impact of a woman’s temporary employment on her husband’s psychological well-being. Although the results from these six studies are inconsistent, overall they suggest that in the UK bad/poor employment is not significantly associated with better health and well-being outcomes than unemployment.

### Other variables

We also looked at gender, the years for which data were collected and study sample sizes for patterns that might identify if a poor job might be better than no job. These did not appear to have any influence on the pattern of results.

## Discussion

The question “Is any job better than no job” appears simple and is policy relevant, but it has no simple answer. After investigating a range of variables, the evidence on whether having a poor or bad job, however defined, is associated with better health and well-being outcomes than being unemployed is inconsistent. Five studies reported findings suggesting that a poor job was significantly better than unemployment (Flint et al, 2013; Guseva Canu et al, 2019; Jang et al, 2015; Maeda et al, 2019; Minelli et al, 2014). However, seven found no significant difference (Bently et al, 2020; Butterworth et al, 2013; Fornell et al, 2018; Sumner et al, 2020; Van Aerden, Gadeyne and Vanrolen, 2017; Kim at al, 2020; Yoo et al. 2016). One study found that a poor job was associated with significantly worse outcomes (Butterworth et al, 2011) and there was one study where we could not establish significance (Fiori et al. 2016). The remaining eleven studies reported mixed results.

A recent study, looking at country level associations (so not meeting our eligibility criteria), also suggests that a poor job may not be associated with better well-being outcomes than unemployment. Scheuring (2020) explored whether fixed-term employees had greater subjective well-being than the unemployed. Using 2012 data on 23 countries from the European Social Survey this study found that for 18 countries those in fixed-term employment reported greater subjective well-being than the unemployed. However, this difference in well-being only appears to be statistically significant in five of those 18 countries.

Despite this inconsistency some potentially useful findings do emerge. Studies using cohort or case-control designs seem more likely to find that a poor job is associated with better outcomes than unemployment. This is particularly the case for outcomes related to mental well-being. Another interesting finding from longitudinal studies is that, whilst transition to a poor job from unemployment is not associated with an improvement in mental health, moving from a poor job to unemployment is associated with a deterioration. This finding is from only three studies but would be worth further exploration.

Studies conducted in the UK suggest that a poor job is not significantly associated with better health and well-being outcomes than unemployment Although a number find that a poor job is better than no job, others find the difference is not significant and there is no clear majority of studies either direction. This might reflect the type of welfare state regime protecting, to some extent, those who are unemployed from health and well-being harms; equally it may reflect job quality, suggesting that poor or bad employment is as damaging to health and well-being as unemployment. However, overall the evidence for this is inconsistent.

Notwithstanding the inconsistent findings, the evidence base we identified is largely too weak to draw any firm conclusion about the relationship between employment status and health and well-being outcomes. Quantitative meta-analysis would have been helpful but studies were too heterogeneous in terms of design, included population, definition of poor job and outcome measures to consider this.

Observational studies (cohort or case control) can sometimes support conclusions about causation, but only when there are sufficient numbers of well-designed studies showing a strong relationship. It is not possible from the evidence base we identified to establish the direction of the relationship between outcome and employment status. All the included studies involved secondary analysis of data from national cohorts with data on both exposure and outcome collected annually. Only one, a case control study, (Gebel and Vosemer, 2014) collected additional data on a monthly basis. This found a significant result for psychological health favouring poor jobs, but the result for physical health was not significant. We found no study that collected data with the primary purpose of exploring the relationship between employment status and health and well-being outcomes. A further consideration is that we cannot establish the temporal sequence between exposure and outcome from our included studies. It is not possible to tell if change in employment status preceded change in outcome, or vice versa.

Risk of bias was an issue in our included studies. Study quality was generally moderate or poor and we found no studies with good external validity. Five of the six that we rated as weak for both internal and external validity reported at least one significant finding. It is recognised that poor quality studies may tend to find greater effects than those of better quality (Moher et al, 1998). However, the weak studies in our review did not report findings in a consistent direction.

Study samples were a particular issue. Although all studies intended to achieve a nationally representative sample, many had poor response rates, and attrition was a concern in some of the longitudinal studies. Generally the outcome measures were very broad, weak and most relied on self-report. In some studies un-validated measures or questions were used. Kim et al. (2019) used a highly subjective question to establish suicide ideation by asking if the respondent had any thoughts of suicide in the past year, despite there being validated measures that could have been used (Harmer et al, 2020). Conversely, Guseva Canu et al (2019) used death certificates to establish cause of death. As we identified, separating the studies into those using validated and non-validated or self-report measures, still failed to establish any relationship between the exposure and outcomes. Although some studies found apparently large effect sizes the outcomes and outcome measures used may have implications for how useful these might be at population level for discriminating between the impact of a poor job and unemployment. The study by Jang et al (2015) reports a relative risk suggesting a substantial reduction for risk of severe depression for those in precarious employment compared to those who were unemployed (RR 0.34, 95% CI 0.24 to 0.47). However, this relies on use of a questionnaire with a cut-off point indicating severe depression rather than clinical assessment by an appropriately trained professional able to diagnose whether or not an individual has severe depression.

Most of the studies we included used simple definitions of a poor job (for example temporary or insecure employment) often without any further explanation or definition. Seven of the 22 (32%) studies we included in the synthesis used a multi-dimensional approach to job quality. In their systematic review Utzet at al. (2020) found a similar proportion, 10 of 32 (31%). Utzet and colleagues found that most of the studies looking at job insecurity, temporariness and those using multi-dimensional approaches reported a significant association with mental health problems. However, their comparator was not unemployment so their findings are not directly comparable with our review.

We did not find any studies that attempted anything approximating to the comprehensive definition of good work used in Fair Society, Healthy Lives (Marmot, 2010). We considered whether this might be related to our decision to include only studies that were based on a sample intended to be representative of the economically active national population. We made this decision to maximise the likelihood that the studies we included would generalise to the UK setting. This meant that all the studies we included involved secondary analysis of data derived from national cohort studies. The primary purpose of these national cohorts was not to explore the relationship between employment status and health and well-being outcomes. This meant that data on occupational status in our included studies was primarily collected as an indicator of socio-economic status rather than as a measure of type of employment. This may offer some explanation for the studies’ lack of multidimensional definitions of type of employment.

We excluded 13 studies at full text because they did not use a national sampling frame. (Further details of excluded studies are in the supplementary material). We wondered if this had led us to exclude studies where the primary purpose was to explore the impact of quality or type of employment. Further consideration of these 13 studies found only one where the primary purpose was to look at the relationship between labour market status and health. This was part of a Spanish study, Immigration, Labour and Health (ITSAL), looking at the employment and working conditions of immigrant workers and their relationship to health (Robert at al. 2014). We excluded this because of the study population and lack of a poor job vs unemployed comparison. On this basis we think it is unlikely that we would have excluded studies where the primary purpose was to explore the relationship between quality of employment (or unemployment) and health and well-being.

Other systematic reviews report inconsistent evidence on the relationship between the quality of employment and health related outcomes. Amiri and Behnezhad (2020) found an increased risk of mortality associated with job strain in European but not in American studies. It is not clear what the reference group is but the assumption is those without job strain rather than the unemployed. They also found a significant relationship in men but not women. Broadly in line with our findings Kim and von dem Knesebeck (2015) looked at health related risks associated with job insecurity and unemployment. They also found inconsistent evidence reporting a strong association between mental health and both exposures with no clear direction of effect.

As noted by Rönnblad and colleagues (2019) there is a lack of high quality prospective studies with policy relevant results. Systematic reviews often conclude calling for more and better research. Employment is an important determinant of health and there is a need for a better understanding of the characteristics of work that have benefit for health. This needs prospective longitudinal studies where the primary focus is on the relationship between labour market status and health and well-being outcomes. Such studies would need to adopt a multi-dimensional definition of poor work, use objective and reliable outcome measures and collect data frequently so that the direction of the relationship between work and health might be established. Studies of this type could address many of limitations that we have identified in the studies included in this review.

Our systematic review has a number of limitations. We may not have identified all relevant studies because we did not contact authors working in this field, search the content lists of relevant journals or undertake citation tracking of the studies we included. We limited included studies to those with an abstract in English. This led to the exclusion of four studies published in German that we had included at abstract but could not translate to assess at full text. Publication bias was not assessed, but findings in either direction are likely to be of equal interest.

## Conclusion

The current evidence base available to answer the question “Is any job better than no job” is characterised by studies that differ quite widely in the relationships they consider, the type and robustness of the outcomes they measure and the definitions they use. There are also limitations in study design and conduct. Despite this inconsistency and limitations, some potential useful findings do emerge. Studies using cohort or case-control designs, seem more likely to find that a poor job is associated with better outcomes than unemployment. This is the case particularly for outcomes related to mental well-being. A small number of studies looking at employment transitions find that moving to a poor job from unemployment is not associated with improved mental health but moving from a poor job to unemployment is associated with a deterioration. This finding is from only three studies but would be worth further exploration.

Studies conducted in the UK suggest that a poor job is not significantly associated with better health and well-being outcomes than unemployment. The studies we identified do not allow us to distinguish whether this lack of association is as a result of the state welfare regime preventing some of the worst ills associated with unemployment or a reflection of job quality.

Overall the evidence base is inconsistent. In summary, it is not safe to assume that any job will lead to better health and well-being outcomes than no job, or that increasing employment through poor quality jobs will improve health overall.

## Data Availability

All relevant data are contained in the manuscript and supplementary material

## References

Amiri, S. and Behnezhad, S. (2020) ’Job strain and mortality ratio: a systematic review and meta-analysis of cohort studies’, Public Health, 181: 24–33.

Bentley, R., Baker, E., Martino, E., Li, Y. and Mason, K. (2021) ’Alcohol and tobacco consumption: What is the role of economic security?’, Addiction, 116(7): 1882–91.

Butterworth, P., Leach, L. S., McManus, S.and Stansfeld, S. A. (2013) ’Common mental disorders, unemployment and psychosocial job quality: is a poor job better than no job at all?’, Psychological Medicine, 43(8): 1763–72.

Butterworth, P., Leach, L. S., Strazdins, L., Olesen, S. C., Rodgers, B. and Broom, D. H. (2011) ’The psychosocial quality of work determines whether employment has benefits for mental health: results from a longitudinal national household panel survey’, Occupational and Environmental Medicine, 68(11): 806.

Carré, F., Findlay, P., Tilly, C. and Warhurst, C. (2012) ’Job Quality: Scenarios, Analysis and Intervention’, in Warhurst, C., Carré, F., Findlay, P. and Tilly, C. (eds), Are bad jobs inevitable?: trends, determinants and responses to job quality in the twenty-first century, Basingstoke, Palgrave McMillan.

Chandola, T. and Zhang, N. (2018) ’Re-employment, job quality, health and allostatic load biomarkers: prospective evidence from the UK Household Longitudinal Study’, International Journal of Epidemiology, 47(1): 47–57.

Cortès-Franch, I., Escribà-Agüir, V., Benach, J. and Artazcoz, L. (2018) ’Employment stability and mental health in Spain: towards understanding the influence of gender and partner/marital status’, BMC Public Health, 18(1): 425.

Cortes-Franch, I., Puig-Barrachina, V., Vargas-Leguas, H., Arcas, M. M. and Artazcoz, L. (2019) ’Is being employed always better for mental wellbeing than being unemployed? Exploring the role of gender and welfare state regimes during the Economic Crisis’, International Journal of Environmental Research & Public Health [Electronic Resource], 16(23): 29.

Crossley, T. F., Fisher, P., Levell, P. and Low, H. (2021) A year of COVID: the evolution of labour market and financial inequalities through the crisis. Working Paper 21/39, London: Institute for Fiscal Studies.

Davies, G. (2021) Addressing skills and labour shortages post-Brexit, London: Chartered Institute of Personnel and Development.

Fair Work Commission (2019) Fair Work Wales. Report of the Fair Work Commission, Cardiff: Fair Work Commission.

Fair Work Convention (2016) Fair Work Framework, Edinburgh: Fair Work Convention.

Fiori, F., Rinesi, F., Spizzichino, D. and Di Giorgio, G. (2016) ’Employment insecurity and mental health during the economic recession: An analysis of the young adult labour force in Italy’, Social Science & Medicine, 153: 90–8.

Flint, E., Bartley, M., Shelton, N. and Sacker, A. (2013) ’Do labour market status transitions predict changes in psychological well-being?’, Journal of Epidemiology and Community Health, 67(9): 796.

Fornell, B., Correa, M., López del Amo, M. P. and Martín, J. J. (2018) ’Influence of changes in the Spanish labor market during the economic crisis (2007–2011) on perceived health’, Quality of Life Research, 27(8): 2095–105.

Gebel, M. and Voßemer, J. (2014) ’The impact of employment transitions on health in Germany. A difference-in-differences propensity score matching approach’, Social Science & Medicine, 108: 128–36.

Griep, Y., Kinnunen, U., Natti, J., De Cuyper, N., Mauno, S., Makikangas, A. and De Witte, H. (2016) ’The effects of unemployment and perceived job insecurity: a comparison of their association with psychological and somatic complaints, self-rated health and life satisfaction’, International Archives of Occupational & Environmental Health, 89(1): 147–62.

Grzywacz, J. G. and Dooley, D. (2003) ’“Good jobs” to “bad jobs”: replicated evidence of an employment continuum from two large surveys’, Social Science & Medicine, 56(8): 1749–60.

Guseva Canu, I., Bovio, N., Mediouni, Z., Bochud, M., Wild, P. and Swiss National Cohort. (2019) ’Suicide mortality follow-up of the Swiss National Cohort (1990-2014): sex-specific risk estimates by occupational socio- economic group in working-age population’, Social Psychiatry & Psychiatric Epidemiology, 54(12): 1483–95.

Harmer, B., Lee, S., Duong, T. V. H. and Saadabadi, A. (2021) ’Suicidal Ideation’, StatPearls, Treasure Island (FL), StatPearls PublishingCopyright © 2021, StatPearls Publishing LLC.

Inanc, H. (2016) Unemployment, temporary work and subjective well-being: Gendered effect of spousal labour market insecurity in the United Kingdom. Working Paper 70, Paris: OECD.

Inanc, H. (2018) ’Unemployment, Temporary Work, and Subjective Well-Being: The Gendered Effect of Spousal Labor Market Insecurity’, American Sociological Review, 83(3): 536–66.

International Labor Organization (1999), Decent Work, Report of the Director-General. Proceedings of the 87th Session, International Labor Conference, Geneva, International Labor Organization.

Jackson, R., Ameratunga, S., Broad, J., Connor, J., Lethaby, A., Robb, G., Wells, S., Glasziou, P. and Heneghan, C. (2006) ’The GATE frame: critical appraisal with pictures’, Evidence Based Medicine, 11(2): 35.

Jang, S. Y., Jang, S. I., Bae, H. C., Shin, J. and Park, E. C. (2015) ’Precarious employment and new-onset severe depressive symptoms: a population-based prospective study in South Korea’, Scandinavian Journal of Work, Environment & Health, 41(4): 329–37.

Kim, H. D. and Park, S. G. (2021) ’Employment status change and new-onset depressive symptoms in permanent waged workers’, Safety & Health at Work, 12(1): 108–13.

Kim, I. H., Muntaner, C., Vahid Shahidi, F., Vives, A., Vanroelen, C. and Benach, J. (2012) ’Welfare states, flexible employment, and health: a critical review’, Health Policy, 104(2): 99–127.

Kim, S. S., Muntaner, C., Kim, H., Jeon, C. Y. and Perry, M. J. (2013) ’Gain of employment and depressive symptoms among previously unemployed workers: a longitudinal cohort study in South Korea’, American Journal of Industrial Medicine, 56(10): 1245–50.

Kim, S. S., Subramanian, S., Sorensen, G., Perry, M. J. and Christiani, D. C. (2012) ’Association between change in employment status and new-onset depressive symptoms in South Korea - a gender analysis’, Scandinavian Journal of Work, Environment & Health, 38(6): 537–45.

Kim, T. J. and von dem Knesebeck, O. (2015) ’Is an insecure job better for health than having no job at all? A systematic review of studies investigating the health-related risks of both job insecurity and unemployment’, BMC Public Health, 15(1): 985.

Kim, W., Ki, M., Choi, M. and Song, A. (2019) ’Comparable Risk of Suicidal Ideation between Workers at Precarious Employment and Unemployment: Data from the Korean Welfare Panel Study, 2012-2017’, International Journal of Environmental Research & Public Health [Electronic Resource], 16(16): 07.

Kim, Y., Zaitsu, M., Tsuno, K., Li, X., Lee, S., Jang, S. and Kawachi, I. (2020) ’Occupational Differences in C-Reactive Protein Among Working-Age Adults in South Korea’, Journal of Occupational and Environmental Medicine, 62(3).

Koranyi, I., Jonsson, J., Rönnblad, T., Stockfelt, L. and Bodin, T. (2018) ’Precarious employment and occupational accidents and injuries – a systematic review’, Scandinavian Journal of Work, Environment & Health, 10.5271/sjweh.3720(4): 341–50.

Leaker, D. (2021) Labour market overview, UK: October 2021, Newport: ONS.

Maeda, M., Filomeno, R., Kawata, Y., Sato, T., Maruyama, K., Wada, H., Ikeda, A., Iso, H. and Tanigawa, T. (2019) ’Association between unemployment and insomnia-related symptoms based on the Comprehensive Survey of Living Conditions: a large cross-sectional Japanese population survey’, Industrial Health, 57(6): 701–10.

Marmot, M., Allen, J., Boyce, T., Goldblatt, P. and Morrison, J. (2020) Health equity in England: the Marmot review 10 Years On, London Institute of Health Equity

Marmot, M., Allen, J., Boyce, T., Goldblatt, P. and Morrison, J. (2021) Build Back Fairer in Greater Manchester: Health Equity and dignified lives. London: Institute of Health Equity.

Marmot, M., Allen, J., Goldblatt, P., Boyce, T., McNeish, D., Grady, M. and Geddes, I. (2010) Fair society, healthy lives: the Marmot review; strategic review of health inequalities in England post-2010, London: The Marmot Review.

Matilla-Santander, N., Martín-Sánchez, J. C., González-Marrón, A., Cartanyà-Hueso, À., Lidón-Moyano, C. and Martínez-Sánchez, J. M. (2021) ’Precarious employment, unemployment and their association with health-related outcomes in 35 European countries: a cross-sectional study’, Critical Public Health, 31(4): 404–15.

Minelli, L., Pigini, C., Chiavarini, M. and Bartolucci, F. (2014) ’Employment status and perceived health condition: longitudinal data from Italy’, BMC Public Health, 14(1): 946.

Moher, D., Pham, B., Jones, A., Cook, D. J., Jadad, A. R., Moher, M., Tugwell, P. and Klassen, T. P. (1998) ’Does quality of reports of randomised trials affect estimates of intervention efficacy reported in meta-analyses?’, The Lancet, 352(9128): 609–13.

Nabarro, B. (2021) UK economic outlook: the future isn’t what it used to be., London: Institute for Fiscal Studies.

National Institute for Health and Care Excellence (2012), Methods for the development of NICE public health guidance (third edition). London, NICE.

Office for Budget Responsibility (2021), Economic and fiscal outlook October 2021. London, HMSO

Park, S. J., Kim, S. Y., Lee, E. S. and Park, S. (2020) ’Associations among employment status, health behaviors, and mental health in a representative sample of South Koreans’, International Journal of Environmental Research & Public Health [Electronic Resource], 17(7): 03.

Robert, G., Martínez, J. M., García, A. M., Benavides, F. G. and Ronda, E. (2014) ’From the boom to the crisis: changes in employment conditions of immigrants in Spain and their effects on mental health’, European Journal of Public Health, 24(3): 404–09.

Rönnblad, T., Grönholm, E., Jonsson, J., Koranyi, I., Orellana, C., Kreshpaj, B., Chen, L., Stockfelt, L. and Bodin, T. (2019) ’Precarious employment and mental health: a systematic review and meta-analysis of longitudinal studies’, Scandinavian Journal of Work, Environment & Health, 10.5271/sjweh.3797(5): 429–43.

Scheuring, S. (2020) ’The Effect of Fixed-Term Employment on Well-Being: Disentangling the Micro-Mechanisms and the Moderating Role of Social Cohesion’, Social Indicators Research, 152(1): 91–115.

Scheuring, S., Voßemer, J., Baranowska-Rataj, A. and Tattarini, G. (2021) ’Does Fixed-Term Employment Have Spillover Effects on the Well-Being of Partners? A Panel Data Analysis for East and West Germany’, Journal of Happiness Studies, 22(7): 3001–21.

Sumner, R. C., Bennett, R., Creaven, A. M. and Gallagher, S. (2020) ‘Unemployment, employment precarity, and inflammation’, Brain, Behavior, & Immunity, 83: 303–08.

Taylor, M., Marsh, G., Nicol, D.andBroadbent, P. (2017), Good Work: The Taylor Review of Modern Working Practices. In: E. I. S. Department for Business (ed), London.

Utzet, M., Valero, E., Mosquera, I. and Martin, U. (2020) ’Employment precariousness and mental health, understanding a complex reality: a systematic review’, International Journal of Occupational Medicine and Environmental Health, 33(5): 569–98.

Van Aerden, K., Gadeyne, S. and Vanroelen, C. (2017) ’Is any job better than no job at all? Studying the relations between employment types, unemployment and subjective health in Belgium’, Archives of Public Health, 75: 55.

van der Noordt, M., Ijzelenberg, H., Droomers, M. and Proper, K. I. (2014) ’Health effects of employment: a systematic review of prospective studies’, Occupational and Environmental Medicine, 71(10): 730.

Waddell, G. and Burton, A. K. (2006), Is work good for your health and well-being? London, The Stationery Office.

Yoo, K. B., Park, E. C., Jang, S. Y., Kwon, J. A., Kim, S. J., Cho, K. H., Choi, J. W., Kim, J. H. and Park, S. (2016) ’Association between employment status change and depression in Korean adults’, BMJ Open, 6(3): e008570.

Yoon, S., Kim, J. Y., Park, J. and Kim, S. S. (2017) ’Loss of permanent employment and its association with suicidal ideation: a cohort study in South Korea’, Scandinavian Journal of Work, Environment & Health, 43(5): 457–64.

