## Supplementary material for "Is any job better than no job? : A systematic review"

#### Search strategies

**Databases:** Medline, Medline In-Process & Other Non-Indexed Citations and Daily & Medline Epub Ahead of Print, Embase, PsycInfo, HMIC, ASSIA, TRIP Database, Google Scholar and websites

**Database: Ovid MEDLINE(R) and Epub Ahead of Print, In-Process & Other Non-Indexed Citations and Daily <1946 to April 17, 2020>**
Search Strategy:
--------------------------------------------------------------------------------
1 Unemployment/ (6785)
2 (unemploy* or "un employ*" or "non employ*" or jobseeker? or "lay off*" or layoff* or redundan* or joblessness).ti,ab,kf. (58417)
3 or/1-2 (60929)
4 ("self employ*" or freelance* or "free lance*" or "sole trader*" or soletrader* or entrepreneur*).ti,ab,kf. (4478)
5 Shift Work Schedule/ or Work Schedule Tolerance/ or Workload/ (26852)
6 Working Poor/ (15)
7 Employment/ (45606)
8 work/ (19864)
9 Occupations/ (22804)
10 or/7-9 (85207)
11 uncertainty/ (12296)
12 contract services/ (10523)
13 or/11-12 (22815)
14 and/10,13 (577)
15 ((poor* or heavy or bad or wors* or precari* or unfav* or hard or negative or detrimental or advers* or unsafe* or dangerous* or hazard* or denigrat* or unfair* or insecur* or unstabl* or uncertaint* or instab* or negative or disadvantag* or quality or stress* or strain or night or overnight or demand* or control or autonom* or complex* or unskill* or peripheral or contingent or tempor* or seasonal or "fixed term" or casual or irregular or "long hour?" or unreliable or passive or repetitive or elementary or alienat* or worr* or unhealth* or underpa* or exploitat* or overtime or condition?) adj1 (work* or job? or employ* or labour* or labor* or occupation* or profession* or shift?)).ti,ab,kf. (75179)
16 (shift* adj1 (work* or job? or employ*)).ti,ab,kf. (6819)
17 ((low* or basic or minim* or subminim* or bad or wors* or poor* or bad or precari* or unfav* or hard or negative or detrimental or advers* or unsafe* or dangerous* or hazard* or reduc* or depress* or detrimental* or denigrat* or unfair* or insecur* or unstabl* or uncertaint* or instab* or negative or disadvantag* or quality or shift? or schedul* or night or demand* or strain* or control or autonom* or unskill* or peripheral or contingent or tempor* or "part time" or "fixed term" or casual or hour? or unreliable or elementa* or unhealth* or underpa* or exploitat* or overtime) adj1 (salary or salaries or wage? or earning? or contract?)).ti,ab,kf. (2598)
18 or/4-6,14-17 (106233)
19 and/3,18 (1610)
20 exp cohort studies/ (1979919)
21 (cohort? or longitudinal or prospective or retrospective or "follow* up?").ti,ab,kf. (2285275)
22 Cross-sectional studies/ (324264)
23 ("Cross sectional" or survey? or questionnaire?).ti,ab,kf. (1196220)

24 or/20-23 (4007926)
25 and/19,24 (864)
26 limit 25 to english language (776)
27 (exp africa/ or exp asia/ or exp Americas/) not (Europe/ or exp Australia/ or Austria/ or Belgium/ or exp Canada/ or Chile/ or Czech Republic/ or exp Denmark/ or Estonia/ or Finland/ or France/ or exp Germany/ or Greece/ or Hungary/ or Iceland/ or Ireland/ or Israel/ or exp Italy/ or exp Japan/ or Latvia/ or Lithuania/ or Luxembourg/ or Mexico/ or Netherlands/ or New Zealand/ or exp Norway/ or Poland/ or Portugal/ or Slovenia/ or exp Korea/ or Spain/ or Sweden/ or Switzerland/ or Turkey/ or exp United Kingdom/ or exp United States/) (943515)
28 26 not 27 (679)
29 limit 19 to "reviews (maximizes specificity)" (30)
30 28 or 29 (699)

***************************
**Database: Embase <1974 to 2020 April 17>**
Search Strategy:
--------------------------------------------------------------------------------
1 unemployment/ (19538)
2 (unemploy* or "un employ*" or "non employ*" or jobseeker? or "lay off*" or layoff* or redundan* or joblessness).ti,ab,od. (75882)
3 or/1-2 (75884)
4 self employment/ (657)
5 ("self employ*" or freelance* or "free lance*" or "sole trader*" or soletrader* or entrepreneur*).ti,ab,od. (5652)
6 exp shift work/ or workload/ (45026)
7 Working Poor/ (41)
8 parttime employment/ or temporary employment/ (1804)
9 employment/ (60495)
10 occupation/ (38616)
11 work/ (31492)
12 or/9-10 (97389)
13 uncertainty/ (28200)
14 employment contract/ (2)
15 or/13-14 (28202)
16 and/12,15 (196)
17 ((poor* or heavy or bad or wors* or precari* or unfav* or hard or negative or detrimental or advers* or unsafe* or dangerous* or hazard* or denigrat* or unfair* or insecur* or unstabl* or uncertaint* or instab* or negative or disadvantag* or quality or stress* or strain or night or overnight or demand* or control or autonom* or complex* or unskill* or peripheral or contingent or tempor* or seasonal or "fixed term" or casual or irregular or "long hour?" or unreliable or passive or repetitive or elementary or alienat* or worr* or unhealth* or underpa* or exploitat* or overtime or condition?) adj1 (work* or job? or employ* or labour* or labor* or occupation* or profession* or shift?)).ti,ab,od. (127085)
18 ((low* or basic or minim* or subminim* or bad or wors* or poor* or bad or precari* or unfav* or hard or negative or detrimental or advers* or unsafe* or dangerous* or hazard* or reduc* or depress* or detrimental* or denigrat* or unfair* or insecur* or unstabl* or uncertaint* or instab* or negative or disadvantag* or quality or shift? or schedul* or night or demand* or strain* or control or autonom* or unskill* or peripheral or contingent or tempor* or "part time" or "fixed term" or casual or hour? or unreliable or elementa* or unhealth*

or underpa* or exploitat* or overtime) adj1 (salary or salaries or wage? or earning? or contract?)).ti,ab,od. (3399)
19 (shift* adj1 (work* or job? or employ*)).ti,ab,od. (10319)
20 or/4-8,16-19 (178385)
21 and/3,20 (2616)
22 cohort analysis/ (568188)
23 (cohort? or longitudinal or prospective or retrospective or "follow* up?").ti,ab,od. (3879859)
24 cross-sectional study/ (343646)
25 ("Cross sectional" or survey? or questionnaire?).ti,ab,od. (1734695)
26 or/22-25 (5298158)
27 and/21,26 (1455)
28 limit 27 to english language (1365)
29 (exp africa/ or exp asia/ or exp "South and Central America"/) not (Europe/ or exp Australia/ or Austria/ or Belgium/ or exp Canada/ or Chile/ or Czech Republic/ or exp Denmark/ or Estonia/ or Finland/ or France/ or exp Germany/ or Greece/ or Hungary/ or Iceland/ or Ireland/ or Israel/ or exp Italy/ or exp Japan/ or Latvia/ or Lithuania/ or Luxembourg/ or Mexico/ or Netherlands/ or New Zealand/ or Norway/ or Poland/ or Portugal/ or Slovenia/ or exp Korea/ or Spain/ or Sweden/ or Switzerland/ or Turkey/ or exp United Kingdom/ or exp United States/) (1156687)
30 28 not 29 (1174)
31 limit 21 to "reviews (maximizes specificity)" (51)
32 30 or 31 (1207)

***************************

**Database: APA PsycInfo <1806 to April Week 2 2020>**

Search Strategy:

--------------------------------------------------------------------------------

1 unemployment/ (4226)
2 (unemploy* or "un employ*" or "non employ*" or jobseeker? or "lay off*" or layoff* or redundan* or joblessness).ti,ab,id. (23459)
3 or/1-2 (23717)
4 self-employment/ (379)
5 ("self employ*" or freelance* or "free lance*" or "sole trader*" or soletrader* or entrepreneur*).ti,ab,id. (10178)
6 work scheduling/ or work load/ or Job Security/ (5249)
7 exp working conditions/ (28820)
8 Quality of Work Life/ (1552)
9 employment/ (15478)
10 occupations/ (8791)
11 or/9-10 (23966)
12 uncertainty/ or doubt/ (8539)
13 and/11-12 (46)
14 ((poor* or heavy or bad or wors* or precari* or unfav* or hard or negative or detrimental or advers* or unsafe* or dangerous* or hazard* or denigrat* or unfair* or insecur* or unstabl* or uncertaint* or instab* or negative or disadvantag* or quality or stress* or strain or night or overnight or demand* or control or autonom* or complex* or unskill* or peripheral or contingent or tempor* or seasonal or "fixed term" or casual or irregular or "long hour?" or unreliable or passive or repetitive or elementary or alienat* or worr* or unhealth* or underpa*

or exploitat* or overtime or condition?) adj1 (work* or job? or employ* or labour* or labor* or occupation* or profession* or shift?)).ti,ab,id. (43927)
15 (shift* adj1 (work* or job? or employ*)).ti,ab,id. (2587)
16 ((low* or basic or minim* or subminim* or bad or wors* or poor* or bad or precari* or unfav* or hard or negative or detrimental or advers* or unsafe* or dangerous* or hazard* or reduc* or depress* or detrimental* or denigrat* or unfair* or insecur* or unstabl* or uncertaint* or instab* or negative or disadvantag* or quality or shift? or schedul* or night or demand* or strain* or control or autonom* or unskill* or peripheral or contingent or tempor* or "part time" or "fixed term" or casual or hour? or unreliable or elementa* or unhealth* or underpa* or exploitat* or overtime) adj1 (salary or salaries or wage? or earning? or contract?)).ti,ab,id. (2234)
17 or/4-8,13-16 (80466)
18 and/3,17 (1775)
19 cohort analysis/ (1405)
20 (cohort? or longitudinal or prospective or retrospective or "follow* up?").ti,ab,id. (333433)
21 ("Cross sectional" or questionnaire? or survey?).ti,ab,id. (543668)
22 or/19-21 (803869)
23 and/18,22 (652)
24 limit 23 to english language (590)
25 limit 18 to "reviews (maximizes specificity)" (88)
26 24 or 25 (647)

***************************

**Database: HMIC Health Management Information Consortium <1979 to March 2020>**

Search Strategy:

--------------------------------------------------------------------------------

1 exp unemployment/ (1124)
2 (unemploy* or "un employ*" or "non employ*" or jobseeker? or "lay off*" or layoff* or redundan* or joblessness).af. (2895)
3 or/1-2 (2895)
4 ("self employ*" or freelance* or "free lance*" or "sole trader*" or soletrader* or entrepreneur*).af. (586)
5 exp working hours/ (1386)
6 exp occupational diseases/ (4881)
7 illegal employment/ or part time work/ or seasonal employment/ or temporary employment/ (153)
8 exp "conditions of employment"/ (15349)
9 exp Contracts of employment/ (1015)
10 exp Contract conditions/ (58)
11 ((poor* or heavy or bad or wors* or precari* or unfav* or hard or negative or detrimental or advers* or unsafe* or dangerous* or hazard* or denigrat* or unfair* or insecur* or unstabl* or uncertaint* or instab* or negative or disadvantag* or quality or stress* or strain or night or overnight or demand* or control or autonom* or complex* or unskill* or peripheral or contingent or tempor* or seasonal or "fixed term" or casual or irregular or "long hour?" or unreliable or passive or repetitive or elementary or alienat* or worr* or unhealth* or underpa* or exploitat* or overtime or condition?) adj1 (work* or job? or employ* or labour* or labor* or occupation* or profession* or shift?)).af. (5478)
12 (shift* adj1 (work* or job? or employ*)).af. (484)

13 ((low* or basic or minim* or subminim* or bad or wors* or poor* or bad or precari* or unfav* or hard or negative or detrimental or advers* or unsafe* or dangerous* or hazard* or reduc* or depress* or detrimental* or denigrat* or unfair* or insecur* or unstabl* or uncertaint* or instab* or negative or disadvantag* or quality or shift* or schedul* or night or demand* or strain* or control or autonom* or unskill* or peripheral or contingent or tempor* or "part time" or "fixed term" or casual or hour? or unreliable or elementa* or unhealth* or underpa* or exploitat* or overtime) adj1 (salary or salaries or wage? or earning? or contract?)).af. (395)
14 or/4-13 (25153)
15 and/3,14 (485)
16 exp cohort studies/ (1159)
17 (cohort? or longitudinal or prospective or retrospective or "follow* up?").af. (21176)
18 ("Cross sectional" or survey? or questionnaire?).af. (46755)
19 or/16-18 (61992)
20 and/15,19 (82)
21 (review or "meta analy*" or metaanaly*).ti. (13963)
22 (medline or pubmed or embase or cochrane or psycinfo).ab. (2855)
23 or/21-22 (14957)
24 and/15,23 (4)
25 or/20,24 (83)
26 (review or "meta analy*" or metaanaly*).ti. (13963)
27 (medline or pubmed or embase or cochrane or psycinfo).ab. (2855)
28 or/21-22 (14957)
29 and/15,23 (4)
30 or/20,24 (83)

**************************

**Applied Social Sciences Index & Abstracts (ASSIA)**

Search Strategy

S18 S14 OR S15 OR S16 OR S17 263
S17 S13 AND S7 1
S16 S13 AND S9 195
S15 S13 AND S8 43
S14 S13 AND S6 39
S13 S10 OR S11 OR S12 346,708
S12 ti((review or "meta analys*" or metanalys*)) OR ab((review or "meta analys*" or metanalys*)) 118,989
S11 ti((cohort? OR longitudinal OR prospective OR retrospective OR "follow up" OR "followed up" or "follow ups" OR "followed ups")) OR ab((cohort? OR longitudinal OR prospective OR retrospective OR "follow up" OR "followed up" or "follow ups" OR "followed ups")) 103,802
S10 ti("Cross sectional" or survey? or questionnaire?) OR ab("Cross sectional" or survey? or questionnaire?) 161,450
S9 S1 AND S5 591
S8 S1 AND S4 136
S7 S1 AND S3 7
S6 S1 AND S2 110
S5 ti(((poor* or heavy or bad or wors* or precari* or unfav* or hard or negative or detrimental or advers* or unsafe* or dangerous* or hazard* or

denigrat* or unfair* or insecur* or unstabl* or uncertaint* or instab* or negative or disadvantag* or quality or stress* or strain or night or overnight or demand* or control or autonom* or complex* or unskill* or peripheral or contingent or tempor* or seasonal or "fixed term" or casual or irregular or "long hours" or unreliable or passive or repetitive or elementary or alienat* or worr* or unhealth* or underpa* or exploitat* or overtime or condition?) N/1 (work* or job? or employ* or labour* or labor* or occupation* or profession* or shift?))) OR ab(((poor* or heavy or bad or wors* or precari* or unfav* or hard or negative or detrimental or advers* or unsafe* or dangerous* or hazard* or denigrat* or unfair* or insecur* or unstabl* or uncertaint* or instab* or negative or disadvantag* or quality or stress* or strain or night or overnight or demand* or control or autonom* or complex* or unskill* or peripheral or contingent or tempor* or seasonal or "fixed term" or casual or irregular or "long hours" or unreliable or passive or repetitive or elementary or alienat* or worr* or unhealth* or underpa* or exploitat* or overtime or condition?) N/1 (work* or job? or employ* or labour* or labor* or occupation* or profession* or shift?))) 18,051

S4 ti(((low* or basic or minim* or subminim* or bad or wors* or poor* or bad or precari* or unfav* or hard or negative or detrimental or advers* or unsafe* or dangerous* or hazard* or reduc* or depress* or detrimental* or denigrat* or unfair* or insecur* or unstabl* or uncertaint* or instab* or negative or disadvantag* or quality or shift? or schedul* or night or demand* or strain* or control or autonom* or unskill* or peripheral or contingent or tempor* or "part time" or "fixed term" or casual or hour? or unreliable or elementa* or unhealth* or underpa* or exploitat* or overtime) N/1 (salary or salaries or wage? or earning? or contract?))) OR ab(((low* or basic or minim* or subminim* or bad or wors* or poor* or bad or precari* or unfav* or hard or negative or detrimental or advers* or unsafe* or dangerous* or hazard* or reduc* or depress* or detrimental* or denigrat* or unfair* or insecur* or unstabl* or uncertaint* or instab* or negative or disadvantag* or quality or shift? or schedul* or night or demand* or strain* or control or autonom* or unskill* or peripheral or contingent or tempor* or "part time" or "fixed term" or casual or hour? or unreliable or elementa* or unhealth* or underpa* or exploitat* or overtime) N/1 (salary or salaries or wage? or earning? or contract?))) 1,545

S3 ti((shift* N/1 (work* or job? or employ))) OR ab((shift* N/1 (work* or job? or employ))) 734

S2 ti(("self employed" OR "self employment") or freelance* or ("free lance" OR "free lanced" OR "free lancer" OR "free lancers") or ("sole trader" OR "sole traders") or soletrader* or entrepreneur*) OR ab(("self employed" OR "self employment") or freelance* or ("free lance" OR "free lanced" OR "free lancer" OR "free lancers") or ("sole trader" OR "sole traders") or soletrader* or entrepreneur*) 2,722

S1 ti((unemploy* or "un employment" or "un employed" or "non employed" or "non employment" or jobseeker? or "lay off" or layoff* or redundan* or joblessness)) OR ab((unemploy* or "un employment" or "un employed" or "non employed" or "non employment" or jobseeker? or "lay off" or layoff* or redundan* or joblessness)) 8,201-----------------------------------------------------

**TRIP Database Search**

#3 #1 AND #2 50

#2 (Title:work* or job? or employ* or labour* or labor* or occupation* or profession* or shift? or salary or salaries or wage? or earning? or contract?)

#1 (title:unemploy* or "un employment" or "un employed" or "non employed" or "non employment" or jobseeker? or "lay off" or layoff* or redundan* or joblessness)

******************

**Google scholar searches**

Singular
allintitle:Insecure job unemployment 4
allintitle:Insecure work unemployment 1
allintitle:Insecure employment unemployment 2
allintitle:Insecurity job unemployment 77
allintitle:Insecurity work unemployment 10
allintitle:Insecurity employment unemployment 11
allintitle:bad job unemployment 1
allintitle:bad work unemployment 4
allintitle:bad employment unemployment 5
allintitle:Precarious job unemployment 1
allintitle:Precarious work unemployment 19
allintitle:Precarious employment unemployment 20
allintitle:Insecure job no 3
allintitle:Insecure work no 2
allintitle:Insecure employment no 2
allintitle:bad job no 6
allintitle:bad work no 27
allintitle:bad employment no 3
allintitle:Precarious job no 0
allintitle:Precarious work no 10
allintitle:Precarious employment no 7

Plural
allintitle:Insecure jobs unemployment 6
allintitle:Insecure works unemployment 0
allintitle:Insecure employments unemployment 0
allintitle:Insecurity jobs unemployment 0
allintitle:Insecurity works unemployment 0
allintitle:Insecurity employments unemployment 0
allintitle:bad jobs unemployment 11
allintitle:bad works unemployment 0
allintitle:bad employments unemployment 0
allintitle:Precarious jobs unemployment 5
allintitle:Precarious works unemployment 0
allintitle:Precarious employments unemployment 0
allintitle:Insecure jobs no 1
allintitle:Insecure works no 0

allintitle:Insecure employments no 0
allintitle:bad jobs no 44
allintitle:bad works no 6
allintitle:bad employments no 1
allintitle:Precarious jobs no 0
allintitle:Precarious works no 0
allintitle:Precarious employments no 0
allintitle:work conditions unemployment 17
allintitle:job conditions unemployment 5
allintitle:employment conditions unemployment 41
allintitle:work quality unemployment 19
allintitle:job quality unemployment 68
allintitle:employment quality unemployment 25

Update Search 2021
Date: 14-20 May 2021

Databases: Medline (Proquest Dialog), Embase (Proquest Dialog), PsycInfo (Proquest Dialog), HMIC (OVID), ASSIA (Proquest), TRIP Database, Google Scholar and websites

**Medline ( Proquest Dialog 1946-Current)**

S1 MESH.EXACT("Unemployment") 7204
S2 TI,AB,SU(unemploy* or "un employ*" or "non employ*" or jobseeker[*1] or "lay off*" or layoff* or redundan* or joblessness) 66094
S3 S2 OR S1 66094
S4 TI,AB,SU("self employ*" or freelance* or "free lance*" or "sole trader*" or soletrader* or entrepreneur*) 6517
S5 (MESH.EXACT("Work Schedule Tolerance")) OR MESH.EXACT("Workload") OR (MESH.EXACT("Shift Work Schedule")) 28373
S6 (MESH.EXACT("Working Poor")) 16
S7 MESH.EXACT("Employment") 47222
S8 MESH.EXACT("Work") 20108
S9 MESH.EXACT("Occupations") 23360
S10 S9 OR S8 OR S7 87522
S11 MESH.EXACT("uncertainty") 13793
S12 MESH.EXACT("contract services") 10539
S13 S12 OR S11 24328
S14 S13 AND S10 586
S15 TI,AB((poor* or heavy or bad or wors* or precari* or unfav* or hard or negative or detrimental or advers* or unsafe* or dangerous* or hazard* or denigrat* or unfair* or insecur* or unstabl* or uncertaint* or instab* or negative or disadvantag* or quality or stress* or strain or night or overnight or demand* or control or autonom* or complex* or unskill* or peripheral or contingent or tempor* or seasonal or "fixed term" or casual or irregular or "long hour[*1]" or unreliable or passive or repetitive or elementary or alienat* or worr* or unhealth* or underpa* or exploitat* or overtime or condition[*1]) Near/1 (work* or job[*1] or employ* or labour* or labor* or occupation* or profession* or shift[*1])) 130723
S16 TI,AB,SU(shift* Near/1 (work* or job[*1] or employ*)) 9037

S17 TI,AB((low* or basic or minim* or subminim* or bad or wors* or poor* or bad or precari* or unfav* or hard or negative or detrimental or advers* or unsafe* or dangerous* or hazard* or reduc* or depress* or detrimental* or denigrat* or unfair* or insecur* or unstabl* or uncertaint* or instab* or negative or disadvantag* or quality or shift[*1] or schedul* or night or demand* or strain* or control or autonom* or unskill* or peripheral or contingent or tempor* or "part time" or "fixed term" or casual or hour[*1] or unreliable or elementa* or unhealth* or underpa* or exploitat* or overtime) Near/1 (salary or salaries or wage[*1] or earning[*1] or contract[*1])) 3953
S18 S17 OR S16 OR S15 OR S14 OR S6 OR S5 OR S4 165564
S19 S18 AND S3 2340
S20 (MESH.EXACT.EXPLODE("Cohort Studies")) 2127953
S21 TI,AB(cohort[*1] or longitudinal or prospective or retrospective or "follow* up[*1]") 2509672
S22 (MESH.EXACT.EXPLODE("Cross-sectional studies")) 363786
S23 TI,AB("Cross sectional" or survey[*1] or questionnaire[*1]) 1313512
S24 rtype.exact("Systematic Review") 152912
S25 TI((systematic or rapid) Near/3 (review or meta*)) 159839
S26 rtype.exact("Meta-analysis") 131422
S27 TI(meta-analys* or metaanalys*) 128737
S28 S27 OR S26 OR S25 OR S24 OR S23 OR S22 OR S21 OR S20 4573207
S29 S28 AND S19 1237
S30 (MESH.EXACT.EXPLODE("africa") or MESH.EXACT.EXPLODE("asia") or MESH.EXACT.EXPLODE("Americas")) not (MESH.EXACT("Europe") or MESH.EXACT.EXPLODE("Australia") or MESH.EXACT("Austria") or MESH.EXACT("Belgium") or MESH.EXACT.EXPLODE("Canada") or MESH.EXACT("Chile") or MESH.EXACT("Czech Republic") or MESH.EXACT.EXPLODE("Denmark") or MESH.EXACT("Estonia") or MESH.EXACT("Finland") or MESH.EXACT("France") or MESH.EXACT.EXPLODE("Germany") or MESH.EXACT("Greece") or MESH.EXACT("Hungary") or MESH.EXACT("Iceland") or MESH.EXACT("Ireland") or MESH.EXACT("Israel") or MESH.EXACT.EXPLODE("Italy") or MESH.EXACT.EXPLODE("Japan") or MESH.EXACT("Latvia") or MESH.EXACT("Lithuania") or MESH.EXACT("Luxembourg") or MESH.EXACT("Mexico") or MESH.EXACT("Netherlands") or MESH.EXACT("New Zealand") or MESH.EXACT.EXPLODE("Norway") or MESH.EXACT("Poland") or MESH.EXACT("Portugal") or MESH.EXACT("Slovenia") or MESH.EXACT.EXPLODE("Korea") or MESH.EXACT("Spain") or MESH.EXACT("Sweden") or MESH.EXACT("Switzerland") or MESH.EXACT("Turkey") or MESH.EXACT.EXPLODE("United Kingdom") or MESH.EXACT.EXPLODE("United States")) 1032118
S31 S29 not S30 1095
S32 (S31) and (la.exact("English")) 1003°
******************

**Embase (Proquest Dialog 1947-current)**

S1 EMB.EXACT("unemployment") 23804*
S2 TI,AB,SU(unemploy* or "un employ*" or "non employ*" or jobseeker[*1] or "lay off*" or layoff* or redundan* or joblessness) 85122*
S3 S2 OR S1 85122*
S4 (EMB.EXACT("self employment")) 805°
S5 TI,AB,SU("self employ*" or freelance* or "free lance*" or "sole trader*" or soletrader* or entrepreneur*) 6476*

S6 (EMB.EXACT("shift work")) OR EMB.EXACT("workload") 53461*
S7 EMB.EXACT("Working Poor") 50°
S8 EMB.EXACT("parttime employment") OR EMB.EXACT("temporary employment") 2973°
S9 EMB.EXACT("employment") 78909*
S10 EMB.EXACT("occupation")72849*
S11 EMB.EXACT("work") 49802*
S12 S11 OR S10 OR S9 190384*
S13 EMB.EXACT("uncertainty") 40028*
S14 EMB.EXACT("employment contract") 35°
S15 S14 OR S13 40063*
S16 S15 AND S12 353°
S17 TI,AB((poor* or heavy or bad or wors* or precari* or unfav* or hard or negative or detrimental or advers* or unsafe* or dangerous* or hazard* or denigrat* or unfair* or insecur* or unstabl* or uncertaint* or instab* or negative or disadvantag* or quality or stress* or strain or night or overnight or demand* or control or autonom* or complex* or unskill* or peripheral or contingent or tempor* or seasonal or "fixed term" or casual or irregular or "long hour[*1]" or unreliable or passive or repetitive or elementary or alienat* or worr* or unhealth* or underpa* or exploitat* or overtime or condition[*1]) Near/1 (work* or job[*1] or employ* or labour* or labor* or occupation* or profession* or shift[*1])) 171255*
S18 TI,AB((low* or basic or minim* or subminim* or bad or wors* or poor* or bad or precari* or unfav* or hard or negative or detrimental or advers* or unsafe* or dangerous* or hazard* or reduc* or depress* or detrimental* or denigrat* or unfair* or insecur* or unstabl* or uncertaint* or instab* or negative or disadvantag* or quality or shift[*1] or schedul* or night or demand* or strain* or control or autonom* or unskill* or peripheral or contingent or tempor* or "part time" or "fixed term" or casual or hour[*1] or unreliable or elementa* or unhealth* or underpa* or exploitat* or overtime) Near/1 (salary or salaries or wage[*1] or earning[*1] or contract[*1])) 4757°
S19 TI,AB,SU(shift* Near/1 (work* or job[*1] or employ*)) 15015*
S20 S19 OR S18 OR S17 OR S16 OR S8 OR S7 OR S6 OR S5 OR S4 236374*
S21 S20 AND S3 3340°
S22 (EMB.EXACT("cohort analysis")) 733851*
S23 TI,AB(cohort[*1] or longitudinal or prospective or retrospective or "follow* up[*1]") 3956389*
S24 (EMB.EXACT("cross-sectional study")) 420674*
S25 TI,AB("Cross sectional" or survey[*1] or questionnaire[*1]) 1828153*
S26 rtype.exact("Systematic Review") 0°
S27 (EMB.EXACT("systematic review")) 319992*
S28 TI((systematic or rapid) Near/3 (review or meta*)) 190380*
S29 (EMB.EXACT("meta analysis")) 249750*
S30 TI(meta-analys* or metaanalys*) 160386*
S31 S30 OR S29 OR S28 OR S27 OR S26 OR S25 OR S24 OR S23 OR S22 5839581*
S32 S31 AND S21 1821°
S33 (S32) and (la.exact("English")) 1720°
S34 (EMB.EXACT.EXPLODE("africa") or EMB.EXACT.EXPLODE("asia") or EMB.EXACT.EXPLODE("South and Central America")) not (EMB.EXACT("Europe") or EMB.EXACT.EXPLODE("Australia") or EMB.EXACT("Austria") or EMB.EXACT("Belgium") or EMB.EXACT.EXPLODE("Canada") or

EMB.EXACT("Chile") or EMB.EXACT("Czech Republic") or EMB.EXACT.EXPLODE("Denmark") or EMB.EXACT("Estonia") or EMB.EXACT("Finland") or EMB.EXACT("France") or EMB.EXACT.EXPLODE("Germany") or EMB.EXACT("Greece") or EMB.EXACT("Hungary") or EMB.EXACT("Iceland") or EMB.EXACT("Ireland") or EMB.EXACT("Israel") or EMB.EXACT.EXPLODE("Italy") or EMB.EXACT.EXPLODE("Japan") or EMB.EXACT("Latvia") or EMB.EXACT("Lithuania") or EMB.EXACT("Luxembourg") or EMB.EXACT("Mexico") or EMB.EXACT("Netherlands") or EMB.EXACT("New Zealand") or EMB.EXACT("Norway") or EMB.EXACT("Poland") or EMB.EXACT("Portugal") or EMB.EXACT("Slovenia") or EMB.EXACT.EXPLODE("Korea") or EMB.EXACT("Spain") or EMB.EXACT("Sweden") or EMB.EXACT("Switzerland") or EMB.EXACT("Turkey") or EMB.EXACT.EXPLODE("United Kingdom") or EMB.EXACT.EXPLODE("United States")) 1416408*
S35 S33 not S34 1481°
******************

**PsycInfo (Proquest Dialog)**

Set# Searched for Results
S1 SU.EXACT("Unemployment") 5904
S2 TI,AB,SU(unemploy* or "un employ*" or "non employ*" or jobseeker[*1] or "lay off*" or layoff* or redundan* or joblessness) 25471
S3 S2 OR S1 25471
S4 SU.EXACT("Self-Employment") 484
S5 TI,AB,SU("self employ*" or freelance* or "free lance*" or "sole trader*" or soletrader* or entrepreneur*) 11392
S6 (SU.EXACT("work scheduling")) OR (SU.EXACT("work load")) OR (SU.EXACT("Job security")) 5749
S7 (SU.EXACT.EXPLODE("Working Conditions")) 30443
S8 (SU.EXACT("Quality of Work Life")) 1651
S9 (SU.EXACT("employment")) 13001
S10 (SU.EXACT("occupations")) 13043
S11 S10 OR S9 25432
S12 SU.EXACT("uncertainty") OR SU.EXACT("doubt") 11392
S13 S12 AND S11 63
S14 TI,AB((poor* or heavy or bad or wors* or precari* or unfav* or hard or negative or detrimental or advers* or unsafe* or dangerous* or hazard* or denigrat* or unfair* or insecur* or unstabl* or uncertaint* or instab* or negative or disadvantag* or quality or stress* or strain or night or overnight or demand* or control or autonom* or complex* or unskill* or peripheral or contingent or tempor* or seasonal or "fixed term" or casual or irregular or "long hour[*1]" or unreliable or passive or repetitive or elementary or alienat* or worr* or unhealth* or underpa* or exploitat* or overtime or condition[*1]) Near/1 (work* or job[*1] or employ* or labour* or labor* or occupation* or profession* or shift[*1])) 66094
S15 TI,AB(shift* Near/1 (work* or job[*1] or employ*))3322
S16 TI,AB((low* or basic or minim* or subminim* or bad or wors* or poor* or bad or precari* or unfav* or hard or negative or detrimental or advers* or unsafe* or dangerous* or hazard* or reduc* or depress* or detrimental* or denigrat* or unfair* or insecur* or unstabl* or uncertaint* or instab* or negative or disadvantag* or quality or shift[*1] or schedul* or night or demand* or strain* or control or autonom* or unskill* or peripheral or contingent or tempor* or "part

time" or "fixed term" or casual or hour[*1] or unreliable or elementa* or unhealth* or underpa* or exploitat* or overtime) Near/1 (salary or salaries or wage[*1] or earning[*1] or contract[*1])) 3124
S17 S16 OR S15 OR S14 OR S13 OR S8 OR S7 OR S6 OR S5 OR S4 105168
S18 S17 AND S3 2370
S19 (SU.EXACT("Cohort Analysis")) 1546
S20 TI,AB(cohort[*1] or longitudinal or prospective or retrospective or "follow* up[*1]") 353216
S21 TI,AB("Cross sectional" or questionnaire[*1] or survey[*1]) 580097
S22 (SU.EXACT("Systematic Review")) 31483
S23 TI((systematic or rapid) Near/3 (review or meta*)) 25840
S24 (SU.EXACT("Meta Analysis")) 27526
S25 TI(meta-analys* or metaanalys*) 20211
S26 S25 OR S24 OR S23 OR S22 OR S21 OR S20 OR S19 900878
S27 S26 AND S18 890
S28 (S27) and (la.exact("English")) 838°

******************

**HMIC (OVID) Health Management Information Consortium <1979 to March 2021>**

1 systematic reviews/ (3196)
2 meta analysis/ (778)
3 exp unemployment/ (1140)
4 (unemploy* or "un employ*" or "non employ*" or jobseeker? or "lay off*" or layoff* or redundan* or joblessness).af. (2928)
5 or/3-4 (2928)
6 ("self employ*" or freelance* or "free lance*" or "sole trader*" or soletrader* or entrepreneur*).af. (598)
7 exp working hours/ (1394)
8 exp occupational diseases/ (4881)
9 illegal employment/ or part time work/ or seasonal employment/ or temporary employment/ (154)
10 exp "conditions of employment"/ (15511)
11 exp Contracts of employment/ (1020)
12 exp Contract conditions/ (58)
13 ((poor* or heavy or bad or wors* or precari* or unfav* or hard or negative or detrimental or advers* or unsafe* or dangerous* or hazard* or denigrat* or unfair* or insecur* or unstabl* or uncertaint* or instab* or negative or disadvantag* or quality or stress* or strain or night or overnight or demand* or control or autonom* or complex* or unskill* or peripheral or contingent or tempor* or seasonal or "fixed term" or casual or irregular or "long hour?" or unreliable or passive or repetitive or elementary or alienat* or worr* or unhealth* or underpa* or exploitat* or overtime or condition?) adj1 (work* or job? or employ* or labour* or labor* or occupation* or profession* or shift?)).af. (5578)
14 (shift* adj1 (work* or job? or employ*)).af. (485)
15 ((low* or basic or minim* or subminim* or bad or wors* or poor* or bad or precari* or unfav* or hard or negative or detrimental or advers* or unsafe* or dangerous* or hazard* or reduc* or depress* or detrimental* or denigrat* or unfair* or insecur* or unstabl* or uncertaint* or instab* or negative or disadvantag* or quality or shift* or schedul* or night or demand* or strain* or control or autonom* or unskill* or peripheral or contingent or tempor* or "part

time" or "fixed term" or casual or hour? or unreliable or elementa* or unhealth* or underpa* or exploitat* or overtime) adj1 (salary or salaries or wage? or earning? or contract?)).af. (402)
16 or/6-15 (25389)
17 and/5,16 (494)
18 exp cohort studies/ (1160)
19 (cohort? or longitudinal or prospective or retrospective or "follow* up?").af. (21368)
20 ("Cross sectional" or survey? or questionnaire?).af. (47267)
21 or/18-20 (62620)
22 and/17,21 (87)
23 systematic reviews/ or meta analysis/ (3607)
24 (review or "meta analy*" or metaanaly*).ti. (14168)
25 (medline or pubmed or embase or cochrane or psycinfo).ab. (2918)
26 or/23-25 (15819)
27 and/17,26 (5)
28 or/22,27 (89)

******************

**Applied Social Sciences Index & Abstracts (ASSIA) (Proquest 1987-current)**

S1 ti((unemploy* or "un employment" or "un employed" or "non employed" or "non employment" or jobseeker[*1] or "lay off" or layoff* or redundan* or joblessness)) OR ab((unemploy* or "un employment" or "un employed" or "non employed" or "non employment" or jobseeker[*1] or "lay off" or layoff* or redundan* or joblessness)) Applied Social Sciences Index & Abstracts (ASSIA) 8546

S2 ti(("self employed" OR "self employment") or freelance* or ("free lance" OR "free lanced" OR "free lancer" OR "free lancers") or ("sole trader" OR "sole traders") or soletrader* or entrepreneur*) OR ab(("self employed" OR "self employment") or freelance* or ("free lance" OR "free lanced" OR "free lancer" OR "free lancers") or ("sole trader" OR "sole traders") or soletrader* or entrepreneur*) Applied Social Sciences Index & Abstracts (ASSIA) 2872

S3 ti((shift* N/1 (work* or job[*1] or employ))) OR ab((shift* N/1 (work* or job[*1] or employ))) Applied Social Sciences Index & Abstracts (ASSIA) 793

S4 ti(((low* or basic or minim* or subminim* or bad or wors* or poor* or bad or precari* or unfav* or hard or negative or detrimental or advers* or unsafe* or dangerous* or hazard* or reduc* or depress* or detrimental* or denigrat* or unfair* or insecur* or unstabl* or uncertaint* or instab* or negative or disadvantag* or quality or shift[*1] or schedul* or night or demand* or strain* or control or autonom* or unskill* or peripheral or contingent or tempor* or "part time" or "fixed term" or casual or hour[*1] or unreliable or elementa* or unhealth* or underpa* or exploitat* or overtime) N/1 (salary or salaries or wage[*1] or earning[*1] or contract[*1]))) OR ab(((low* or basic or minim* or subminim* or bad or wors* or poor* or bad or precari* or unfav* or hard or negative or detrimental or advers* or unsafe* or dangerous* or hazard* or reduc* or depress* or detrimental* or denigrat* or unfair* or insecur* or unstabl* or uncertaint* or instab* or negative or disadvantag* or quality or shift[*1] or schedul* or night or demand* or strain* or control or autonom* or unskill* or peripheral or contingent or tempor* or "part time" or "fixed term" or casual or hour[*1] or unreliable or elementa* or unhealth* or underpa* or exploitat* or overtime) N/1 (salary or

salaries or wage[*1] or earning[*1] or contract[*1]))) Applied Social Sciences Index & Abstracts (ASSIA) 1607
S5 ti(((poor* or heavy or bad or wors* or precari* or unfav* or hard or negative or detrimental or advers* or unsafe* or dangerous* or hazard* or denigrat* or unfair* or insecur* or unstabl* or uncertaint* or instab* or negative or disadvantag* or quality or stress* or strain or night or overnight or demand* or control or autonom* or complex* or unskill* or peripheral or contingent or tempor* or seasonal or "fixed term" or casual or irregular or "long hours" or unreliable or passive or repetitive or elementary or alienat* or worr* or unhealth* or underpa* or exploitat* or overtime or condition[*1]) N/1 (work* or job[*1] or employ* or labour* or labor* or occupation* or profession* or shift[*1]))) OR ab(((poor* or heavy or bad or wors* or precari* or unfav* or hard or negative or detrimental or advers* or unsafe* or dangerous* or hazard* or denigrat* or unfair* or insecur* or unstabl* or uncertaint* or instab* or negative or disadvantag* or quality or stress* or strain or night or overnight or demand* or control or autonom* or complex* or unskill* or peripheral or contingent or tempor* or seasonal or "fixed term" or casual or irregular or "long hours" or unreliable or passive or repetitive or elementary or alienat* or worr* or unhealth* or underpa* or exploitat* or overtime or condition[*1]) N/1 (work* or job[*1] or employ* or labour* or labor* or occupation* or profession* or shift[*1]))) Applied Social Sciences Index & Abstracts (ASSIA) 18955
S6 S1 AND S2 Applied Social Sciences Index & Abstracts (ASSIA)
These databases are searched for part of your query. 119
S7 S1 AND S3 Applied Social Sciences Index & Abstracts (ASSIA)
These databases are searched for part of your query. 7
S8 S1 AND S4 Applied Social Sciences Index & Abstracts (ASSIA)
These databases are searched for part of your query. 140
S9 S1 AND S5 Applied Social Sciences Index & Abstracts (ASSIA)
These databases are searched for part of your query. 612
S10 ti("Cross sectional" or survey[*1] or questionnaire[*1]) OR ab("Cross sectional" or survey[*1] or questionnaire[*1]) Applied Social Sciences Index & Abstracts (ASSIA) 173339
S11 ti((cohort[*1] OR longitudinal OR prospective OR retrospective OR "follow up" OR "followed up" or "follow ups" OR "followed ups")) OR ab((cohort[*1] OR longitudinal OR prospective OR retrospective OR "follow up" OR "followed up" or "follow ups" OR "followed ups")) Applied Social Sciences Index & Abstracts (ASSIA) 111201
S12 ti((review or ("meta analyses" OR "meta analysis") or metanalys*)) OR ab((review or ("meta analyses" OR "meta analysis") or metanalys*)) Applied Social Sciences Index & Abstracts (ASSIA) 125351
S13 S10 OR S11 OR S12 Applied Social Sciences Index & Abstracts (ASSIA)
These databases are searched for part of your query. 368983
S14 S13 AND S6 Applied Social Sciences Index & Abstracts (ASSIA)
These databases are searched for part of your query. 42
S15 S13 AND S8 Applied Social Sciences Index & Abstracts (ASSIA)
These databases are searched for part of your query. 42
S16 S13 AND S9 Applied Social Sciences Index & Abstracts (ASSIA)
These databases are searched for part of your query. 203
S17 S13 AND S7 Applied Social Sciences Index & Abstracts (ASSIA)
These databases are searched for part of your query. 1

******************

**TRIP Database Search**

#3 #1 AND #2 64

#2 (Title:work* or job? or employ* or labour* or labor* or occupation* or profession* or shift? or salary or salaries or wage? or earning? or contract?)

#1 (title:unemploy* or "un employment" or "un employed" or "non employed" or "non employment" or jobseeker? or "lay off" or layoff* or redundan* or joblessness)

******************

**Google scholar Searches**

Singular
allintitle:Insecure job unemployment 4
allintitle:Insecure work unemployment 2
allintitle:Insecure employment unemployment 2
allintitle:Insecurity job unemployment 85
allintitle:Insecurity work unemployment 9
allintitle:Insecurity employment unemployment 12
allintitle:bad job unemployment 1
allintitle:bad work unemployment 4
allintitle:bad employment unemployment 7
allintitle:Precarious job unemployment 1
allintitle:Precarious work unemployment 22
allintitle:Precarious employment unemployment 28
allintitle:Insecure job no 2
allintitle:Insecure work no 2
allintitle:Insecure employment no
allintitle:bad job no 1
allintitle:bad work no 26
allintitle:bad employment no 4
allintitle:Precarious job no 0
allintitle:Precarious work no 11
allintitle:Precarious employment no 8

Plural
allintitle:Insecure jobs unemployment 6
allintitle:Insecure works unemployment 0
allintitle:Insecure employments unemployment 0
allintitle:Insecurity jobs unemployment 0
allintitle:Insecurity works unemployment 0
allintitle:Insecurity employments unemployment 0
allintitle:bad jobs unemployment 12
allintitle:bad works unemployment 0
allintitle:bad employments unemployment 0
allintitle:Precarious jobs unemployment 6
allintitle:Precarious works unemployment 0

allintitle:Precarious employments unemployment  0
allintitle:Insecure jobs no   0
allintitle:Insecure works no   0
allintitle:Insecure employments no  0
allintitle:bad jobs no   48
allintitle:bad works no  7
allintitle:bad employments no   1
allintitle:Precarious jobs no  0
allintitle:Precarious works no  0
allintitle:Precarious employments no   0
allintitle:work conditions unemployment   26
allintitle:job conditions unemployment     4
allintitle:employment conditions unemployment   51
allintitle:work quality unemployment   33
allintitle:job quality unemployment    89
allintitle:employment quality unemployment   38

**Websites**

What Works well-being (Work) https://whatworkswellbeing.org/product-category/work/     2
ILO International labor organisation https://www.ilo.org/global/lang--en/index.htm
Labordoc (ILO digital repository)
https://labordoc.ilo.org/discovery/search?vid=41ILO_INST:41ILO_V2&lang=en
Eurofound  European Foundation for the Improvement of Living and Working Conditions
https://www.eurofound.europa.eu/publications
IES Institute for employment studies https://www.employment-studies.co.uk/
IZA Institute of Labor economics https://www.iza.org/
OECD Employment
https://www.oecd.org/employment/job-quality.htm
https://www.oecd.org/health/mental-health-and-work.htm
Work Foundation http://www.theworkfoundation.com/
Joseph Rowntree foundation https://www.jrf.org.uk/

### Studies excluded at full text

| Source | Reason for exclusion |
|---|---|
| Ali SM and Lindstrom, M. (2006) Psychosocial work conditions, unemployment, and leisure-time physical activity: a population-based study. *Scand J of Public Health* 34 (2):pp.209-16 | Study sample not drawn from a nationally representative population |
| Ali SM and Lindstrom M (2008) Psychosocial work conditions, unemployment and health locus of control: a population-based study *Scand J Public Health* 36(4):pp.429-435 | Outcome not relevant, does not meet inclusion criteria |
| Amick BC et al. (2002). Relationship between all-cause mortality and cumulative working life course psychosocial and physical exposures in the United States labor market from 1968 to 1992. *Psychosom Med* 64(3):pp.370-81 | No unemployed comparison |
| Anderson J G (2002). Coping with long-term unemployment: Economic security, labour market integration and well-being: Results from a Danish panel study, 1994-1999. *International Journal of Social Welfare*. 11(3):pp.178-190 | No unemployed comparison |
| Arnetz BB et al (1991) Neuroendocrine and immunologic effects of unemployment and job insecurity. *Psychother Psychosom* 55 (2-4):pp.76-80 | No relevant between group comparison |
| Backhaus M and Zhang W. (2011) Unemployment and mental health who is (not) affected? Evaluation and equity audit of the domestic radon programme in England. Eur J Public Health 22(3):pp.429-433 | No relevant comparison |
| Berth, H et al. (2008) Unemployment, job insecurity and the need for psychosocial support. *Arbeitslosigkeitserfahrungen, Arbeitsplatzunsicherheit und der Bedarf an psychosozialer Versorgung.* 70(5): pp.289-294 | German, unable to translate |
| Berth H et al. (2006) Unemployment, job insecurity and life satisfaction: Results of a study with young adults in the new German states. *Sozial-und Praventivmedizin* 50(6):361-9 | German, unable to translate |
| Berth H, Forster P, Brahler E. (2003) Unemployment, job insecurity and their consequences for health in a sample of young adults. *Bundesverband der Arzte des Offentlichen Gesundheitsdienstes* 65(10):pp.555-560 | German, unable to translate |
| Blanquet, M. et al. (2017) Occupational status as determinant of mental health inequities in French young people: is fairness needed? Results of a cross-sectional multicentre observational survey. *International Journal for Equity in Health* 16(1):pp.142 | Study sample not drawn from a nationally representative population |
| Borra C and Garcia FG. Job satisfaction, insecurity and the Great Recession: The effect of others´ unemployment. *XX Encuentro Economía Pública: Estado …* | Unable to obtain copy of paper |
| Brenner SO and Starrin B (1998) Unemployment and health in Sweden: Public issues and private trouble. *Journal of Social Issues.* 44(4):pp.125-140 | No relevant comparison |
| Brockner J et al.(1992) Layoffs, job insecurity, and survivors' work effort: Evidence of an inverted-U relationship. *Academy of Management Journal* 35(2):pp.413-425 | No unemployed comparison group |
| Broom D et al. (2006) The lesser evil: bad jobs or unemployment? A survey of mid-aged Australians. *Soc Sci Med* 63(3):pp.576-86 | Study sample not drawn from a nationally representative population |
| Brydsten A, Hammarstrom, A and San Sebastian, M. (2018). Health inequalities between employed and unemployed in northern Sweden: a decomposition analysis of social determinants for mental health. *International Journal for Equity in Health* 17(7):pp.59 | No relevant comparison |
| Burchell B. (2011) A temporal comparison of the effects of unemployment and job insecurity on wellbeing. *Sociological Research Online* 16(1):9 | No unemployed comparison |
| Burgard SA, Brand JE. and House JS (2009) Perceived job insecurity and worker health in the United States *Soc Sci Med* 69(5):777-785 | No relevant comparison |

| Source | Reason for exclusion |
|---|---|
| Campbell-Jamison F, Worrall L and Cooper (2001) Downsizing in Britain and its effects on survivors and their organizations *Anxiety, Stress & Coping: An International Journal* 14 (1):pp.35-58 | Not a relevant study, no relevant comparison |
| Canivet C et al. (2016) Precarious employment is a risk factor for poor mental health in young individuals in Sweden: a cohort study with multiple follow-ups. *BMC Public Health* 16:pp. 687 | No relevant comparison |
| Chambel MJ, Lopes S. and Batista J. (2016) The effects of temporary agency work contract transitions on well-being Int Arch Occup Environ Health 89(9):pp. 1215-1228 | Study sample not drawn from a nationally representative population and no relevant comparison |
| Charles KK and Decicca P (2008) Local labor market fluctuations and health: is there a connection and for whom? *J Health Econ* 27(6):pp.1532-50 | Not a relevant study, no relevant comparison |
| Clark A, Knabe A and Ratzel S (2010) Boon or bane? Others' unemployment, well-being and job insecurity *Labour Economics* 17:pp.52-61 | Did not report any relevant outcomes |
| Costes FXB, Julia M and Benach J (2016) What is the impact of temporary employment on self-assessed health? A panel study in Catalonia 2002-2012 *Occupational and Environmental Medicine* 73(Suppl 1)pp.A39 | Conference abstract, does not meet inclusion criteria for source type |
| Dalglish SL et al. (2015) Work characteristics and suicidal ideation in young adults in France Soc Psychiatry Psychiatr Epidemiol 50 (4):pp.613-620 | Study sample not drawn from a nationally representative population |
| De Witte H (1999) Job insecurity and psychological well-being: Review of the literature and exploration of some unresolved issues *European Journal of Work and Organizational Psychology* 8(2):155-177 | No relevant comparison |
| Dekker SW and Schaufeli WB (1995) The effects of job insecurity on psychological health and withdrawal: A longitudinal study *Aust Psychol* 30(1): pp.57-63 | No relevant comparison |
| Dooley D, Fielding J and Levi L. (1996) Health and unemployment. *Annu Rev Public Health* 17:pp.449-465 | Not a relevant study |
| Dooley D, Prause J (1998) Unemployment and alcohol misuse in the National Longitudinal Survey of Youth. *J Stud Alcohol* 59(6): pp.669-80 | No relevant comparison |
| Estrella ML et al. (2018) The association of employment status with ideal cardiovascular health factors and behaviors among Hispanic/Latino adults: Findings from the Hispanic Community Health Study/Study of Latinos (HCHS/SOL) *PLoS ONE* 13(11): e0207652 | Study sample not drawn from a nationally representative population |
| Farrants K et al. (2020) Job demands and job control and future labor market situation: An 11-year prospective study of 2.2 million employees. *Occup. Environ. Med.* 62(6): pp.403-411 | No relevant comparison or outcomes |
| Feather NT and O'Brien GE (1986) A longitudinal analysis of the effects of different patterns of employment and unemployment on school-leavers *British journal of* psychology 77:pp.4/- | No relevant comparison |
| Ferrie, JE (1997) Labour market status, insecurity and health. *J Health Psychol* 2(3):pp.373-397 | Not a relevant study |
| Ferrie JE et al. (2001) Employment status and health after privatisation in white collar civil servants: Prospective cohort study *BMJ* 322(7287):pp.647-651 | Study sample not drawn from a nationally representative population |
| Ferrie JE et al. (2001) Job insecurity in white-collar workers: toward an explanation of associations with health *J Occup Health Psychol* 6(1):pp.26-42 | Not a relevant study, no relevant comparison |
| Ferrie JE et al. (2003) Future uncertainty and socioeconomic inequalities in health: the Whitehall II study *Soc Sci Med* 57(4):pp.637-46 | Not a relevant study, no relevant comparison |
| Flovik L, Knardahl S and Christensen JO (2019) Organizational change and employee mental health: A prospective multilevel study | No relevant comparison |

| Source | Reason for exclusion |
|---|---|
| of the associations between organizational changes and clinically relevant mental distress *Scand J Work Environ Health* 45(2):pp.134-145 | |
| Friedmann ML and Webb AA (1995) Family health and mental health six years after economic stress and unemployment. *Issues in Mental Health Nursing* 16(1):pp.51-66 | Not a relevant study, no relevant comparison |
| Ganson KT et al. (2021) Job insecurity and symptoms of anxiety and depression among young US adults during COVID-19. *J Adolesc Health* 68(1): pp.53-56 | No relevant comparison |
| Giudici F and Morselli D (2019) 20 Years in the world of work: A study of (nonstandard) occupational trajectories and health *Soc Sci Med* 224:pp.138-148 | No relevant comparison |
| Graetz B (1993) Health consequences of employment and unemployment: longitudinal evidence for young men and women *Soc Sci Med* 36(6):pp.715-724 | No relevant comparison |
| Gray BJ et al (2019) Employment status and impact on mental wellbeing in the UK working age population: a cross-sectional analysis *Lancet* 394(Suppl 2):pp.S44 | Did not meet inclusion criteria for source type |
| Hallsten L, Grossi G, Westerlund H (1999) Unemployment, labour market policy and health in Sweden during years of crisis in the 1990's *Int Arch Occup Environ Health* 72Suppl:pp.S28-30 | Unable to obtain full text copy |
| Hernando-Rodriguez JC et al. (2020) Sickness absence trajectories following labour market participation patterns: a cohort study in Catalonia (Spain), 2012-2014 *BMC Public Health* 20(1): pp.1306 | No unemployed comparator |
| Hibbard, JH and Pope CR (1985) Employment status, employment characteristics, and women's health *Women & Health* 10(1):pp.59-77 | No relevant comparison |
| Hughes A et al. (2015) Elevated inflammatory biomarkers during unemployment: modification by age and country in the UK Jo Epidemiol Community Health 69(7):pp. 673-679 | No relevant comparison |
| Iversen L and Sabroe S (1998) Psychological well-being among unemployed and employed people after a company closedown: a longitudinal study *Journal of Social Issues*: 44 (Winter 88):pp.141-152 | No relevant comparison |
| Izdebski ZW, Mazur J (2021) Changes in mental well-being of adult Poles in the early period of the COVID-19 pandemic with reference to their occupational activity and remote work *Int J Occup Med Environ Health* 34(2): pp.251-262 | Does not meet inclusion criteria for population |
| Jamison CS, Wallace M and Jamison PL (2004) Contemporary work characteristics, stress, and ill health *Am J Human Biol* 16(1):pp.43-56 | No relevant comparison |
| Khlat M et al. (2014) Mortality gradient across the labour market core-periphery structure: a 13-year mortality follow-up study in north-eastern France *Int Arch Occup Environ Health* 87(7): pp.725-733 | Study sample not drawn from a nationally representative population |
| Kivimaki M et al. (2003) Temporary employment and risk of overall and cause-specific mortality Am J Epidemiol 158(7):pp.663-668 | Study sample not drawn from a nationally representative population |
| Kwon K et al (2016) Association between employment status and self-rated health: Korean working conditions survey *Ann Occup Environ Med* 28(1 43) DOI 10.1186/s40557-016-0126-z | No relevant comparison |
| LaMontagne AD et al. (2016) Psychosocial job quality, mental health, and subjective wellbeing: a cross-sectional analysis of the baseline wave of the Australian Longitudinal Study on Male Health *BMC Public Health* 16 (Suppl 3):pp.1049 | No relevant comparison |
| Leach LS et al. (2010) The limitations of employment as a tool for social inclusion *BMC Public Health* 10(61):pp.621 doi 10.1186/1471-2458-10-621 | Study sample not drawn from a nationally representative population |
| Lee ES and Park S (2019) Patterns of change in employment status and their association with self-rated health, perceived daily stress, | No relevant comparison |

| Source | Reason for exclusion |
|---|---|
| and sleep among young adults in South Korea *Int J Environ Res Public Health* 16(4491):doi:10.3390/ijerph16224491 | |
| Levi L et al. (1984) The psychological, social, and biochemical impacts of unemployment in Sweden: Description of a research project *Int J Mental Health* 13(1-2):pp.18-34 | No relevant comparison |
| Lindstrom M (2004) Psychosocial work conditions, social capital, and daily smoking: a population based study *Tob Control* 13(3):pp.289-295 | Study sample not drawn from a nationally representative population |
| Lindstrom M (2005) Psychosocial work conditions, unemployment and self-reported psychological health: a population-based study *Occup Med (Oxf)* 55(7):pp.568-71 | Study sample not drawn from a nationally representative population |
| Lindstrom M, Ali SM and Rosvall M (2012) Socioeconomic status, labour market connection, and self-rated psychological health: the role of social capital and economic stress *Scand J Public Health* 40(1):pp.51-60 | No relevant comparison, study sample not drawn from a nationally representative population |
| Livanos I and Zangelidis A (2013) Unemployment, labor market flexibility, and absenteeism: A pan-European study *Industrial Relations: A Journal of Economy & Society* 52(2):pp.492-515 | No relevant comparison |
| Lopez Gomez MA et al. (2017) Employment history indicators and mortality in a nested case-control study from the Spanish WORKing life social security (WORKss) cohort *PLoS ONE* 16(6):pp.e0178486 | No relevant comparison |
| Lopez MA et al. (2016) Labour market trajectories and mortality *Occup Environ Med* 73(suppl1) | Conference abstract does not meet inclusion criteria for source type |
| Makikangas A et al. (2011) A person-centred approach to investigate the development trajectories of job-related affective well-being: A 10-year follow-up study *J Occup Org Psychol* 84(2):pp.327-346 | No relevant comparison |
| Marchand A, Drapeau A and Beaulieu-Prevost D 2011 Employment, non-employment and psychological distress in Canada *Am J Epidemiol* 11:pp.S155 | Conference abstract does not meet inclusion criteria for source type |
| Marchand A, Drapeau A and Beaulieu-Prevost D (2012) Psychological distress in Canada: the role of employment and reasons of non-employment *Int J Soc Psychiatry* 58(6):pp.596-604 | No relevant comparison |
| Mencia PM, Prieto DC (2021) Job status and depressive symptoms in older employees: An empirical analysis with SHARE (survey of health, aging and retirement in Europe) data *Eur J Ment Health* 15(2): pp.168-177 | No relevant poor job comparison |
| Montfort SS et al. (2015) A longitudinal examination of re-employment quality on internalizing symptoms and job-search intentions *J Occup Health Psychol* 20(1):pp.50-61 | No relevant comparison |
| Morrell S et al. (1994) A cohort study of unemployment as a cause of psychological disturbance in Australian youth *Soc Sci Med* 38(11):pp.1553-64 | No relevant comparison |
| O'Brien GE and Feather NT (1990) The relative effects of unemployment and quality of employment on the affect, work values and personal control of adolescents *J Occup Psychol* 63:pp.51-165 | Study sample not drawn from a nationally representative population |
| Otto K and Dalbert C (2013) Are insecure jobs as bad for mental health and occupational commitment as unemployment? Equal threat or downward spiral *Horizons of Psychology* 22:pp.27-38A et al | Study sample not drawn from a nationally representative population |
| Page A et al (2013) The role of under-employment and unemployment in recent birth cohort effects in Australian suicide *Soc Sci Med* 93:pp.155-162 | No relevant comparison |
| Park S et al. (2020) Precarious employment as compared with unemployment reduces the risk of depression in the elderly in Korea. *J Occup Environ Med* 62(10): ppe559-e566 | Does not meet inclusion criteria for population |
| Perreault M et al (2020) Transitional Employment and Psychological Distress: a Longitudinal Study *Psychiatr Q* 25:pp.25 | No relevant comparison |

| Source | Reason for exclusion |
|---|---|
| Raymo JM and Shibata A (2017) Unemployment, non-standard employment, and fertility: Insights from Japans "Lost 20 Years" *Demography* 54(6):pp.2301-2329 | Outcome not relevant, did not meet inclusion criteria |
| Robert G et al (2014) From the boom to the crisis: changes in employment conditions of immigrants in Spain and their effects on mental health *Eur J Public Health* 24(3):pp.404-409 | Study sample not drawn from a nationally representative population (Nb. this should have been excluded at title) |
| Rugulies R et al. (2010) Occupational position and its relation to mental distress in a random sample of Danish residents *Int Arch of Occup Environ Health* 83(6):pp.625-629 | No relevant comparison |
| Rugulies R et al (2010) Job insecurity and the use of antidepressant medication among Danish employees with and without a history of prolonged unemployment: a 3.5-year follow-up study *J Epidemiol Community Health* 64(1):pp.75-81 | No relevant comparison |
| Russo C, Terraneo M (2020) Mental well-being among workers: A cross national analysis of job insecurity impact on the workforce. *Soc Indic Res* 152: pp.421-442 | No unemployed comparator |
| Scheuring S (2020) The effect of fixed-term employment on well-being: Disentangling the micro-mechanisms and the moderating role of social cohesion. *Soc Indic Res* 152: pp.91-115 | No relevant comparison – comparison is at country level |
| Stokes G and Cochrane R (1984) A study of the psychological effects of redundancy and unemployment *J Occup Psychol* 57(4):pp.309-322 | No relevant comparison |
| Strandh M (2000 Different exit routes from unemployment and their impact on mental well-being: The role of the economic situation and the predictability of the life course *Work, Employment and Society* 14(3):pp.459-479 | No relevant comparison |
| Vanthomme K et al (2017) Site-specific cancer mortality inequalities by employment and occupational groups: a cohort study among Belgian adults, 2001-2011 *BMJ Open* 7(11):pp.e015216 | No relevant comparison |
| Virtanen P et al (2008) Employment trajectory as determinant of change in health-related lifestyle: the prospective HeSSup study *Eur J Public Health* 18(5)pp.504-508 | No relevant comparison |
| Wanberg R (1995) A longitudinal study of the effects of unemployment and quality of reemployment *Journal of Vocational Behavior* 46(1):pp.40-54 | No relevant comparison |
| Wege N, Angerer P and Li J(2017) Effects of Lifetime Unemployment Experience and Job Insecurity on Two-Year Risk of Physician-Diagnosed Incident Depression in the German Working Population *Int J Environ Res Public Health* 14(8):pp.11 | No relevant comparison |
| Wolinska W et al. (2020) Analysis of the relationship between insomnia and adult chronic diseases with regard to working conditions. *Fam Med Prim* 22(3): pp.228-234 | No relevant poor job comparator |

### Table S1. Data extraction and analysis

#### Full results of all included studies

<table>
<tr><th>Reference/<br>Study design</th><th>Data source/<br>Population range</th><th>Poor/bad job definition<br>Outcome</th><th>Results from paper</th><th>Is any job better than no job?<br>Where OR has been calculated by OES exposure is bad/poor job, is unadjusted and data used to calculate may have been derived from percentages in study results tables</th></tr>
<tr><td>Bently R et al. 2020<br><br>Cross sectional</td><td>Household Income and Labour Dynamics in Australia (HILDA) 2018<br><br>25 to 64 years</td><td>Insecure employment, included self-employment, casual or labour hire but not fixed term contracts<br><br>Alcohol and tobacco use</td><td>Odds ratio and 95% confidence intervals. The effect of economic security on tobacco and alcohol consumption (table 4, page 1888)<br><br><table><tr><th></th><th>Smoking status (yes/no)</th><th>Drinking status (yes/no)</th><th>Risky drinking (yes/no)</th></tr><tr><td>Secure employment</td><td>Ref</td><td>Ref</td><td>Ref</td></tr><tr><td>Insecure employment</td><td>1.01 (0.88 to 1.14)</td><td>0.87 (0.77 to 0.98)</td><td>0.98 (0.89 to 1.08)</td></tr><tr><td>Unemployed</td><td>1.32 (1.07 to 1.62)</td><td>0.66 (0.55 to 0.80)</td><td>0.82 (0.69 to 0.98)</td></tr></table><br>Model adjusted for separation, long-term health condition, education, age, household structure.</td><td>No: compared with those in insecure employment those who are unemployed are more likely to report that they smoke however, this difference is not statistically significant.<br><br>No: Compared with those in insecure employment those who are unemployed are less likely to report that they drink but this difference is not statistically significant<br><br>No: Compared with those in insecure employment those who are unemployed are less likely to report risky drinking but this difference is not statistically significant</td></tr>
<tr><td>Butterworth P et al. 2013<br><br>Cross sectional</td><td>English Adult Psychiatric Morbidity Survey, 2007<br><br>21 to 54 years</td><td>Psychosocial job quality; demands, security, esteem from colleagues, clients, customers and managers, job control<br><br>Common mental disorders</td><td>Odds ratios and 95% confidence intervals. Relationship between employment circumstances and common mental disorders (table 3, page 810)<br><br><table><tr><td>Employed (optimal)</td><td>0.16 ( 0.10 to 0.26) p<0.001</td></tr><tr><td>One adversity</td><td>0.28 (0.18 to 0.43) p<0.001</td></tr><tr><td>Two adversities</td><td>0.58 (0.36 to 0.92) p=0.022</td></tr><tr><td>Poorest quality jobs</td><td>1.00 reference</td></tr><tr><td>Unemployed</td><td>0.79 (0.41 to 1.54) p=0.492</td></tr></table><br>Model adjusted for demographic and socioeconomic measures (age, sex, partner status, debt and housing tenure)</td><td>No: compared to those in the poorest quality jobs those who were unemployed were less likely to report common mental disorders but this difference is not statistically significant</td></tr>
<tr><td>Butterworth P et al. 2011<br><br>Cohort</td><td>Household, Income and Labour Dynamics in Australia 2001 to 2007<br><br>20 to 55 years at wave 1</td><td>Psychosocial job quality; demand, complexity, control, security, hours and shift work<br><br>Mental health</td><td>Coefficients (and SEs) from a series of longitudinal random-intercept regression models assessing the relationship between employment circumstances and mental health (table 3, page 810)<br><br><table><tr><th>Employment continuum</th><th>Model A</th><th>Model B<br>Add<br>covariates</th><th>Model C<br>Add<br>Neuroticism</th><th>Model D<br>Add<br>between-person</th><th>Model E<br>Lagged predictors<br>of change in MH</th></tr><tr><td>Optimal jobs</td><td>5.95 (0.33)***</td><td>5.77 (0.33)***</td><td>5.40 (0.35)***</td><td>5.04 (0.34)***</td><td>6.27 (0.79)***</td></tr><tr><td>1 Adversity</td><td>4.10 (0.32)***</td><td>4.00 (0.32)***</td><td>3.84 (0.34)***</td><td>3.38 (0.33)***</td><td>4.26 (0.78)***</td></tr><tr><td>2 Adversities</td><td>2.42 (0.33)***</td><td>2.32 (0.33)***</td><td>2.40 (0.35)***</td><td>1.85 (0.33)***</td><td>2.89 (0.83)***</td></tr><tr><td>Poorest jobs</td><td>Ref</td><td>Ref</td><td>Ref</td><td>Ref</td><td>Ref</td></tr><tr><td>Unemployment</td><td>-0.07 (0.55)</td><td>1.36 (0.55)*</td><td>1.61 (0.58)**</td><td>1.3 (0.56)*</td><td>3.09 (1.32)*</td></tr></table><br>*p<0.05 **p<0.01 ***p<0.001<br><br>Model B covariates included age, sex, partnered status, physical disability, % of post education life in paid employment, educational qualifications, experience of financial hardship and residence in a socially disadvantaged area<br><br>Model C incorporated neuroticism because negative affect may influence reports of job quality and mental health; this measure was only collected on one occasion<br><br>Model D incorporated experience of poorest job quality, unemployment or not in labour force<br><br>Model E looked at the effect of employment conditions on mental health on lagged employed circumstances and covariates - data from 12 months previous was used to predict change in mental health over 12 months - it only included those who did not change employment circumstance between adjacent waves</td><td>No: Those in the poorest quality jobs showed significantly greater decline in their mental health overtime than those who were unemployed</td></tr>
</table>

<table>
<tr><th>Reference/<br>Study design</th><th>Data source/<br>Population range</th><th>Poor/bad job definition<br>Outcome</th><th>Results from paper</th><th>Is any job better than no job?<br>Where OR has been calculated by OES exposure is bad/poor job, is unadjusted and data used to calculate may have been derived from percentages in study results tables</th></tr>
<tr><td>Chandola and Zhang 2018<br><br>Cohort</td><td>Understanding Society, UK Household Longitudinal Survey 2009 to 2012<br><br>35 to 75 years</td><td>Job quality; earnings, security, quality of working environment<br><br>Allostatic load – biomarkers related to chronic stress. Physical and mental wellbeing</td><td>Associations between job adversity, allostatic load and self-reported health, multiple regression coefficients at wave 3. Reference remained unemployed β coefficients and 95% confidence interval (table 3, page 54)<br><br>Good quality job One adverse measure At least two adverse measures Overall p-value<br>Allostatic load -0.387 (-1.003 to 0.230) -0.262 (-0.476 to -0.047) 0.512 (0.320 to 0.706) <0.001<br>SF-12 PCS -0.701 (-4.611 to 3.209) - 0.490 (-1.671 to 0.691) 1.914 (-3.599 to 7.426) 0.784<br>SF-12 MCS 5.541 (-2.841 to 13.923) 3.103 (0.966 to 5.240) 2.299 (-2.406 to 7.005) 0.035<br><br>Adjusted for age, gender, highest qualification, housing tenure, marital status, BMI has CVD/diabetes or not SF 12 PHS and MHS composite scores, GHQ-12, number medicines prescribed, log transformation of household income, race/ethnicity, number of children in household, number of people in household and year of latest employment</td><td>No: those who transitioned into a poor quality job had overall significantly higher levels of allostatic load than those who remained unemployed.<br><br>No: there was no difference in mental component score between those who transitioned into poor quality work and those who remained unemployed.<br><br>No: those who transitioned into a poor quality jobs (at least two adverse measures) were more likely to have a GHQ 12 score indicating distress than those who remained unemployed but this difference is not statistically significant<br><table><tr><th></th><th>GHQ 12 score distressed</th><th>GHQ 12 score not-distressed</th></tr><tr><td>Job with at least 2 adverse measures</td><td>47</td><td>79</td></tr><tr><td>Remained unemployed</td><td>19</td><td>45</td></tr></table>Relative risk 1.25 95% CI 0.80 to 1.96, p=0.3089</td></tr>
<tr><td>Cortes-Franch T et al. 2019<br><br>Cross sectional</td><td>European Social Survey 2010<br><br>16 to 64 years</td><td>Low quality job; earnings, prospects, intrinsic job quality, working time quality, participation and representation<br><br>Mental wellbeing</td><td>Odds ratios and 95% confidence intervals for the association of employment situation with poor mental wellbeing by sex and country group. UK and Ireland are liberal countries. Unemployed non-active is those who would like a job but are not actively looking for one. (table 4, page 8)<br><br>Men<br>Conservative Liberal Eastern European Southern European Social-Democratic<br>(n=3543) (n=1343) (n=1690) (n=1354) (n=394)<br>Unemployed - non-active 1 1 1 1 1<br>Unemployed-active 0.60 (0.32 to 1.15) 0.51 (0.23 to 1.12) 1.07 (0.57 to 2.01) 1.57 (0.75 to 3.25) 0.82 (0.13 to 5.22)<br>Low quality job 0.46 (0.25 to 0.83)* 0.53 (0.26 to 1.10) 0.73 (0.41 to 1.29) 0.68 (0.33 to 1.43) 1.26 (0.24 to 6.59)<br><br>Women<br>Unemployed - non-active 1 1 1 1 1<br>Unemployed - active 1.43 (0.74 to 2.75) 1.58 (0.59 to 4.27) 0.97 (0.49 to 1.92) 0.85 (0.42 to 1.72) 0.79 (0.07 to 9.69)<br>Low quality job 1.18 (0.66 to 2.13) 0.79 (0.32 to 1.92) 0.60 (0.33 to 1.09) 0.80 (0.42 to 1.54) 0.64 (0.06 to 6.59)<br>*p<0.05<br><br>Adjusted for age, marital status, job category and negative affectivity</td><td>For liberal countries (UK and Ireland)<br>No: Men in low quality jobs were slightly more likely to report poor mental wellbeing than men who were unemployed but the difference is not statistically significant<br><br>Yes: Women who were unemployed were significantly more likely to report poor mental wellbeing than those in low quality jobs<br><br>Odds ratio calculated using data from table 3, page 7 and table 4 page 8.<br><table><tr><th>Males (n=1343)</th><th>Poor mental wellbeing</th><th>No poor mental wellbeing</th></tr><tr><td>Low quality job (33.3% n= 447</td><td>(23.6%) 105</td><td>342</td></tr><tr><td>Unemployed active (10.7% n=143)</td><td>(21.5%) 31</td><td>112</td></tr></table>Odds ratio 1.11, 95% CI 0.70 to 1.75 p=0.6544<br><table><tr><th>Females (n=1198)</th><th>Poor mental wellbeing</th><th>No poor mental wellbeing</th></tr><tr><td>Low quality job (31.6% n= 379)</td><td>(34.7%) 132</td><td>247</td></tr><tr><td>Unemployed active (6.3% n=75)</td><td>(53.8%) 40</td><td>35</td></tr></table>Odds ratio 0.47 95% CI 0.28 to 0.77 p=0.0029</td></tr>
</table>

| Reference/<br>Study design | Data source/<br>Population range | Poor/bad job definition<br>Outcome | Results from paper | Is any job better than no job?<br>Where OR has been calculated by OES exposure is bad/poor job, is unadjusted and data used to calculate may have been derived from percentages in study results tables |
|---|---|---|---|---|
| Cortes-Franch T et al. 2018<br><br>Cross sectional | Spanish National Health Survey 2006 to 2007<br><br>25 to 64 years | Insecure employment, temporary or no contract<br><br>Mental health | Odds ratios and 95% confidence intervals. Association between mental health status and employment stability by sex. (table 2, page 4)<br><br>Men (n=6972) / Women (n=5307)<br>Permanent civil servant: 1 / 1<br>Temporary contract: 1.63 (1.17 to 2.26)** / 1.36 (1.0 to 1.85)*<br>No contract: 2.10 (1.13 to 3.90)* / 1.85 (1.28 to 2.67)**<br>Unemployment = or < 2 years: 3.76 (2.68 to 5.29)*** / 1.98 (1.44 to 2.74)***<br>Unemployment > 2 years: 6.76 (4.08 to 11.19)*** / 1.79 (1.20 to 2.68)**<br><br>* p<0.05 **p<0.01 ***p<0.001<br><br>Adjusted for age and social class<br><br>Note that there are inconsistencies in the number of reported participants across the tables in this paper – all tables give different totals, this is not explained in the text. This may be an error but there is no correction or comment on this in the online journal | Yes: Men who were employed in a temporary contract were less likely to report poor mental health status than those who were unemployed (for any length of time). This is likely to be significant.<br><br>Yes: Men who were employed with no contract were less likely to report poor mental health than those who had been unemployed for two years or less. This is likely to be significant.<br><br>No: Men who were employed with no contract were less likely to report poor mental health than those who had been unemployed for more than two years. This is unlikely to be significant.<br><br>No: Women who were employed with no, or a temporary contract were less likely to report poor mental health than women who had been unemployed for any length of time. This difference is unlikely to be statistically significant. |
| Fiori et al. 2016<br><br>Cross sectional | Health Conditions and Access to Health Services Survey, Italy, 2005 and 2013<br><br>18 to 39 years | Insecure employment<br><br>Mental health | Effects of status in the labour market on mental health score, 2013 unstandardised β coefficients (table 1, page 93)<br><br>Model 1a status in labour market<br>Men / Women / Men/women<br>Permanent employment: ref / ref<br>Fixed term employment: 1.684** / 2.547***<br>Atypical employment: 2.763* / 2.614*<br>In search of new job: 7.798*** / 4.765*** / ***<br>In search of first job: 5.053*** / 2.392** / **<br><br>Model 1b status in the labour market + occupational status<br>Permanent employment medium-high: ref / ref<br>Fixed term employment medium high: 2.193** / 1.833*<br>Fixed term low: 2.278** / 3.747***<br>Atypical contract: 3.473** / 2.806*<br>In search of a new job: 8.590*** / 5.047*** / ***<br>In search of first job: 5.830*** / 2.675***<br><br>Models 1a and b adjusted for age, area of residence, living arrangement and educational qualification<br><br>Effects of status in labour market and survey year on mental health score by gender, pooled data for the years 2005 and 2013 unstandardised β coefficients (table 5, page 95)<br>Men / Women<br>Permanent employment: ref / ref<br>Fixed term employment: 2.197*** / 1.947***<br>Atypical contract: 2.541*** / 2.323***<br>In search of a new job: 7.543*** / 3.960***<br>In search of a first job: 4.535*** / 1.284**<br><br>Adjusted for age, area of residence, living arrangement and educational qualifications<br><br>*p<0.05 **p<0.01 ***p<0.001 | Yes: Those in fixed term employment and those with an atypical contract were less likely to report poor mental health than those who were looking for a new job. Is isn't possible to tell if these differences are likely to be statistically significant |

| Reference/<br>Study design | Data source/<br>Population range | Poor/bad job definition<br>Outcome | Results from paper | Is any job better than no job?<br>Where OR has been calculated by OES exposure is bad/poor job, is unadjusted and data used to calculate may have been derived from percentages in study results tables |
|---|---|---|---|---|
| | | | Positive estimated β coefficients indicate worse mental health compared with respondents in the reference category | |
| Flint et al. 2013<br><br>Cohort | British Household Panel Survey<br>1991 to 2007<br><br>16-65 years | Insecure employment<br><br>Psychological wellbeing | Association between labour market status categories and GHQ-12 from multivariate fixed affects model beta coefficient and 95% confidence interval (table 2, page 799)<br><br>Securely employed 0<br>Insecurely employed 1.11 (1.00 to 1.21)<br>Unemployed 2.21 (1.99 to 2.43)<br><br>Adjusted for age, educational attainment, physical health problems, spousal joblessness, spousal GHQ-12 caseness, marital status, unemployed spells in the last 12 month, residence in social housing, substance abuse and equivalised household income. | Yes: Compared to when they were securely employed an individual experiencing a spell of insecure employment had a GHQ-12 score elevated by 1.2 units. An unemployed spell was associated with an increase of 2.2 units. Higher scores indicate lower levels of psychological wellbeing. This difference is probably significant |
| Fornell B et al. 2018<br><br>Cross sectional | Survey of Living Conditions, Spain, 2007 to 2011<br><br>16 to 65 years | Insecure employment<br><br>General health | Odds ratios and 95% confidence intervals association between perceived health, unemployment, and employment precariousness. Does not state but implies reference is permanent employment<br><br>Unemployed 1.7484 (1.5274 to 2.0013) $p<0.001$<br>Precariousness 1.3755 (1.1902 to 1.5903) $p<0.001$<br><br>Unclear but analysis appears to be unadjusted | No: Those who were unemployed were more likely to report that their health was bad than those in precarious employment. This difference is unlikely to be statistically significant |
| Gebel and Vosemer 2014<br><br>Case control | German Socio-Economic Panel<br>1995 to 2010<br><br>16 to 54 years | Insecure employment<br><br>Psychological and physical health | Conditional average treatment effects on the treated. ATT is average treatment effect on the treated (health effect of making the transition) and standard error (table 3, page 134)<br><br>Psychological health: ATT, S.E, $N_t/N_c$; Physical health: ATT, S.E, $N_t$/Nc<br><br>Transition into unemployment<br>Fixed term contract at *t* -0.72 0.11 750/2875 0.09 0.12 772/2875<br><br>Permanent contract at *t* -0.81 0.06 1871/73,125 -0.05 0.06 1816/73,125<br><br>Transition out of unemployment<br>Fixed term contract at *t*+1 0.96 0.09 990/4573 0.09 0.09 990/4573<br><br>Permanent contract at t+1 1.05 0.09 1466/4573 0.02 0.09 1465/4573 | No: There was no significant impact on the **physical health** of temporary workers who became unemployed<br><br>Yes: There was a significant negative impact on the **psychological health** of temporary workers who became unemployed |
| Griep Y et al. 2016<br><br>Cross sectional | Living Conditions Survey, Finland, 1994<br><br>18 to 64 years | Perceived job insecurity<br><br>Psychological complaints, self-rated health, life satisfaction | Estimated means (with standard errors) for outcome variables for the employment status groups - paired comparisons with Bonferroni test<br><br>Secure permanent (n=2257); Insecure permanent (n=713); Short-term unemployed (n=662); Long-term unemployed (n=345); F; Partial eta squared<br><br>Psychological complaints 1.33 (0.01) $^{<2}$ 1.42 (0.01) $^{<1.3}$ 1.35 (0.02) $^{<2,4}$ 1.42 (0.02) $^{>3}$ (3,3961) = 18.95*** 0.014<br><br>Subjective complaints 1.23 (0.01) $^{<2,4}$ 1.27 (0.01) $^{>1}$ 1.25 (0.01) 1.28 (0.01) $^{>1}$ (3, 3962) = 10.75*** 0.008<br><br>Self-rated health 3.98 (0.03) $^{<2,4}$ 3.89 (0.03) $^{<1}$ 3.94 (0.03) $^{>4}$ 3.74 (0.04) $^{<1,3}$ (3, 3959) =7.72*** 0.006<br><br>Life satisfaction 3.24 (0.02) $^{>2,3,4}$ 3.13 (0.02) $^{<1; >3,4}$ 3.00 (0.03) $^{>4; <1,2}$ 2.86 (0.03) $^{<1,2,3}$ (3, 3956) = 61.98*** 0.045<br><br>Adjusted for gender, age, education, marital status, children less than 18 years old living at home, income, type of living area and long-term illness/injury. In the pairwise comparisons for the | No: Insecure permanent employees reported significantly more **psychological complaints** than short term unemployed individuals; there was no difference between insecure permanent employees and those who were long-term unemployed<br><br>Insecure permanent employees reported more **subjective complaints** than the long-term unemployed but fewer than the short-term unemployed<br><br>Insecure permanent employees rated their **health** as better than the long-term unemployed but worse than the short-term unemployed<br><br>Yes: Insecure permanent employees reported significantly higher **life satisfaction** than both the short and long-term unemployed |

<table>
<tr><th>Reference/<br>Study design</th><th>Data source/<br>Population range</th><th>Poor/bad job definition<br>Outcome</th><th>Results from paper</th><th>Is any job better than no job?<br>Where OR has been calculated by OES exposure is bad/poor job, is unadjusted and data used to calculate may have been derived from percentages in study results tables</th></tr>
<tr><td></td><td></td><td></td><td>significance level test used Bonferroni adjustment for multiple comparisons. Only significant pairwise comparisons are reported ($p<0.05$), The superscript numbers indicate for which between-classes comparisons the means differ significantly ($p<0.05$) The < or > preceding the superscript numbers indicate the direction of the significance difference in means.<br>*$p<0.05$, **$p<0.01$, ***$p<0.001$</td><td></td></tr>
<tr><td>Grzywacz JG and Dooley D 2003<br><br>Cross sectional</td><td>National Survey of Midlife Development, USA, 1995<br><br>25 to 74 years</td><td>Inadequate employment; low pay and psychological aspects<br><br>Physical health, depression</td><td>Logistic regression results estimating the association between employment status and poor/fair physical health (unweighted data).<br><br><table><tr><th>Employment status</th><th>B</th><th>SE (B)</th><th>odds ratio</th></tr><tr><td>Optimal employment</td><td>Reference</td><td></td><td></td></tr><tr><td>Barely adequate</td><td>0.45*</td><td>0.26</td><td>1.57</td></tr><tr><td>Inadequate</td><td>0.59**</td><td>0.26</td><td>1.80</td></tr><tr><td>Unemployed</td><td>1.81****</td><td>0.31</td><td>6.14</td></tr></table><br>* $p \le 0.10$<br>** $p \le 0.05$<br>*** $p \le 0.01$<br>**** $p \le 0.001$ (two tailed)<br><br>Adjusted for sociodemographic characteristics<br><br>Logistic regression estimating the association between employment status and depression<br><br><table><tr><th>Employment status</th><th>B</th><th>SE (B)</th><th>odds ratio</th></tr><tr><td>Optimal employment</td><td>Reference</td><td></td><td></td></tr><tr><td>Barely adequate</td><td>0.21</td><td>0.23</td><td>1.24</td></tr><tr><td>Inadequate</td><td>0.72***</td><td>0.23</td><td>2.06</td></tr><tr><td>Unemployed</td><td>1.22****</td><td>0.30</td><td>3.37</td></tr></table><br>Adjusted for age, gender, race/ethnicity, and socioeconomic status<br><br>We did not include data from the California Work and Health Survey because the study population was not nationally representative</td><td>Yes: those who were in inadequate employment were significantly less likely to report poor/fair health than those who were unemployed.<br><br>No: those who were in inadequate employment were less likely to meet the criteria for depression than those who were unemployed. However, this difference is not statistically significant<br><br>Text states that there were 3032 participants in MIDUS. Footnotes from table 1 say 21 participants excluded from MIDUS so assume overall participants 3011. Table 2b gives % from MIDUS sample in the different employment categories.<br><br><table><tr><td>Total MIDUS</td><td>3011</td></tr><tr><td>Inadequate employment</td><td>(15.4%) 464</td></tr><tr><td>Unemployed</td><td>(4.1%) 123</td></tr></table><br>From Fig 1 page 1756<br><table><tr><td>Fair/poor health</td><td></td></tr><tr><td>Inadequate employment</td><td>(17%) 79</td></tr><tr><td>Unemployed</td><td>(31.5%) 39</td></tr></table><br><table><tr><td></td><td>Fair/poor health</td><td>Good health</td></tr><tr><td>Inadequate employment</td><td>79</td><td>385</td></tr><tr><td>Unemployed</td><td>39</td><td>84</td></tr></table>Odds ratio 0.44, 95% CI 0.28 to 0.69 p=0.0004<br><br>From fig 2 page 1756<br><table><tr><td>Depression</td><td></td></tr><tr><td>Inadequate employment</td><td>(18.6%) 86</td></tr><tr><td>Unemployed</td><td>(23.9%) 29</td></tr></table><br><table><tr><td></td><td>Depression</td><td>No depression</td></tr><tr><td>Inadequate employment</td><td>86</td><td>378</td></tr><tr><td>Unemployed</td><td>29</td><td>94</td></tr></table>Odds ratio 0.73, 95% CI 0.46 to 1.19 p=0.2114</td></tr>
<tr><td>Guseva Canu et al. 2019<br><br>Cohort</td><td>Swiss census data 1990 to 2004<br><br>18 to 65 years</td><td>Occupational status lowest skilled and unskilled employees<br><br>Suicide</td><td>Mortality from suicide in Swiss working-age population (1990-2014) by selected socio-economic factors and by sex: Swiss National Cohort (1990-2014) (table 2)<br><table><tr><th></th><th>Men SMR</th><th>Men DSR</th><th>Women SMR</th><th>Women DSR</th></tr><tr><td>Unskilled employees and workers</td><td>0.90 (0.85 to 0.95) (1450 deaths)</td><td>26.25 (24.73 to 27.77)</td><td>0.88 (0.81 to 0.96) (559 deaths)</td><td>9.57 (8.59 to 10.56)</td></tr><tr><td>Unemployed /job seeking</td><td>1.95 (1.80 to 2.12)</td><td>52.94 (46.32 to 59.56)</td><td>2.11 (1.87 to 2.37)</td><td>21.84 (16.85 to 26.84)</td></tr></table></td><td>Yes: Unskilled employees and workers were at significantly lower risk of dying by suicide than those who were unemployed.<br><br>Data from table 1, page 1486<br><table><tr><td>Men</td><td>Suicide</td><td>No suicide</td></tr><tr><td>Unskilled employees and</td><td>1450</td><td>281,718</td></tr></table></td></tr>
</table>

<table>
<tr><th>Reference/<br>Study design</th><th>Data source/<br>Population range</th><th>Poor/bad job definition<br>Outcome</th><th>Results from paper</th><th>Is any job better than no job?<br>Where OR has been calculated by OES exposure is bad/poor job, is unadjusted and data used to calculate may have been derived from percentages in study results tables</th></tr>
<tr><td></td><td></td><td></td><td>(568 deaths) (281 deaths)</td><td><table>
<tr><td>workers<br>(n=283,168)</td><td></td><td></td></tr>
<tr><td>Unemployed/job seeking<br>(n=77,181)</td><td>568</td><td>76,613</td></tr>
</table>
Relative risk 0.69, 95% CI 0.63 to 0.77, p<0.0001
<table>
<tr><td>Women</td><td>Suicide</td><td>No suicide</td></tr>
<tr><td>Unskilled employees and workers<br>(n=264,046)</td><td>559</td><td>263,487</td></tr>
<tr><td>Unemployed/job seeking<br>(n=90,812)</td><td>281</td><td>90,531</td></tr>
</table>
Relative risk 0.68, 95% CI 0.59 to 0.79, p<0.0001</td></tr>
<tr><td>Inanc H 2018<br>(see also Inanc H 2016)<br>Cross sectional</td><td>British Household Panel Survey, 8 waves 1991 onwards<br><br>20 to 65 years</td><td>Insecure employment<br><br>Psychological wellbeing and life satisfaction</td><td>Wives subjective wellbeing as a function of husbands' labour market insecurity, fixed effects regressions. Coefficients and assume standard error – tables in paper not labelled<br><br>
<table>
<tr><td>Husband<br>Temporary vs unemployed</td><td>Psychological wellbeing (n=7420)</td><td>Life satisfaction (n=6486)</td></tr>
<tr><td></td><td>-0.006 (0.005)</td><td>-0.002 (0.007)</td></tr>
<tr><td>Self</td><td>0.043 (0.005)***</td><td>0.026 (0.008)***</td></tr>
</table>
Reference is unemployed<br><br>***p<0.001<br><br>Husbands' subjective well-being as a function of wives labour market insecurity. Fixed effects regressions. Coefficients and assume standard error – tables in paper not labelled<br><br>
<table>
<tr><td>Wife</td><td>(n=7175)</td><td>(n=6240)</td></tr>
<tr><td>Temporary vs unemployed</td><td>-0.010 (0.005)**</td><td>-0.009 (0.007)</td></tr>
<tr><td>Self</td><td>0.047 (0.004)***</td><td>0.030 (0.007)***</td></tr>
</table>
**p<0.01 ***p<0.001<br><br>Adjusted for partners respective wellbeing and life satisfaction, regional unemployment rate, current duration of partnership, number and presence of children, married or cohabiting, age, year of interview</td><td>No: A husbands temporary employment had a greater negative effect on their wife's psychological wellbeing and life satisfaction than if he were unemployed.<br><br>Yes: However, a husband's own psychological wellbeing and life satisfaction was significantly greater when they were in temporary employment, compared to being unemployed.<br><br>No: A wife's temporary employment had a significantly greater negative effect on their husband's psychological wellbeing and but not on their life satisfaction than if she were unemployed.<br><br>Yes: However, a wife's own psychological wellbeing and life satisfaction was greater when they were in temporary employment, compared to being unemployed.</td></tr>
<tr><td>Jang et al. 2015<br><br>Cohort</td><td>Korean Welfare Panel Study 2007 to 2013<br><br>19 to 65 years</td><td>Insecure employment<br><br>Severe depressive symptoms</td><td>Generalised estimating equation analysing the effect of yearly employment status on new-onset of severe depressive symptoms during the 6-year follow-up period, among individuals who were waged workers at baseline in 2007. Odds ratios and 95% confidence intervals. Reference is full time permanent (table 2, page 334)<br><br>
<table>
<tr><td>Employment status each year from 2008</td><td>Males</td><td>Females</td></tr>
<tr><td>Unemployed</td><td>2.90 (1.85 to 4.57)<br>(33 cases)</td><td>1.85 (1.17 to 2.90)<br>(46 cases)</td></tr>
<tr><td>Precarious</td><td>1.59 (1.10 to 2.30)<br>(63 cases)</td><td>1.41 (0.94 to 2.12)<br>(70 cases)</td></tr>
</table></td><td>Yes: in workers who were employed at baseline, those who moved into precarious employment were at significantly lower risk of developing severe depressive symptoms than those who became unemployed.<br>From table 1, page 333
<table>
<tr><td></td><td>Precarious</td><td>Unemployed</td></tr>
<tr><td>Males<br>(n=2214)</td><td>(18.7%) 414</td><td>(3.3%) 73</td></tr>
<tr><td>Females<br>(n=1276)</td><td>(28.5%) 364</td><td>(13.1%) 167</td></tr>
</table>
Table 2 for number of cases with depressive symptoms
<table>
<tr><td>Males</td><td>Depressive symptoms</td><td>No symptoms</td></tr>
</table></td></tr>
</table>

| Reference/ Study design | Data source/ Population range | Poor/bad job definition Outcome | Results from paper | Is any job better than no job? Where OR has been calculated by OES exposure is bad/poor job, is unadjusted and data used to calculate may have been derived from percentages in study results tables |
|---|---|---|---|---|
| | | | Generalised estimating equation analysing the effect of yearly employment and new-onset of severe depressive symptoms during 6 year follow up period among individuals who were waged workers at baseline in 2007. Odds ratios and 95% confidence intervals. Reference is full time permanent<br><br>Head of household: Males / Females; Non-head of household: Males / Females<br>Employment status each year<br><br>Unemployed — Head of household: Males 3.10 (1.88 to 5.13); Females 6.10 (2.17 to 17.12) (36 cases); Non-head of household: Males 2.69 (0.93 to 7.76) (43 cases); Females 1.31 (0.78 to 2.18)<br>Precarious — Head of household: Males 1.52 (1.02 to 2.25); Females 4.19 (1.70 to 10.32) (85 cases); Non-head of household: Males 2.13 (0.71 to 6.34) (48 cases); Females 1.03 (0.63 to 1.68)<br><br>Adjusted for previous year's CES-D 11 score, age in years, equalized household income, education level, head of household status, marital status, residential area, occupation at baseline, company size at baseline, exposure to hazardous environment in workplace, chronic disease, perceived health status, smoking status, alcohol use, and observation year | Precarious \| 63 \| 351<br>Unemployed \| 33 \| 40<br>Relative risk 0.34, 95% CI 0.24 to 0.47, p<0.001<br>Females \| Depressive symptoms \| No symptoms<br>Precarious \| 70 \| 294<br>Unemployed \| 46 \| 121<br>Relative risk 0.70, 95% CI 0.51 to 0.97, p=0.0296 |
| Kim HD, Park SG 2021<br><br>Cohort<br><br>This study not included in synthesis | Korean Welfare Panel Study 2017 to 2018<br><br>19 to 59 years | Precarious employment<br><br>New onset depressive symptoms | Association between employment status change and new-onset depressive symptoms in permanent waged workers at baseline (2017), follow up 2018. Odds ratio and 95% confidence interval (table 2, page 111)<br><br>Males / Females<br>Permanent: Ref / Ref<br>Precarious: 3.15 (1.30 to 7.65) / 1.99 (0.75 to 5.25)<br>Unemployed: 4.50 (1.19 to 17.06) / 5.12 ( 0.88 to 29.75)<br><br>Adjusted for age, religion, education, marital status, head of household, self-rated health | No: Those who transitioned from permanent to precarious employment were less likely to report new-onset depressive symptoms than those who transitioned from permanent employment to unemployment but the difference is not statistically significant |
| Kim SS et al. 2012<br><br>Cohort<br><br>This study not included in synthesis | Korean Welfare Panel Study 2007 to 2009<br><br>18 years and above | Insecure employment<br><br>New onset depressive symptoms | Association between change in employment status and new-onset depressive symptoms (2008). Results for sub population - this excludes those with a disability or chronic disease and without depressive symptoms in 2006 and 2007. Odds ratios and 95% confidence intervals. Employment status baseline 2007, follow up 2008 (n= 2021) (table 3, page 541)<br><br>Baseline permanent<br>Follow up permanent 1.0<br>Follow up precarious 1.66 (0.97 to 2.84)<br>Follow up unemployed 0.57 (0.80 to 4.05)<br><br>Baseline precarious<br>Follow up precarious 1.44 (0.94 to 2.23)<br>Follow up permanent 1.22 (0.75 to 1.96)<br>Follow up unemployed 2.67 (0.84 to 8.40)<br><br>Gender-specific association between change in employment status and new-onset depressive symptoms (2008) fully adjusted model. Baseline 2007, follow up 2008. Odds ratios and 95% confidence intervals (table 4, page 541)<br><br>Male (n= 1296) / Female (n=725)<br>Baseline permanent<br>Follow up permanent: 1.0 / 1.0<br>Follow up precarious: 1.19 (0.57 to 2.50) / 2.88 (1.24 - 6.66) p<0.05<br>Follow up unemployed: 1.54 (0.21 to 10.98) / None became depressed<br><br>Baseline precarious<br>Follow up precarious: 1.59 (0.90 to 2.81) / 1.50 (0.69 to 3.25)<br>Follow up permanent: 0.76 (0.39 to 1.46) / 2.57 (1.20 to 5.52) p<0.05<br>Follow up unemployed: 0.84 (0.10 to 7.0) / 8.01 (1.99 to 32.26 p<0.01<br><br>Adjusted for age, sex, marital status, educational level, household equivalised income, residential area and smoking status at baseline | No: In workers in precarious employment at baseline those who were unemployed at follow up were more likely to report new onset of depressive symptoms than those who remained in precarious employment but the differences were not statistically significant.<br><br>Males in precarious employment at baseline and at follow up were more likely to report new onset depressive symptoms than those who were in precarious employment at baseline but unemployed at follow up but the difference is unlikely to be statistically significant.<br><br>Females in precarious employment at baseline and follow up were less likely to report new onset depressive symptoms at follow up than those who were in precarious employment at baseline and unemployed at follow up, but the difference is unlikely to be statistically significant. |

<table>
<tr><th>Reference/<br>Study design</th><th>Data source/<br>Population range</th><th>Poor/bad job definition<br>Outcome</th><th>Results from paper</th><th>Is any job better than no job?<br>Where OR has been calculated by OES exposure is bad/poor job, is unadjusted and data used to calculate may have been derived from percentages in study results tables</th></tr>
<tr><td>Kim SS et al. 2013<br><br>Cohort<br><br>This study not included in synthesis</td><td>Korean Welfare Panel Study 2007 to 2010<br><br>18 years and above</td><td>Insecure employment<br><br>New onset depressive symptoms</td><td>Association between gain of employment and depressive symptoms at follow up (2008 or 2010) among workers who were unemployed at baseline (2007 or 2009). Full population excluded respondents who had depressive symptoms at baseline (2007 or 2009); subpopulation excluded respondents who depressive symptoms in any of the two prior years (2006/2007 or 2008/2009). Rate ratios and 95% confidence intervals (table 2)<br><br>
<table>
<tr><th></th><th>Full population (n=308)</th><th></th><th>Subpopulation (n=249)</th><th></th></tr>
<tr><td>Unemployed</td><td>n= 88</td><td>1</td><td>n= 66</td><td>1</td></tr>
<tr><td>Part-time precarious</td><td>n= 26</td><td>0.51 (0.15 to 1.76)</td><td>n= 20</td><td>0.87 (0.21 to 3.60)</td></tr>
<tr><td>Full time precarious</td><td>n= 105</td><td>0.38 (0.18 to 0.83) p<0.05</td><td>n= 91</td><td>0.49 (0.19 to 1.25)</td></tr>
<tr><td>Full time permanent</td><td>n= 89</td><td>0.26 (0.11 to 0.63) p<0.001</td><td>n= 72</td><td>0.27 (0.08 to 0.86) p<0.05</td></tr>
</table>
<br>Adjusted for age, sex, education, marital status, residential area, household income, having a chronic disease, having a disability, smoking at baseline (2007 or 2009)</td><td>No: in workers in permanent employment at baseline those who were in precarious employment at follow up were more likely to report new onset of depressive symptoms than those who were unemployed but the difference was not significant. In workers in precarious employment at baseline those who were unemployed at follow up were more likely to report new onset of depressive symptoms than those who remained in precarious employment but the differences were not significant</td></tr>
<tr><td>Kim W et al. 2019<br><br>Cohort<br><br>Same as Yoon</td><td>Korean Welfare Panel Study 2012 to 2017<br><br>25 to 59 years</td><td>Insecure employment<br><br>Suicidal ideation in the past year</td><td>Employment factors associated with suicide ideation. Odds ratios and 95% confidence intervals. Reference is remaining in permanent employment<br><br>Employment transition<br>
<table>
<tr><td>Precarious to precarious</td><td>1.86 (1.21 to 2.85)</td></tr>
<tr><td>Precarious to unemployment</td><td>1.43 (1.05 to 1.95)</td></tr>
<tr><td>Unemployment to precarious</td><td>1.11 (0.49 to 2.50)</td></tr>
<tr><td>Unemployment to unemployment</td><td>1.57 (0.86 to 2.87)</td></tr>
</table>
<br>Adjusted for sex, age, region, education level, marital status, income, disability, chronic disease, depression and year</td><td>Yes: Those who remained in precarious employment were significantly less likely to report suicide ideation than those who remained unemployed.<br><br>Yes: Those who remained in precarious employment were significantly less likely to report suicide ideation than those who moved from precarious employment to unemployment.<br><br>No: Those who transitioned from unemployment to precarious employment were slightly less likely to report suicide ideation than those who transitioned from precarious employment to unemployment but the difference is not statistically significant<br><br>No: Those who transitioned from unemployment to precarious employment were more likely to report suicide ideation than those who remained unemployed. However, this difference is not statistically significant<br><br>From table 1, page 5
<table>
<tr><th></th><th>Suicide ideation</th><th>No suicide ideation</th></tr>
<tr><td>Precarious to precarious (n=5166)</td><td>148</td><td>5018</td></tr>
<tr><td>Precarious to unemployment (n=953)</td><td>42</td><td>911</td></tr>
<tr><td>Unemployment to precarious (n=1161)</td><td>47</td><td>1114</td></tr>
<tr><td>Unemployment to unemployment (n=6796)</td><td>266</td><td>6503</td></tr>
</table>
<br>
<table>
<tr><th></th><th>Suicide ideation</th><th>No suicide ideation</th></tr>
<tr><td>Precarious to precarious</td><td>148</td><td>5018</td></tr>
</table></td></tr>
</table>

<table>
<tr><th>Reference/<br>Study design</th><th>Data source/<br>Population range</th><th>Poor/bad job definition<br>Outcome</th><th>Results from paper</th><th>Is any job better than no job?<br>Where OR has been calculated by OES exposure is bad/poor job, is unadjusted and data used to calculate may have been derived from percentages in study results tables</th></tr>
<tr><td></td><td></td><td></td><td></td><td>
<table>
<tr><td>Unemployment to unemployment</td><td>266</td><td>6503</td></tr>
</table>
Relative risk 0.73, 95% CI 0.60 to 0.89, p=0.0017
<table>
<tr><th></th><th>Suicide ideation</th><th>No suicide ideation</th></tr>
<tr><td>Precarious to precarious</td><td>148</td><td>5018</td></tr>
<tr><td>Precarious to unemployment</td><td>42</td><td>911</td></tr>
</table>
Relative risk 0.65, 95% CI 0.46 to 0.91, p=0.0119
<table>
<tr><th></th><th>Suicide ideation</th><th>No suicide ideation</th></tr>
<tr><td>Unemployment to precarious</td><td>47</td><td>1114</td></tr>
<tr><td>Precarious to unemployment</td><td>42</td><td>911</td></tr>
</table>
Relative risk 0.92, 95% CI 0.61 to 1.38, p=0.6827
<table>
<tr><th></th><th>Suicide ideation</th><th>No suicide ideation</th></tr>
<tr><td>Unemployment to precarious</td><td>47</td><td>1114</td></tr>
<tr><td>Unemployment to unemployment</td><td>266</td><td>6503</td></tr>
</table>
Relative risk 1.03, 95% CI 0.76 to 1.40, p=0.8479
</td></tr>
<tr><td>Kim Y et al. 2020<br><br>Cross sectional</td><td>Korean National Health and Nutrition Examination Survey, 2015<br><br>19 to 65 years</td><td>Precarious employment – not further defined<br><br>C-reactive protein (inflammatory marker)</td><td>Association between occupational class and high sensitivity C-reactive protein. Beta coefficient and 95% confidence intervals. C reactive protein (log-transformed, mg/l) model 4 (table 2, page 198)<br><br>
<table>
<tr><td>Blue collar workers</td><td>Ref</td></tr>
<tr><td>Unemployed</td><td>0.14 (-0.04 to 0.31)</td></tr>
<tr><td>Precarious work</td><td>0.05 (-0.04 to 0.14)</td></tr>
</table>
Adjusted for age, sex, education, religion, household income, marital status, work-related characteristics (weekly hours, shift work), history of severe chronic condition, smoking, drinking, physical activity, perceived stress and presence of metabolic syndrome and BMI class</td><td>No: Those in precarious employment had lower levels of C-reactive protein than those who were unemployed but this difference is not statistically significant</td></tr>
<tr><td>Maeda et al. 2019<br><br>Cross sectional</td><td>Comprehensive Survey of Living Conditions, Japan, 2010<br><br>20 to 59 years</td><td>Not regularly employed – not further defined<br><br>Insomnia related symptoms</td><td>Multivariate odds ratios and 95% confidence intervals for insomnia related symptoms Reference is regularly employed<br><br>
<table>
<tr><th></th><th>Unemployed</th><th>Non regularly employed</th></tr>
<tr><td>Men</td><td>2.5 (1.8 to 3.4) n=1,075, with symptoms 71</td><td>1.1 (0.8 to 1.6) n=2,083 with symptoms 38</td></tr>
<tr><td>Women</td><td>1.9 (1.5 to 2.5) n=3483 with symptoms 148</td><td>1.1 (0.9 to 1.4) n=7,807 with symptoms 171</td></tr>
</table>
Adjusted for age, marital status, education level, mental illness, smoking status, parental status, household expenditure and insomnia related chronic diseases</td><td>Yes: Those who not regularly employed were significantly less likely to report insomnia related symptoms then those who were unemployed.<br><br>From table 2, page 706
<table>
<tr><th>Men</th><th>Insomnia symptoms</th><th>No insomnia symptoms</th></tr>
<tr><td>Not regularly employed (n=2083)</td><td>38</td><td>2045</td></tr>
<tr><td>Unemployed (n=1075)</td><td>71</td><td>1004</td></tr>
</table>
Odds ratio 0.26, 95% CI 0.18 to 0.39, p <0.0001
<table>
<tr><th>Women</th><th>Insomnia symptoms</th><th>No insomnia symptoms</th></tr>
<tr><td>Not regularly employed (n=7087)</td><td>171</td><td>6916</td></tr>
</table>
</td></tr>
</table>

<table>
<tr><th>Reference/<br>Study design</th><th>Data source/<br>Population range</th><th>Poor/bad job definition<br>Outcome</th><th>Results from paper</th><th>Is any job better than no job?<br>Where OR has been calculated by OES exposure is bad/poor job, is unadjusted and data used to calculate may have been derived from percentages in study results tables</th></tr>
<tr><td></td><td></td><td></td><td></td><td><table><tr><td>Unemployed (n=3483)</td><td>148</td><td>3335</td></tr></table>Odds ratio 0.56, 95% CI 0.44 to 0.70, p <0.001</td></tr>
<tr><td>Matilla-Santander N et al. 2020<br><br>Cross sectional</td><td>European Working Conditions Survey, 6th wave, 2015<br><br>15 years and over</td><td>Precarious employment; temporariness, not being able to exercise rights, vulnerability, disempowerment and wages<br><br>Bad health, health problems</td><td>Prevalence of health-related outcomes among highly precariously, not precariously employed and unemployed individuals in Europe in 2015 (overall quartiles % and 95% confidence intervals, unadjusted prevalence)<br><br><table>
<tr><th></th><th>Non-precarious n=13,663</th><th>Highly precarious 4th quartile n=4769</th><th>Unemployed n=253</th></tr>
<tr><td>Bad health status</td><td>20.8 (16.79 to 23.82)</td><td>24.08 (20.06 to 28.61)</td><td>33.3 (24.4 to 43.5)</td></tr>
<tr><td>Hearing problems</td><td>4.10 (3.03 to 5.54)</td><td>5.80 (4.82 to 6.96)</td><td>4.09 (1.90 to 8.55)</td></tr>
<tr><td>Skin problems</td><td>6.58 (5.28 to 8.18)</td><td>10.91 (9.07 to 13.07)</td><td>9.13 (5.21 to 15.53)</td></tr>
<tr><td>Headaches, eyestrain</td><td>37.18 (30.13 to 44.83)</td><td>44.89 (37.08 to 52.96)</td><td>36.23 (29.20 to 43.89)</td></tr>
<tr><td>Anxiety</td><td>14.21 (10.29 to 19.29)</td><td>22.19 (14.47 to 32.46)</td><td>21.14 (14.63 to 29.54)</td></tr>
<tr><td>Fatigue</td><td>35.91 (27.98 to 44.69)</td><td>48.27 (36.49 to 60.25)</td><td>38.95 (30.43 to 48.20)</td></tr>
<tr><td>Backache</td><td>41.63 (37.54 to 45.84)</td><td>52.56 (47.22 to 57.85)</td><td>47.20 (39.32 to 55.22)</td></tr>
<tr><td>Muscular pain in upper limbs</td><td>39.46 (35.40 to 44.65)</td><td>50.75 (45.12 to 56.36)</td><td>40.28 (33.07 to 47.93)</td></tr>
<tr><td>Muscular pain in lower limbs</td><td>28.92 (23.90 to 34.51)</td><td>37.46 (32.86 to 42.30</td><td>35.54 (28.94 to 42.74)</td></tr>
<tr><td>Injuries</td><td>6.58 (5.29 to 8.17)</td><td>10.83 (8.63 to 13.51)</td><td>6.18 (3.59 to 10.4)</td></tr>
</table></td><td>Yes: Those in highly precarious employment were significantly less likely to report bad health status than those who were unemployed.<br><br>No: Those in highly precarious employment were more likely to report hearing problems, skin problems, anxiety, backache and muscular pain in lower limbs. These differences are not statistically significant<br><br>No: Those in highly precarious employment were significantly more likely to report headaches/eyestrain, fatigue, muscular pain in upper limbs and injuries.<br><br>Table 2, page 7<table>
<tr><th></th><th>Bad health status</th><th>Not bad health status</th></tr>
<tr><td>Highly precarious employment (n=4769)</td><td>(24.08%) 1148</td><td>3621</td></tr>
<tr><td>Unemployed (n=253)</td><td>(33.3%) 84</td><td>169</td></tr>
</table>Odds ratio 0.64, 95% CI 0.49 to 0.84, p=0.0011<br><br><table>
<tr><th></th><th>Hearing problems</th><th>No hearing problems</th></tr>
<tr><td>Highly precarious employment (n=4769)</td><td>(5.8%) 277</td><td>4492</td></tr>
<tr><td>Unemployed (n=253)</td><td>(4.09%) 10</td><td>243</td></tr>
</table>Odds ratio 1.50, 95% CI 0.79 to 2.85, p=0.2183<br><br><table>
<tr><th></th><th>Skin problems</th><th>No skin problems</th></tr>
<tr><td>Highly precarious employment (n=4769)</td><td>(10.91%) 520</td><td>4249</td></tr>
<tr><td>Unemployed (n=253)</td><td>(9.13%) 23</td><td>230</td></tr>
</table>Odds ratio 1.22, 95% CI 0.79 to 1.90 p=0.3663<br><br><table>
<tr><th></th><th>Headaches, eye strain</th><th>No headaches, eyestrain</th></tr>
<tr><td>Highly precarious employment (n=4769)</td><td>(44.89%) 2150</td><td>2619</td></tr>
<tr><td>Unemployed (n=253)</td><td>(36.23%) 92</td><td>161</td></tr>
</table>Odds ratio 1.44, 95% CI 1.11 to 1.87 p=0.0068</td></tr>
</table>

| Reference/ Study design | Data source/ Population range | Poor/bad job definition Outcome | Results from paper | Is any job better than no job? Where OR has been calculated by OES exposure is bad/poor job, is unadjusted and data used to calculate may have been derived from percentages in study results tables |
|---|---|---|---|---|

| | Anxiety | No anxiety |
|---|---|---|
| Highly precarious employment (n=4769) | (22.19%) 1058 | 3711 |
| Unemployed (n=253) | (21.14%) 53 | 200 |

Odds ratio 1.08, 95% CI 0.78 to 1.47 p=0.6444

| | Fatigue | No fatigue |
|---|---|---|
| Highly precarious employment (n=4769) | (48.27%) 2302 | 2467 |
| Unemployed (n=253) | (38.95%) 99 | 154 |

Odds ratio 1.45, 95% CI 1.12 to 1.88, p=0.0048

| | Backache | No backache |
|---|---|---|
| Highly precarious employment (n=4769) | (52.56%) 2507 | 2262 |
| Unemployed (n=253) | (47.20%) 119 | 134 |

Odds ratio 1.25, 95% CI 0.97 to 1.60, p=0.0865

| | Muscular pain upper limbs | No muscular pain upper limbs |
|---|---|---|
| Highly precarious employment (n=4769) | (50.75%) 2420 | 2349 |
| Unemployed (n=253) | (40.28%) 102 | 151 |

Odds ratio 1.53, 95% CI 1.18 to 1.97, p=0.0013

| | Muscular pain lower limbs | No muscular pain lower limbs |
|---|---|---|
| Highly precarious employment (n=4769) | (37.46%) 1786 | 2983 |
| Unemployed (n=253) | (35.54%) 90 | 163 |

Odds ratio 1.08, 95% CI 0.83 to 1.41, p=0.5476

| | Injuries | No injuries |
|---|---|---|
| Highly precarious employment (n=4769) | (10.83%) 516 | 4253 |
| Unemployed (n=253) | (6.18%) 16 | 237 |

Odds ratio 1.80, 95% CI 1.07 to 3.00, p=0.0255

<table>
<tr><th>Reference/<br>Study design</th><th>Data source/<br>Population range</th><th>Poor/bad job definition<br>Outcome</th><th>Results from paper</th><th>Is any job better than no job?<br>Where OR has been calculated by OES exposure is bad/poor job, is unadjusted and data used to calculate may have been derived from percentages in study results tables</th></tr>
<tr><td>Minelli L et al. 2014<br><br>Cross sectional</td><td>Survey on Household Income, Italy, 2006 to 2010<br><br>15 to 64 years</td><td>Insecure employment (included apprenticeships, on-project jobs and seasonal jobs)<br>General health</td><td>Fixed effects (FE) and random effects (RE) ordered Logit models for self-reported health status M2 includes the interaction of employment condition and time. Coefficients and standard errors. Those significant at 10% level are in bold<br><br>
<table>
<tr><td></td><td>M1</td><td></td><td>M2</td><td></td></tr>
<tr><td>Ref: Permanent</td><td>FE</td><td>RE</td><td>FE</td><td>RE</td></tr>
<tr><td>Temporary</td><td>-0.147 (0.100)</td><td>-4.000 (0.058)</td><td>-0.159 (0.159)</td><td>-0.455 (0.095)</td></tr>
<tr><td>Unemployed</td><td>-0.317 (0.150)</td><td>-0.710 (0.086)</td><td>-0.366 (0.237)</td><td>-0.820 (0.143)</td></tr>
</table>
<br>Fixed-effects ordered logit models for self-reported health status by gender. Coefficients and standard errors. Those significant at 10% level are in bold. (table 4)<br><br>
<table>
<tr><td></td><td>Male</td><td></td><td>Female</td><td></td></tr>
<tr><td>Ref: Permanent</td><td>M1 (s.e)</td><td>M2 (s.e.)</td><td>M1 (s.e)</td><td>M2 (s.e)</td></tr>
<tr><td>Temporary</td><td>-0.288 (0.145)</td><td>-0.331 (0.234)</td><td>0.080 (0.163)</td><td>0.159 (0.247)</td></tr>
<tr><td>Unemployed</td><td>-0.423 (0.212)</td><td>-0.371 (0.326)</td><td>-0.160 (0.245)</td><td>-0.456 (0.385)</td></tr>
</table></td><td>Yes: Being unemployed had a significantly greater negative effect on perceived health when compared with being in temporary employment.<br><br>Data from table 1, page 4. Does not give number of participants so have used number of observations as stated in text
<table>
<tr><td>N =37,782 total observations</td><td>Poor/very poor health</td><td>Not very poor health</td></tr>
<tr><td>Temporary employment (7.72%) n= 2917</td><td>(0.07% + 1.20%) 37</td><td>2880</td></tr>
<tr><td>Unemployed (3.95%) n= 1492</td><td>(0.47% + 3.96) 66</td><td>1426</td></tr>
</table>
Odds ratio 0.27, 95% CI 0. 18 to 0.42 p<0.0001</td></tr>
<tr><td>Park SJ et al. 2020<br><br>Cross sectional</td><td>Korean National Health and Nutrition Survey, 2013 to 2017<br><br>15 to 59 years</td><td>Insecure employment<br><br>Mental health, general health, health behaviours</td><td>Association between employment status and health after adjusting for the explanatory variables. Odds ratios and 95% confidence intervals. Temporary job is reference<br><br>
<table>
<tr><td>Health related variable</td><td>Unemployed vs temporary All</td><td>Unemployed vs temporary Men</td><td>Unemployed vs temporary Women</td></tr>
<tr><td>Current smoking</td><td>0.88 (0.70 - 1.11)</td><td>0.90 (0.70 to 1.16)</td><td>0.90 (0.59 to 1.38)</td></tr>
<tr><td>High risky alcohol use</td><td>0.70 (0.54 to 0.91)</td><td>0.66 (0.47 to 0.92)</td><td>0.86 (0.58 to 1.27)</td></tr>
<tr><td>Mental health service use</td><td>1.49 (0.92 to 2.42)</td><td>2.33 (1.10 to 4.95)</td><td>1.10 (0.57 to 2.13)</td></tr>
<tr><td>Health related QOL (coefficient + SE)</td><td>-0.002 (0.002)</td><td>-0.003 (0.003)</td><td>-0.001 (0.003)</td></tr>
<tr><td>Perceived poor health</td><td>1.32 (1.06 to 1.65)</td><td>1.28 (0.92 to 1.89)</td><td>1.34 (0.98 to 1.83)</td></tr>
<tr><td>High daily perceived stress</td><td>0.85 (0.72 to 1.01)</td><td>0.84 (0.64 to 1.10)</td><td>0.89 (0.72 to 1.10)</td></tr>
<tr><td>Depressed mood</td><td>1.36 (1.02 to 1.81)</td><td>1.35 (0.85 to 2.14)</td><td>1.35 (0.93 to 1.97)</td></tr>
<tr><td>Depression symptoms</td><td>1.22 (0.76 to 1.97)</td><td>0.68 (0.26 to 1.77)</td><td>1.66 (0.93 to 1.97)</td></tr>
<tr><td>Ever been diagnosed with depression</td><td>0.85 (0.58 to 1.27)</td><td>1.19 (0.61 to 2.32)</td><td>0.65 (0.40 to 1.06)</td></tr>
<tr><td>Suicide ideation or planning</td><td>1.14 (0.73 to 1.77)</td><td>1.65 (0.87 to 3.14)</td><td>0.80 (0.43 to 1.47)</td></tr>
<tr><td>Suicide attempt</td><td>0.95 (0.30 to 3.04)</td><td>2.42 (0.52 to 11.13)</td><td>No result</td></tr>
</table>
<br>Adjusted for age, education, household income and marital status</td><td>No: Compared to those in temporary employment those who were unemployed were less likely to report current smoking, risky alcohol consumption, high daily perceived stress, to have ever been diagnosed with depression or to report a suicide attempt but more likely to report mental health service use, depression symptoms or suicide ideation or planning. Only the difference in reported alcohol consumption is likely to be statistically significant.<br><br>Yes: Compared to those in temporary employment, those who were unemployed were more likely to report perceived poor health, this difference may be statistically significant<br><br>Unemployed people were more like to report a lower health related quality of life score when compared with those in temporary employment</td></tr>
<tr><td>Scheuring S et al. 2021<br><br>Cohort</td><td>German Socio-Economic panel 1995 to 2017<br><br>18 to 65 years</td><td>Fixed term employment<br><br>Life satisfaction</td><td>Spill-over effects of fixed-term employment on the well-being of partners (life satisfaction), transition from unemployment to fixed-term employment. Beta coefficient (figure 1, page 11)<br><br>For transitions by men beta coefficient 0.30, error bars do not cross zero<br><br>For transition by women beta coefficient 0.08, error bars cross zero</td><td>Yes: For women their partners transition from unemployment to fixed term employment was associated with a significant improvement in their life satisfaction compared with their partners remaining unemployed<br><br>No: For men their partners transition from unemployment to fixed term employment was not associated with a significant improvement in their life satisfaction compared with their partner remaining unemployed</td></tr>
<tr><td>Sumner RC et al. 2020<br><br>Cross sectional</td><td>Understanding Society Survey, UK, 2010 to 2011<br><br>16 to 64 years</td><td>Insecure employment<br><br>Biomarkers C-reactive protein and fibrinogen</td><td>Multiple linear regressions for the association between employment subtypes and CRP and fibrinogen Reference is unemployment Beta coefficient and SE<br><br>
<table>
<tr><td></td><td>C-reactive protein</td><td>Fibrinogen</td></tr>
</table></td><td>No: There was no significant difference in C-reactive protein or fibrinogen levels between those who were unemployed and those employed in temporary jobs</td></tr>
</table>

<table>
<tr><th>Reference/<br>Study design</th><th>Data source/<br>Population range</th><th>Poor/bad job definition<br>Outcome</th><th>Results from paper</th><th>Is any job better than no job?<br>Where OR has been calculated by OES exposure is bad/poor job, is unadjusted and data used to calculate may have been derived from percentages in study results tables</th></tr>
<tr><td></td><td></td><td></td><td>Permanent -0.04 (0.08) p = 0.625 -0.13 (0.04) p = 0.004<br>Temporary -0.03 (0.13) p = 0.796 -0.04 (0.06) p = 0.467<br><br>Adjusted for individual, socio-economic and health behaviours and conditions characteristics</td><td></td></tr>
<tr><td>Van Aerden K, Gadeyne S, Vanrolen C. 2017<br><br>Cross sectional</td><td>Belgian Generations and Gender Study, 2008 to 2010<br><br>18 to 64 years</td><td>Precarious employment; unstable, low income and involuntary part-time employment<br><br>Mental health, general health</td><td>Relationship between labour market position and poor general health. Odds ratios and 95% confidence intervals. Reference is standard jobs<br><br>Precarious jobs 1.53 (1.04 to 2.26)*<br><br>Unemployed 1.85 (1.29 to 2.65)**<br><br>Relations between labour market position and poor mental health. Odds ratios and 95% confidence intervals. Reference is standard jobs<br><br>Precarious jobs 1.74 (1.10 to 2.75)*<br><br>Unemployed 2.70*** (1.77 to 4.13)<br><br>Adjusted for sex, age, household situation and social support<br>* p <0.05 ** p<0.01 *** p<0.001</td><td>No: Those who were unemployed were more likely to report poor general and mental health than those in precarious jobs, but these differences are unlikely to be statistically significant</td></tr>
<tr><td>Yoo et al. 2016<br><br>Cohort</td><td>Korean Welfare Panel Study 2008 to 2011<br><br>18 years and above</td><td>Insecure employment<br><br>Depression</td><td>Association between employment status change and depression. Odds ratios and 95% confidence intervals. Reference is permanent to permanent<br><br>Employment status<br>Precarious to precarious 1.54 (1.30 to 1.83)<br>Unemployment to unemployment 1.45 (1.23 to 1.70)<br>Unemployment to precarious 1.34 (1.07 to 1.68)<br>Precarious to unemployment 1.65 (1.32 to 2.06)<br><br>Association between employment status change and depression by sex and head of household. Odds rations and 95% confidence intervals). Reference is permanent to permanent<br><br>Men: Non-head of household, Head of household; Women: Non-head of household, Head of household<br><br>Precarious to precarious 2.36 (1.26 to 4.45) 1.47 (1.09 to 2.0) 1.26 (0.97 to 1.65) 2.07 (1.28 to 3.36)<br>Unemployment to unemployment 1.84 (1.02 to 3.32) 1.72 (1.21 to 2.45) 1.03 (0.81 to 1.30) 2.71 (1.63 to 4.51)<br>Unemployment to precarious 2.65 (1.29 to 5.45) 1.22 (0.74 to 2.02) 0.93 (0.66 to 1.29) 2.04 (1.12 to 3.69)<br>Precarious to unemployment 2.33 (1.11 to 4.91) 1.64 (1.05 to 2.58) 1.39 (0.99 to 1.95) 1.83 (1.02 to 3.30)<br><br>Adjusted for age, sex, marital status, region, education, self-rated health, smoking, alcohol, lag(CES-D) and year</td><td>No: Those who remained in precarious employment at baseline and follow up were more likely to have depression than those who were unemployed at baseline and follow up but the difference is unlikely to be significant. Those who moved from unemployment to precarious employment were more likely to have depression than those who moved from precarious employment to unemployment but this difference is unlikely to be statistically significant<br><br>(NB not limited to new onset depression but the analysis controlled for the previous years depression score)</td></tr>
<tr><td>Yoon et al. 2017<br><br>Cohort<br><br>This study not included in synthesis</td><td>Korean Welfare Panel Study 2011 to 2015<br><br>Age not given</td><td>Insecure employment<br><br>Suicidal ideation (among workers without lifetime suicide ideation at baseline)</td><td>Change in employment status and its association with suicidal ideation among workers without lifetime suicidal ideation at baseline. Odds ratios and 95% confidence intervals. N=3423. Reference is permanent employment at baseline and follow up.<br><br>Full time precarious 2.33 (1.09 to 4.99)*<br>Part time precarious 3.94 (1.46 to 10.64)*<br>Unemployed 1.89 (0.21 to 17.48)<br><br>* p<0.05<br><br>Adjusted for gender, age, marital status, equalised household income, residential area, chronic disease, disability and occupation and baseline depressive symptoms</td><td>No: In those who were in permanent employment at baseline those who were unemployed at follow up were less likely to report suicide ideation than those in full or part time precarious employment. This difference is not statistically significant.<br><br>Data from table 3, page 461<table><tr><th></th><th>Suicide ideation</th><th>No suicide ideation</th></tr><tr><td>Precarious employment (full and part time) n= 706</td><td>20</td><td>686</td></tr></table></td></tr>
</table>

| Reference/ Study design | Data source/ Population range | Poor/bad job definition Outcome | Results from paper | Is any job better than no job? Where OR has been calculated by OES exposure is bad/poor job, is unadjusted and data used to calculate may have been derived from percentages in study results tables |
|---|---|---|---|---|
| | | | | <table><tr><td>Unemployment n= 59</td><td>1</td><td>58</td></tr></table>Relative risk 1.67, 95% CI 0.23 to 12.24, p=0.6131 |

**Table S2: Results by objectivity of outcome measure**

| Reference | Outcome | Is any job better than no job? | | |
|---|---|---|---|---|
| | | Poor job significantly better | No significant difference | Poor job significantly worse |
| **OBJECTIVE MEASURES** | | | | |
| Chandola and Zhang 2018 | Allostatic load – biomarkers related to chronic stress | | | ✓ allostatic load transition into poor quality job vs remaining unemployed |
| Guseva Canu et al. 2019 | Suicide | ✓ death from suicide, lowest skilled and unskilled employees vs unemployed | | |
| Kim et al. 2020 | Biomarker C-reactive protein | | ✓ C-reactive protein unemployment vs temporary job | |
| Sumner et al. 2020 | Biomarkers C-reactive protein and fibrinogen | | ✓ C-reactive protein or fibrinogen, unemployment vs temporary job | |
| **VALIDATED MEASURES – SELF-REPORT** | | | | |
| Butterworth et al. 2013 | Common mental disorders (CIS-R) | | ✓ common mental disorders (CIS-R), poorest quality jobs vs unemployed | |
| Butterworth et al. 2011 | Mental health (MHI from SF-36) | | | ✓ decline in mental health over time (SF-36), poorest quality jobs vs unemployed |
| Chandola and Zhang 2018 | Physical and mental wellbeing (component scores from SF-36) | | ✓ SF-12 mental component score transition into poor quality work vs remaining unemployed<br><br>✓ GHQ-12 score transition into low quality job vs remaining unemployed | |
| Cortes-Franch et al. 2019 | Mental wellbeing (WHO-5 | ✓mental well-being (WHO-5) women, unemployed vs low quality job | ✓ mental well-being (WHO-5) men, unemployed vs low quality job | |

| Reference | Outcome | Is any job better than no job? | | |
|---|---|---|---|---|
| | | Poor job significantly better | No significant difference | Poor job significantly worse |
| | well-being index) | | | |
| Cortes-Franch et al. 2018 | Mental health (GHQ-12) | ✓ mental health (GHQ-12) men with a temporary contract vs unemployed (any length of time)<br><br>✓mental health (GHQ-12) men with no contract vs unemployed ≤ 2 years | ✓mental health (GHQ-12) men with no contract vs unemployed > 2 years<br><br>✓mental health (GHQ-12) women no or temporary contract vs any length of unemployment | |
| Fiori et al. 2016 | Mental health (SF 12-MHI) | | ✓ unable to establish significance, mental health (SF-12) fixed term or atypical contract vs looking for a new job | |
| Flint et al. 2013 | Psychological wellbeing GHQ-12) | ✓ lower level of psychological wellbeing (GHQ-12) spell of insecure employment vs spell of unemployment | | |
| Jang et al. 2015 | Severe depressive symptoms (CES-D) | ✓ severe depressive symptoms (CES-D) transition to precarious employment vs transition to unemployment | | |
| Yoo et al. 2016 | Depression (CES-D) | | ✓ depression (CES-D), precarious employment vs unemployment<br><br>✓ depression (CES-D), transition from unemployment to precarious employment vs transition from precarious employment to unemployment | |
| **Mixed – validated tool, self-report and subjective measures (not using validated tool)** | | | | |
| Gebel and Vosemer 2014 | Psychological and physical health (SF-12 and self-report) | ✓ psychological health (SF-12) transition from temporary work to unemployment | ✓ physical health (self-report) transition from temporary work to unemployment | |
| Grzywacz and Dooley 2003 | Physical health (self report) Depression (Composite International Diagnostic | ✓ for fair/poor health (self-report) inadequate employment vs unemployment | ✓ for depression (CIDI-SF) inadequate employment vs unemployment | |

| Reference | Outcome | Is any job better than no job? | | |
|---|---|---|---|---|
| | | Poor job significantly better | No significant difference | Poor job significantly worse |
| | Interview – short form) | | | |
| Inanc 2018 | Psychological wellbeing (GHQ-12) and life satisfaction (single survey question) | ✓ husbands own psychological well-being (GHQ-12) and life satisfaction (self-report) temporary employment vs unemployed<br><br>✓ wives own psychological well-being and life satisfaction temporary employment vs unemployed | ✓ impact of husbands temporary employment on wives psychological well-being and life satisfaction vs unemployed<br><br>✓ impact of wives temporary employment on husbands life satisfaction vs unemployed | ✓ impact of wives temporary employment on husbands psychological well-being vs unemployed |
| Park et al. 2020 | Mental health (PHQ-9), general health (EuroQol-5D, health behaviours (self report) | ✓ perceived poor health (EuroQol-5D) and health related quality of life temporary employment vs unemployment | ✓ self-reported smoking, high daily perceived stress, ever diagnosed with depression, depression symptoms (PHQ-9), suicide attempt, planning or ideation, mental health service use, temporary employment vs unemployment | ✓ risky alcohol consumption (self-report), temporary employment vs unemployment |
| **Subjective measures – self-report not using a validated tool** | | | | |
| Bently et al. 2020 | Smoking (yes/no), drinking (yes/no), risky drinking (number of drinks per week) | | ✓smoking, drinking and risky drinking insecure employment vs unemployment | |
| Fornell et al. 2018 | General health (self-report) | | ✓ self-reported bad health unemployment vs precarious employment | |
| Griep et al. 2016 | Psychological complaints, self-rated health, life satisfaction (all self report) | ✓ life satisfaction (self-report) insecure employment vs short or long term unemployment | ✓ psychological complaints (self-report), insecure employment vs long term unemployment<br><br>✓ self-rated health insecure employment vs short or long term unemployment | ✓ psychological complaints (self-report) insecure employment vs short term unemployment |

| Reference | Outcome | Is any job better than no job? | | |
|---|---|---|---|---|
| | | Poor job significantly better | No significant difference | Poor job significantly worse |
| | | | ✓ subjective complaints insecure employment vs long or short term employment | |
| Kim et al. 2019 | Suicidal ideation in the past year (single question) | ✓ suicide ideation remained in precarious employment vs those who remained unemployed<br><br>✓ suicide ideation remained in precarious employment vs those who moved from precarious employment to unemployment | ✓ suicide ideation transitioned from unemployment to precarious employment vs those who transitioned from precarious employment to unemployment<br><br>✓ suicide ideation transitioned from unemployment to precarious employment vs those who remained unemployed | |
| Maeda et al. 2019 | Insomnia related symptoms (self report) | ✓ insomnia related symptoms, not regularly employed vs unemployed | | |
| Matilla-Santander et al. 2020 | Bad health, health problems (self report) | ✓ for bad health status (self-report) unemployed vs precarious employment | ✓ for hearing problems, skin problems, anxiety, backache and muscular pain in lower limbs (self-report) unemployed vs precarious employment | ✓ for headaches or eyestrain, fatigue, muscular pain in upper limbs and injuries (self-report) unemployed vs precarious employment |
| Minelli et al 2014 | General health (self report) | ✓ perceived health (self-report), unemployed vs temporary employment | | |
| Scheuring et al. 2021 | Life satisfaction (self-report) | ✓ life satisfaction for women associated with their partners transition from unemployment to fixed term employment | ✓ life satisfaction for men associated with their partners transition from unemployment to fixed term employment | |
| Van Aerden, Gadeyne and Vanrolen. 2017 | Mental health, general health (both self report) | | ✓ general and mental health (self-report), precarious job vs unemployment | |

**Table S3: Results by welfare state regime**

| Reference | Is any job better than no job? | | |
|---|---|---|---|
| | **Poor job significantly better** | **No significant difference** | **Poor job significantly worse** |
| | **LIBERAL (UK AND IRELAND)** | | |
| Butterworth et al. 2013 | | ✓ common mental disorders (CIS-R), poorest quality jobs vs unemployed | |
| Chandola and Zhang 2018 | | ✓ SF-12 mental component score transition into poor quality work vs remaining unemployed<br><br>✓ GHQ-12 score transition into low quality job vs remaining unemployed | ✓ allostatic load transition into poor quality job vs remaining unemployed |
| Cortes-Franch et al. 2019 | ✓mental well-being (WHO-5) women, unemployed vs low quality job | ✓ mental well-being (WHO-5) men, unemployed vs low quality job | |
| Flint et al. 2013 | ✓ lower level of psychological wellbeing (GHQ-12) spell of insecure employment vs spell of unemployment | | |
| Inanc 2018 | ✓ husbands own psychological well-being (GHQ-12) and life satisfaction (self-report) temporary employment vs unemployed<br><br>✓ wives own psychological well-being and life satisfaction temporary employment vs unemployed | ✓ impact of husbands temporary employment on wives psychological well-being and life satisfaction vs unemployed<br><br>✓ impact of wives temporary employment on husbands life satisfaction vs unemployed | ✓ impact of wives temporary employment on husbands psychological well-being vs unemployed |
| Sumner et al. 2020 | | ✓ C-reactive protein or fibrinogen, unemployment vs temporary job | |
| | **SOUTHERN EUROPEAN (ITALY AND SPAIN)** | | |
| Cortes-Franch et al. 2018 | ✓ mental health (GHQ-12) men with a temporary contract<br><br>✓mental health (GHQ-12) men with no contract vs unemployed > 2 years | ✓mental health (GHQ-12) men with no contract vs unemployed ≤ 2 years<br><br>✓mental health (GHQ-12) women no or temporary contract any length of unemployment | |

| Reference | Is any job better than no job? | | |
|---|---|---|---|
| | Poor job significantly better | No significant difference | Poor job significantly worse |
| Fiori et al. 2016 | | ✓ unable to establish significance, mental health (SF-12) fixed term or atypical contract vs looking for a new job | |
| Fornell et al. 2018 | | ✓ self-reported bad health unemployment vs precarious employment | |
| Minelli et al. 2014 | ✓ perceived health (self-report), unemployed vs temporary employment | | |
| **CONSERVATIVE (SWITZERLAND, GERMANY AND BELGIUM)** | | | |
| Gebel and Vosemer 2014 | ✓ psychological health (SF-12) transition from temporary work to unemployment | ✓ physical health (self-report) transition from temporary work to unemployment | |
| Guseva Canu et al. 2019 | ✓ death from suicide, lowest skilled and unskilled employees vs unemployed | | |
| Scheuring et al. 2021 | ✓ life satisfaction for women partners transition from unemployment to fixed term employment | ✓ life satisfaction for men partners transition from unemployment to fixed term employment | |
| Van Aerden, Gadeyne and Vanrolen 2017 | | ✓ general and mental health (self-report), precarious job vs unemployment | |
| **SOCIAL DEMOCRATIC (FINLAND)** | | | |
| Griep et al. 2016 | ✓life satisfaction (self-report) insecure employment vs short or long term unemployment | ✓ psychological complaints (self-report), insecure employment vs long term unemployment<br><br>✓ self-rated health insecure employment vs short or long term unemployment<br><br>✓ subjective complaints insecure employment vs long or short term employment | ✓ psychological complaints (self-report) insecure employment vs short term unemployment |
